## Supplementary material for "Online randomised trials with children: A scoping review": S1 Checklist

### **Supporting Checklist S1:** PRISMA-ScR Checklist from “Online randomised trials with children: A scoping review.”

|  | **ITEM** | **PRISMA-ScR CHECKLIST ITEM** | **REPORTED ON PAGE #** |
| --- | --- | --- | --- |
| **TITLE** | | | |
| Title | 1 | Online Randomised Trials with Children: A Scoping Review | 1 |
| **ABSTRACT** | | | |
| Structured summary | 2 | Paediatric RCTs must content with many of the same challenges that all RCTs do, such as recruitment and retention to the trials. However, paediatric trials often bring additional obstacles in ethical considerations, funding, and publication. Decentralised trial methods are a promising strategy to help overcome some of those obstacles. Our scoping review searched the literature for randomised and quasi-randomised trials conducted online with people under 18, published in English. We found 21 trials that met our criteria. With the objectives of a) identifying what methods and tools had been used to create and conduct online trials with children and b) determine the gaps in the knowledge in this field, we charted the data and performed risk of bias assessments. This scoping review’s aim is to add to the knowledge base and improve trial methods so that online trials with children may be conducted to a higher standard with better inclusion and applicability. | 2-3 |
| **INTRODUCTION** | | | |
| Rationale | 3 | This scoping review will identify, describe, and characterize how online, decentralized trials are conducted with children in order to understand how they may be most effectively employed. | 3 |
| Objectives | 4 | This scoping review aims to determine how online, randomised trials with children are conducted. The objectives of this scoping review are: (a) to identify what methods and tools have been used to create and conduct online trials with children and (b) to determine the gaps in the knowledge in this field.  The population is children. The context is randomised and quasi-randomised trials. The concept is internet-based interventions. | 2, 4 |
| **METHODS** | | | |
| Protocol and registration | 5 | This protocol is published and project is registered at OSF. | [doi.org/10.12688/hrbopenres.13566.1](https://doi.org/10.12688/hrbopenres.13566.1)  [doi.org/10.17605/OSF.IO/WHJXY](http://www.doi.org/10.17605/OSF.IO/WHJXY) |
| Eligibility criteria | 6 | All randomised and quasi-randomised trials with children published in English conducted entirely online. We did not limit our search by date of publication. | 6 |
| Information sources* | 7 | Databases: MEDLINE, CENTRAL, CINAHL, Embase  Clinical Trial Registries: ICTRP, EU Clinical Trials Register, NIH Clinical Trials Register  Preprints: medRxiv, JMIR Preprints, HRB Open Research, and Advance from SAGE  Internet Searches with expert consultation | 5 |
| Search | 8 | The detailed search strategies for all searches are available in the supporting documents. | S2. & S3. Appendices |
| Selection of sources of evidence† | 9 | Pilot tests with two independent reviewers were conducted in order to achieve an inter-rater reliability of 0.75 using Cohen’s kappa. A single reviewer performed remaining screenings. This process was completed for both title/abstract screening and full-text screening. | 7-8 |
| Data charting process‡ | 10 | The data charting tool was modified from the version published in the protocol in an iterative manner by two review authors independently using one full-text study. After the tool was finalized the same two authors independently charted the data for 5 included studies and after, agreement, a single review author charted the data from all remaining studies (n = 15). | 8 |
| Data items | 11 | 1. Citation 2. Type of publication 3. Aim of study 4. Participant ages and mean age 5. Country(ies) of residence of participants 6. Number of participants randomised 7. Gender demographics of participants 8. SES of participants 9. Racial/ethnic demographics of participants 10. Specific participant population targeted 11. Steering/advisory group formation 12. Inclusion/exclusion criteria for participation 13. Online recruitment methods 14. Offline recruitment methods 15. Tools used for consent 16. Caregiver consent requirements 17. Consent validation requirements 18. Pushes/reminders 19. Number of arms 20. Intervention 21. Comparison 22. Technological devices needed for participation 23. Tools used for data protection processes 24. Methods used for data collection 25. Caregiver contribution of data 26. Duration of trial 27. Compensation offered to participants 28. Results relevant to this scoping review 29. Differences between completers and non-completers 30. Attrition rates of randomised participants 31. Limitations of study 32. Funding source 33. Other pertinent aspects of study 34. RoB1 assessment | S4 Form |
| Critical appraisal of individual sources of evidence§ | 12 | We assessed Risk of Bias using the RoB1 tool from the Cochrane Handbook. This was done in duplicate and discrepancies were resolved through discussion and the consultation of a third author when necessary. All included trials were either randomised or quasi-randomised and thus lent themselves to this type of appraisal. | 8 |
| Synthesis of results | 13 | The charted data were summarized and collated into tabular forms in order to characterise methods used in the included trials. | 8 |
| **RESULTS** | | | |
| Selection of sources of evidence | 14 | PRISMA 2020 flow diagram is completed and available in Fig1.  6,957 records screened, 6,550 records excluded, 407 full-text records retrieved and assessed for eligibility, 21 studies included in the scoping review. | 8-9 & Fig 1 |
| Characteristics of sources of evidence | 15 | The following data were charted for all included studies [citations: 1-21]: Basic characteristics: Country of trial conduct, Number of randomised participants, Age range of participants (mean age of participants), Intervention duration, Topic of study, Contribution of data by caregivers, Participant identified gender, Participant identified race and/or ethnicity  Recruitment methods: Online platforms used for recruitment, offline platforms used for recruitment  Consent: Methods of consent acquisition  Data collection: Online methods of data collection  Compensation: Methods and forms of compensation offered to participants  Loss to follow-up: Attrition rates of participants, methods to engage, retain, and enroll participants.  PPI input: Methods used to involve patients and the public  RoB assessment: Risk of bias assessed across seven domains as outlined in the Cochrane Group’s RoB1 tool. | 8-16, Tables 1-3, Figs 1 & 2 |
| Critical appraisal within sources of evidence | 16 | Risk of bias assessment summary and full reports completed using RoB1 tool from Cochrane.  ‘Low’ risk of bias was most common in the following domains: “random sequence generation” (n = 12), “selective reporting” (n = 14), and “other” bias (n = 17).  ‘Unclear’ risk of bias was most common in only one domain, “allocation concealment” (n =11).  ‘High’ risk of bias was most common in “blinding of participants and personnel” (n = 13), “blinding of outcome assessment” (n = 11), and “incomplete outcome data” (n = 11). | 16, Fig 2 & S7 Appendix |
| Results of individual sources of evidence | 17 | For each included source of evidence, the charted data is available in the Results section. | 9-16 & S7 Appendix |
| Synthesis of results | 18 | Summary of charted data and results are available under Results section. | 9-16, Tables 2 & 3, Fig 2 |
| **DISCUSSION** | | | |
| Summary of evidence | 19 | Summary of results and discussion in relation to the review questions are under Discussion section. | 16-20 |
| Limitations | 20 | The limitations of this scoping review were as listed: our search was limited to English language publications, after pilot-test title/abstract and full-text screening, one author completed the remaining screening; after pilot-testing of data charting, one author completed the remaining data charting process. | 20 |
| Conclusions | 21 | The methods, facilitators, and gaps in the knowledge of how online, randomised trials with children are conducted were identified and described; most trials used social media for multiple phases of their trials, although this may not always be the most appropriate platform for these methods. More in-depth consideration of target age groups, population, and PPI input should inform the design and conduct of these trials, and more thorough reporting of the methods is needed in future studies. | 21 |
| **FUNDING** | | | |
| Funding | 22 | This work contributes to one of the authors’ (SL) doctoral projects. SL’s PhD is funded by the Health Research Board-Trials Methodology Research Network in Ireland (grant ref: HRB-TMRN-2021-001) and the College of Medicine, Nursing and Health Sciences, University of Galway, Ireland. | 21 |
