## Supplementary material for "Online randomised trials with children: A scoping review": S2 Appendix

### **Supporting Appendix S2.** Database search strategies developed for MEDLINE (Ovid), CENTRAL, CINAHL (EBSCO), & Embase (Elsevier) for “Online randomised trials with children: A scoping review”

**MEDLINE (Ovid) search strategy conducted 16 February 2022.**

Ovid MEDLINE(R) ALL <1946 to February 16, 2022>

| 1 | (toddler$ or preschool$ or pre-school$ or preschool child$ or kindergarten$ or kinder-garten$ or child$ or kid$ or boy$ or girl$ or pediatric$ or paediatric$ or schoolage$ or school-age or school age$ or pre-teen$ or adolescen$ or teen$ or youth$ or young m#n$ or young wom#n$).tw,kw. |
| --- | --- |
| 2 | Child, Preschool/ |
| 3 | exp Child/ |
| 4 | Adolescent/ |
| 5 | or/1-4 |
| 6 | ((Internet$ or online$ or on-line$ or web$ or computer$ or digital$ or virtual$ or remote$ or decentrali$) adj3 (deliver$ or trial$ or intervention$ or study or studies or random$ or rct$)).tw,kw. |
| 7 | ((internet$ or online$ or on-line$ or web$ or computer$ or laptop or ipad or i-pad or digital$ or virtual$ or remote$ or technolog$ or software or smartphone$ or smart-phone$ or smart phone$ or mobile or iPhone or Android or cellphone$ or cell-phone$ or cell phone$ or social media or twitter or facebook or Instagram or snapchat) adj3 (application$ or app$ or deliver$ or intervention$ or treat$ or therap$ or program$)).tw,kw. |
| 8 | (ehealth or e-health or electronic health or mhealth or m-health or mobile health or telehealth or tele-health or telemedicine or tele-medicine).tw,kw. |
| 9 | Internet-Based Intervention/ |
| 10 | Therapy, Computer-Assisted/ |
| 11 | or/7-10 |
| 12 | 6 and 11 |
| 13 | randomized controlled trial.pt. |
| 14 | controlled clinical trial.pt. |
| 15 | randomi#ed.ab. |
| 16 | placebo$.ab. |
| 17 | drug therapy.fs. |
| 18 | randomly.ab. |
| 19 | trial.ab. |
| 20 | groups.ab. |
| 21 | or/13-20 |
| 22 | exp animals/ not humans.sh. |
| 23 | 21 not 22 |
| 24 | 5 and 12 and 23 |

**CENTRAL search strategy conducted 09 March 2022**

| #1 | MeSH descriptor: [Child, Preschool] this term only |
| --- | --- |
| #2 | MeSH descriptor: [Child] explode all trees |
| #3 | MeSH descriptor: [Adolescent] this term only |
| #4 | (toddler* or preschool* or pre-school* or preschool NEXT child* or kindergarten* or kinder-garten* or child* or kid* or boy* or girl* or pediatric* or paediatric* or schoolage* or school NEXT age* or pre-teen* or adolescen* or teen* or youth* or "young men" or "young man" or "young woman" or "young women"):ti,ab,kw |
| #5 | {or #1-#4} |
| #6 | ((Internet* or online* or on-line* or web* or computer* or digital* or virtual* or remote* or decentrali*) NEAR/3 (deliver* or trial* or intervention* or study or studies or random* or rct*)):ti,ab,kw |
| #7 | ((internet* or online* or on-line* or web* or computer* or laptop or ipad or i-pad or digital* or virtual* or remote* or technolog* or software or smartphone* or smart-phone* or smart NEXT phone* or mobile or iPhone* or Android* or cellphone* or cell-phone* or cell NEXT phone* or "social media" or twitter or facebook or Instagram or snapchat) NEAR/3 (application* or app* or deliver* or intervention* or treat* or therap* or program*)):ti,ab,kw |
| #8 | (ehealth or e-health or "electronic health" or mhealth or m-health or "mobile health" or telehealth or tele-health or telemedicine or tele-medicine):ti,ab,kw |
| #9 | MeSH descriptor: [Internet-Based Intervention] this term only |
| #10 | MeSH descriptor: [Therapy, Computer-Assisted] this term only |
| #11 | {OR #7-#10} |
| #12 | #11 AND #6 |
| #13 | #12 AND #5 in Cochrane Protocols, Trials |

**CINAHL (EBSCO) search strategy conducted 17 February 2022.**

| **#** | **Query** |
| --- | --- |
| S37 | S5 AND S34 AND S36 |
| S36 | S6 AND S35 |
| S35 | S7 OR S8 OR S9 OR S10 OR S11 |
| S34 | S33 NOT S32 |
| S33 | S12 OR S13 OR S14 OR S15 OR S16 OR S17 OR S18 OR S19 OR S20 OR S21 OR S22 OR S23 OR S24 OR S25 OR S26 |
| S32 | S30 NOT S31 |
| S31 | MH (human) |
| S30 | S27 OR S28 OR S29 |
| S29 | TI (animal model*) |
| S28 | MH (animal studies) |
| S27 | MH animals+ |
| S26 | AB (cluster W3 RCT) |
| S25 | MH (crossover design) OR MH (comparative studies) |
| S24 | AB (control W5 group) |
| S23 | PT (randomized controlled trial) |
| S22 | MH (placebos) |
| S21 | MH (sample size) AND AB (assigned OR allocated OR control) |
| S20 | TI (trial) |
| S19 | AB (random*) |
| S18 | TI (randomised OR randomized) |
| S17 | MH cluster sample |
| S16 | MH pretest-posttest design |
| S15 | MH random assignment |
| S14 | MH single-blind studies |
| S13 | MH double-blind studies |
| S12 | MH randomized controlled trials |
| S11 | (MH "Therapy, Computer Assisted") |
| S10 | MH ("internet-based intervention") |
| S9 | "internet-based intervention" or ehealth or "web-based intervention" or "electronic-health intervention" or "internet-based therapy" |
| S8 | ((TI ehealth OR AB ehealth) OR (TI e-health OR AB e-health) OR (TI "electronic health" OR AB "electronic health") OR (TI mhealth OR AB mhealth) OR (TI m-health OR AB m-health) OR (TI "mobile health" OR AB "mobile health") OR (TI telehealth OR AB telehealth) OR (TI tele-health OR AB tele-health) OR (TI telemedicine OR AB telemedicine) OR (TI tele-medicine OR AB tele-medicine)) |
| S7 | (((TI internet* OR AB internet*) OR (TI online* OR AB online*) OR (TI on-line* OR AB on-line*) OR (TI web* OR AB web*) OR (TI computer* OR AB computer*) OR (TI laptop OR AB laptop) OR (TI ipad OR AB ipad) OR (TI i-pad OR AB i-pad) OR (TI digital* OR AB digital*) OR (TI virtual* OR AB virtual*) OR (TI remote* OR AB remote*) OR (TI technolog* OR AB technolog*) OR (TI software OR AB software) OR (TI smartphone* OR AB smartphone*) OR (TI smart-phone* OR AB smart-phone*) OR (TI "smart phone*" OR AB "smart phone*") OR (TI mobile OR AB mobile) OR (TI iPhone OR AB iPhone) OR (TI Android OR AB Android) OR (TI cellphone* OR AB cellphone*) OR (TI cell-phone* OR AB cell-phone*) OR (TI "cell phone*" OR AB "cell phone*") OR (TI "social media" OR AB "social media") OR (TI twitter OR AB twitter) OR (TI facebook OR AB facebook) OR (TI Instagram OR AB Instagram) OR (TI snapchat OR AB snapchat)) N3 ((TI application* OR AB application*) OR (TI app* OR AB app*) OR (TI deliver* OR AB deliver*) OR (TI intervention* OR AB intervention*) OR (TI treat* OR AB treat*) OR (TI therap* OR AB therap*) OR (TI program* OR AB program*))) |
| S6 | (((TI Internet* OR AB Internet*) OR (TI online* OR AB online*) OR (TI on-line* OR AB on-line*) OR (TI web* OR AB web*) OR (TI computer* OR AB computer*) OR (TI digital* OR AB digital*) OR (TI virtual* OR AB virtual*) OR (TI remote* OR AB remote*) OR (TI decentral*? OR AB decentrali*)) N3 ((TI deliver* OR AB deliver*) OR (TI trial* OR AB trial*) OR (TI intervention* OR AB intervention*) OR (TI study OR AB study) OR (TI studies OR AB studies) OR (TI random* OR AB random*) OR (TI rct* OR AB rct*))) |
| S5 | S1 OR S2 OR S3 OR S4 |
| S4 | MH adolescence |
| S3 | (MH "Child, Preschool") OR (MH "Child Health") |
| S2 | MH child+ |
| S1 | ((TI toddler* OR AB toddler*) OR (TI preschool* OR AB preschool*) OR (TI pre-school* OR AB pre-school*) OR (TI "preschool child*" OR AB "preschool child*") OR (kindergarten* OR AB kindergarten*) OR (TI kinder-garten* OR AB kinder-garten*) OR (TI child* OR AB child*) OR (TI kid* OR AB kid*) OR (TI boy* OR AB boy*) OR (TI girl* OR AB girl*) OR (TI pediatric* OR AB pediatric*) OR (TI paediatric* OR AB paediatric*) OR (TI schoolage* OR AB schoolage*) OR (TI school-age OR AB school-age) OR (TI "school age*" OR AB "school age*") OR (TI pre-teen* OR AB pre-teen*) OR (TI adolescen* OR AB adolescen*) OR (TI teen* OR AB teen*) OR (TI youth* OR AB youth*) OR (TI "young m*n" OR AB "young m*n") OR (TI "young wom*n" OR AB "young wom*n")) |

**Embase (Elsevier) search strategy conducted 18 February 2022.**

| #31 | #5 AND #12 AND #30 |
| --- | --- |
| #30 | #13 OR #14 OR #15 OR #16 OR #17 OR #18 OR #19 OR #20 OR #21 OR #22 OR #23 OR #24  OR #25 OR #26 OR #27 OR #28 OR #29 |
| #29 | trial:ti |
| #28 | volunteer:ti,ab OR volunteers:ti,ab |
| #27 | (controlled NEAR/7 (study OR design OR trial)):ti,ab |
| #26 | assigned:ti,ab OR allocated:ti,ab |
| #25 | ((assign? OR match OR matched OR allocation) NEAR/5 (alternate OR group? OR intervention? OR patient? OR subject? OR participant?)):ti,ab |
| #24 | 'parallel group*':ti,ab |
| #23 | 'double blind procedure'/de |
| #22 | ((double OR single OR doubly OR singly) NEXT/1 (blind OR blinded OR blindly)):ti,ab |
| #21 | (open NEXT/1 label):ti,ab |
| #20 | (evaluated:ab OR evaluate:ab OR evaluating:ab OR assessed:ab OR assess:ab) AND (compare:ab OR compared:ab OR comparing:ab OR comparison:ab) |
| #19 | compare:ti OR compared:ti OR comparison:ti |
| #18 | placebo:ti,ab |
| #17 | 'intermethod comparison'/de |
| #16 | 'randomization'/de |
| #15 | random*:ti,ab |
| #14 | 'randomized controlled trial' |
| #13 | 'controlled clinical trial' |
| #12 | #10 AND #11 |
| #11 | ((internet* OR online* OR 'on line*' OR web* OR computer* OR digital* OR virtual* OR remote* OR decentrali*) NEAR/3 (deliver* OR trial* OR intervention* OR study OR studies OR random* OR rct*)):ti,ab,kw |
| #10 | #6 OR #7 OR #8 OR #9 |
| #9 | ehealth:ti,ab,kw OR 'e health':ti,ab,kw OR 'electronic health':ti,ab,kw OR mhealth:ti,ab,kw OR 'm health':ti,ab,kw OR 'mobile health':ti,ab,kw OR telehealth:ti,ab,kw OR 'tele health':ti,ab,kw OR telemedicine:ti,ab,kw OR 'tele medicine':ti,ab,kw |
| #8 | ((internet* OR online* OR 'on line*' OR web* OR computer* OR laptop OR ipad OR 'i pad' OR digital* OR virtual* OR remote* OR technolog* OR software OR smartphone* OR 'smart phone*' OR 'smart phone*' OR mobile OR iphone OR android OR cellphone* OR 'cell phone*' OR 'cell phone*' OR 'social media' OR twitter OR facebook OR instagram OR snapchat) NEAR/3 (application* OR app* OR deliver* OR intervention* OR treat* OR therap* OR program*)):ti,ab,kw |
| #7 | 'computer-assisted therapy'/de |
| #6 | 'web-based intervention'/de |
| #5 | #1 OR #2 OR #3 OR #4 |
| #4 | 'adolescent'/exp |
| #3 | 'preschool child'/de |
| #2 | 'child'/exp |
| #1 | toddler*:ti,ab,kw OR preschool*:ti,ab,kw OR 'preschool child*' OR 'pre school*':ti,ab,kw OR kindergarten*:ti,ab,kw OR 'kinder garten*':ti,ab,kw OR child*:ti,ab,kw OR kid*:ti,ab,kw OR boy*:ti,ab,kw OR girl*:ti,ab,kw OR pediatric*:ti,ab,kw OR paediatric*:ti,ab,kw OR schoolage*:ti,ab,kw OR 'school age':ti,ab,kw OR 'school age*':ti,ab,kw OR 'pre teen*':ti,ab,kw OR adolescen*:ti,ab,kw OR teen*:ti,ab,kw OR youth*:ti,ab,kw OR 'young woman*':ti,ab,kw OR 'young women*':ti,ab,kw OR 'young men*':ti,ab,kw OR 'young man*':ti,ab,kw |
