## Supplementary material for "Online randomised trials with children: A scoping review": S3 Appendix

### **Supporting Appendix S3.** Grey literature summary of search terms and strings used in grey literature searches conducted for “Online randomised trials with children: A scoping review”

All grey literature were searched iteratively using the original Medline search strategy to compose terms and strings.

#### **Original Medline search**

1	(toddler$ or preschool$ or pre-school$ or preschool child$ or kindergarten$ or kinder-garten$ or child$ or kid$ or boy$ or girl$ or pediatric$ or paediatric$ or schoolage$ or school-age or school age$ or pre-teen$ or adolescen$ or teen$ or youth$ or young m#n$ or young wom#n$).tw,kw.

2	Child, Preschool/

3	exp Child/

4	Adolescent/

5	or/1-4

6	((Internet$ or online$ or on-line$ or web$ or computer$ or digital$ or virtual$ or remote$ or decentrali$) adj3 (deliver$ or trial$ or intervention$ or study or studies or random$ or rct$)).tw,kw.

7	((internet$ or online$ or on-line$ or web$ or computer$ or laptop or ipad or i-pad or digital$ or virtual$ or remote$ or technolog$ or software or smartphone$ or smart-phone$ or smart phone$ or mobile or iPhone or Android or cellphone$ or cell-phone$ or cell phone$ or social media or twitter or facebook or Instagram or snapchat) adj3 (application$ or app$ or deliver$ or intervention$ or treat$ or therap$ or program$)).tw,kw.

8	(ehealth or e-health or electronic health or mhealth or m-health or mobile health or telehealth or tele-health or telemedicine or tele-medicine).tw,kw.

9	Internet-Based Intervention/

10	Therapy, Computer-Assisted/

##### **Lines and combinations of lines used for trial registry searching:**

1 toddler* or preschool* or pre-school* or preschool child* or kindergarten* or kinder-garten* or child* or kid* or boy* or girl* or pediatric* or paediatric* or schoolage or school-age or pre-teen or adolescen* or teen or youth or young men or young women
2 child
3 Adolescent

6.1	Internet* or online* or on-line* or web* or computer* or digital* or virtual* or remote* or decentrali*

6.2	deliver* or trial* or intervention* or study or studies or random* or rct*

7.1	internet* or online* or on-line* or web* or computer or laptop* or ipad or i-pad or digital* or virtual* or remote* or technolog* or software or smartphone* or smart-phone* or smart phone* or mobile or iPhone or Android or cellphone* or cell-phone* or social media or twitter or facebook or Instagram or snapchat

7.2	application* or app* or deliver* or intervention* or treat* or therap* or program*

8	ehealth or e-health or electronic health or mhealth or m-health or mobile health or telehealth or tele-health or telemedicine or tele-medicine

9	internet intervention

10	Computer-assisted therapy

#### **Trial Registries searched 10/04/2022**

##### **WHO ICTRP:**

Searches completed “In ALL Trials In ALL Recruiting Status”:

1 AND (6.1 AND 6.2): 30

1 AND (7.1 AND 7.2): 70

1 AND 6.1 OR 8: 3

1 AND 7.1 OR 8: 7

1 AND 8 : 31

1 AND 8 OR 9 OR 10: 1

(2 & 3 with OR) AND (6.1 AND 6.2): 5

(2 & 3 with OR) AND (7.1 AND 7.2): 13

(2 & 3 with OR) AND 6.1 OR 8: 0

(2 & 3 with OR) AND 7.1 OR 8: 1

(2 & 3 with OR) AND 8: 5

(2 & 3 with OR) AND 8 OR 9 OR 10: 0

=166

95 after de-duplication

##### **EU trial Registry:**

Searches completed:

6.1: 571

1 AND 6.1: 35

1 AND 7.1: 35

1 AND 6.1 OR 8: 35

1 AND 7.1 OR 8: 35

1 AND 8: 35

1 AND 8 OR 9 OR 10: 35

= 781

61 after de-duplication

##### **NIH CTR:**

Searches completed: “WITH results, population 0-17, including healthy participants”

Internet-based intervention: 13

Web intervention: 33

Decentraliz(s)ed: 0

Remote: 12

Social media: 7

Digital: 38

Computer-assisted therapy: 1

Virtual: 12

Smartphone: 10

e-health: 6

m-health: 6

telehealth: 10

telemedicine: 7

=155 results, 120 after de-duplication

#### **Preprint Searches:**

Search terms used: Internet-based intervention, web-based intervention, online intervention, children, adolescent, decentralized trial, decentralised trial

- Advance (Sage): 325 screened, 324 excluded- completed online 12/04/2022
- JMIR: 89 screened, 85 excluded- completed online 12/04/2022
- HRB Open Research: 116 screened, 116 excluded- completed online 12/04/2022
- medRXiv: 380 results; 379 after de-duplication

#### **Internet Searches:**

Search terms: Internet-based interventions with children, Web-based interventions with children, Online trials with children, Online-based interventions with children, Decentralized trials with children, Decentralised trials with children, Decentralized pediatric trials, Decentralised paediatric trials

- Google Searched 15/04/2022: 15 newly identified titles/abstracts
- Google Scholar Searched 15/04/2022: 8 newly identified titles/abstracts

Out of the 23 identified from the internet, 6 were removed for duplication or not meeting inclusion criteria, 17 internet results added to Covidence
