## Supplementary material for "Online randomised trials with children: A scoping review": S4 Appendix

### **Supporting Form S4.** Data charting tool for “Online randomised trials with children: A scoping review.”

| **Basic Study Characteristics** | |
| --- | --- |
| **Citation** |  |
| **Characteristic reported (indicate with X for decision or enter free text, where applicable)** | |
| Type of study | Randomised trial ☐ \| Cluster randomised ☐; Quasi-randomised trial ☐ \| Individually randomised ☐ |
| Type of publication | Peer-reviewed journal ☐; Pre-print ☐; Other, if so, describe ☐ |
| Aim of study as per author’s description |  |
| **Participant Characteristics** | |
| Participant age demographics | Age range \|; Mean age \| |
| Country(ies) of Participants’ Residence |  |
| Number of participants randomised | Children; Caregivers |
| Gender demographics of participants |  |
| Socioeconomic status of participants |  |
| Race/Ethnicity of participants |  |
| Type of participant targeted | (E.g. LQBTQ, disease-specific, racial/ethnic minority etc.) |
| Children’s advisory group formation | (Was an advisory group formed in the planning/execution of the study?) Yes ☐ No ☐ Unclear ☐ Not Applicable ☐ Not Reported ☐ Ages of advisory group participants \|; Means of recruiting advisory group participants \|; Phases of trial involvement \| |
| Inclusion/ Exclusion Criteria |  |
| **Methods of Recruitment, Retention & Consent** | |
| Online methods/platforms used for recruitment |  |
| Offline methods used for recruitment |  |
| Methods/tools used for online consent/assent acquisition |  |
| Caregiver consent acquired/ waived (If waived, by what authority?) | Yes ☐ No ☐ Unclear ☐ Not Applicable ☐ Not Reported ☐ |
| Children assent acquisition | Yes ☐ No ☐ Unclear ☐ Not Applicable ☐ Not Reported ☐ |
| What, if any, methods were used to validate that assent/consent was informed? | (E.g. validation questions, CAPTCHA, email confirmation, video-conferencing verbal) |
| Pushes/reminders sent to participants | Enrolment in trial Yes ☐ No ☐ Unclear ☐ Not Applicable ☐ Not Reported ☐; Retention/completion in trial Yes ☐ No ☐ Unclear ☐ Not Applicable ☐ Not Reported ☐; Follow-up in trial Yes ☐ No ☐ Unclear ☐ Not Applicable ☐ Not Reported ☐ |
| **Interventions** | |
| Number of arms |  |
| Intervention |  |
| Comparison |  |
| Operating Systems/Devices required for participants’ engagement with trial | (E.g. computer, phone, iOS, Windows, Android, iPhone, Facebook or Instagram account etc.) |
| Tools/software used for data protection processes | (E.g. dedicated website, REDCap, institutional server etc.) |
| Tools/methods used for data collection | (E.g. data submitted via website, video-conferencing etc.) |
| Did caregivers participate/ contribute data to the trial? If so, describe. | Yes ☐ No ☐ Unclear ☐ Not Applicable ☐ Not Reported ☐ |
| Duration of intervention from first to final engagement |  |
| Compensation offered to participants |  |
| **Outcomes** | |
| Results relevant to this scoping review | (E.g. satisfaction with online methods of trial, demographics of participants’ in relation to their outcomes especially in regarding online components, i.e., recruitment, retention, completion etc) |
| Differences between completers & non-completers |  |
| Attrition rates of randomised participants |  |
| Limitations of study as described by authors |  |
| Funding source | Institutional, i.e. governmental, university \| Yes ☐ No ☐ Unclear ☐ Not Applicable ☐ Not Reported ☐; Private, i.e. pharmaceutical, software company \| Yes ☐ No ☐ Unclear ☐ Not Applicable ☐ Not Reported ☐ |
| Other aspects of study pertinent to online trial conception and execution |  |

| Risk of Bias assessment  ([**https://handbook-5-1.cochrane.org/chapter_8/8_assessing_risk_of_bias_in_included_studies.htm**](https://handbook-5-1.cochrane.org/chapter_8/8_assessing_risk_of_bias_in_included_studies.htm)**)** | | | | |
| --- | --- | --- | --- | --- |
| Bias domain | Source of bias | Risk of bias | Support for judgment (use direct quotes where possible with explanatory comments) | Location in text or source (pg. number, figure, table etc.) |
| Selection bias | Random sequence generation | Low risk ☐    High risk ☐  Unclear risk ☐ |  |  |
|  | Allocation concealment | Low risk ☐    High risk ☐  Unclear risk ☐ |  |  |
| Performance bias | Blinding of participants and personnel | Low risk ☐    High risk ☐  Unclear risk ☐ |  |  |
| Detection bias | Blinding of outcome assessment | Low risk ☐    High risk ☐  Unclear risk ☐ |  |  |
| Attrition bias | Incomplete outcome data | Low risk ☐    High risk ☐  Unclear risk ☐ |  |  |
| Reporting bias | Selective reporting | Low risk ☐    High risk ☐  Unclear risk ☐ |  |  |
| Other bias | Anything else, ideally pre-specified | Low risk ☐    High risk ☐  Unclear risk ☐ |  |  |
