## Supplementary material for "Online randomised trials with children: A scoping review": S5 Table

### **Supporting Table S5.** Racial and ethnic minority distribution of participants by author and country in “Online randomised trials with children: A scoping review.”

| **Study (country)** | **Asian/ Asian Desi %** | **Black/ African American %** | **Latino(x)/ Hispanic %** | **Multi-racial/ other/ not listed**  **%** | **Native American/ Alaska Native**  **%** | **Native Hawaiian or other Pacific Islander**  **%** | **White % or non-Latino White %** | **Number of participants** |
| --- | --- | --- | --- | --- | --- | --- | --- | --- |
| Amsalem and Martin, 2022 (U.S.A.) | 8 | 23 | 21 | 9 | 2 | NR | 52 | 1,183 |
| Bragg et al., 2021 (U.S.A.) | NR | 46.50 | NR | NR | NR | NR | 53.50 | 832 |
| Dobias et al., 2021 (U.S.A.) | 7.26 | 9.73 | 21.06 | 2.12 | 5.49 | 1.59 | 75.04 | 565 |
| Egan et al., 2021 (U.S.A.) | 3.80 | 3.30 | 20.80 | 10 | NR | Combined with Asian | 62.10 | 240 |
| Greene et al., 2020 (U.S.A.) | 3 | 3 | 6 | 5 | 1 | Combined with Asian | 87 | 713 |
| Hillhouse et al., 2017 (U.S.A.) | NR | NR | 12.2 | 87.80 defined themselves as “Not Hispanic or Latino” | NR | NR | NR | 443 |
| Lester et al., 2019 (Canada, Columbia, U.S.A.) | NR | NR | 11.76 | 13.73 | NR | NR | 76.47 | 51 |
| Mogil et al., 2021 (U.S.A.)***** | 8.04/ 3.01 | 7.53/ 6.53 | 37.18/ 29.14 | 13.56/ 16.08 | 0.5/0 | 1.5/ 1 | 62.31/ 48.74 | 199 |
| Moreno et al., 2021 (U.S.A.) | NR | 14.67 | 9.27 | 8.55 | NR | NR | 67.56 | 1,520 |
| Nelson et al., 2022 (U.S.A.) | NR | 11 | 26 | 11 | NR | NR | 52 | 154 |
| O'Dea et al., 2020 (Australia) | NR | NR | NR | NR | NR | 3.62**^$^** | NR | 193 |
| Parker, Scull, & Morrison, 2022 (U.S.A.) | NR | 7.69 | 9 | 15.38 | 2.2 | NR | 74.73 | 91 |
| Schleider et al., 2022 (U.S.A.) | 12.64 | 10.48 | 19.21 | NR | 3.75 | 1.59 | 66.56 | 2,452 |
| Schwinn et al., 2015 (U.S.A.) | 6.4 | 7.3 | 12.8 | 7.4 | NR | NR | 66.1 | 236 |

NR= Not reported; NA= Not applicable

*= Data taken from parents, maternal data is listed first, paternal data is listed second.

$= Aboriginal or Torres Straight Islander

Where percentages do not equal 100, it was generally a case that Hispanic or Latino were not mutually exclusive from White/Caucasian as they can be considered an ethnicity and/or heritage, and the individual’s race may be listed differently. An individual may be Hispanic/ Latino and Black or Hispanic/Latino and White, etc., they also may list their ethnicity only as Hispanic/Latino.
