## Supplementary material for "Online randomised trials with children: A scoping review": S6 Table

### **Supporting Table S6.** Recruitment methods of participants reported for “Online randomised trials with children: A scoping review”.

| **Study (country)** | **Online platforms used for recruitment** | **Recruitment panel** | **Specific panel** | **Dedicated website** | **Other Online Advertising** | **Social media platform used for recruitment** | **Specific social media platform used for recruitment** | **Other social media platform used for recruitment (i.e. niche gaming sites)** | **Offline methods used for advertising recruitment to the trial** |
| --- | --- | --- | --- | --- | --- | --- | --- | --- | --- |
| Amsalem and Martin, 2022 (U.S.A.) | External online panel | Yes | Prime Panels | No | No | No | No | No | NA |
| Arnaud et al., 2016 (Belgium, Czech Republic, Germany, Sweden) | Multiple | No | NA | Yes | Yes | Yes | NR | No | Community |
| Bragg et al., 2021 (U.S.A.) | External online panel | Yes | Dynata | No | No | No | No | No | Telephone alerts via Dynata |
| Craig et al., 2016 (U.S.A) | Multiple | No | NA | No | Yes | Yes | NR | No | NA |
| Dobias et al., 2021 (U.S.A.) | Social media | No | NA | No | No | Yes | Instagram | No | NA |
| Egan et al., 2021 (U.S.A.) | Multiple | No | NA | No | Yes | Yes | Facebook, Instagram | Geeks OUT, Transmission Gaming, Reddit, Pitt+Me | NA |
| Ghaderi et al., 2020 (Sweden) | Multiple | No | NA | Yes | Yes | Yes | Facebook, Instagram | No | Schools |
| Greene et al., 2020 (U.S.A.) | Multiple | No | NA | Yes | Yes | Yes | Facebook | No | Community |
| Hillhouse et al., 2017 (U.S.A.) | External online panel | Yes | KnowledgePanel | No | No | No | No | No | NA |
| Kelleher, Moreno, & Wilt, 2018 (U.S.A.) | Social media | No | NA | No | No | Yes | Tumblr | No | NA |
| Lester et al., 2019 (Canada, Columbia, U.S.A.) | Email | No | NA | No | Yes, email only | No | No | No | NA |
| Manicavasagar et al., 2014 (Australia) | Multiple | No | NA | No | Yes | No | No | No | Community |
| Mogil et al., 2021 (U.S.A.) | Social media | No | NA | No | No | Yes | NR | NR | Community |
| Moreno et al., 2021 (U.S.A.) | External online panel | Yes | Qualtrics | No | No | No | No | No | NA |
| Nelson et al., 2022 (U.S.A.) | Social media | No | NA | No | No | Yes | Facebook, Instagram | No | NA |
| O'Connor et al., 2020 (Canada) | Multiple | No | NA | No | Yes | Yes | Facebook, Instagram, Twitter | No | Healthcare professionals, school-based teams |
| O'Dea et al., 2020 (Australia) | Social media | No | NA | No | No | Yes | Facebook | No | NA |
| Parker, Scull, & Morrison, 2022 (U.S.A.) | Social media | No | NA | No | Yes | Yes | NR | No | NA |
| Radomski et al., 2020 (Canada) | Social media | No | NA | No | No | Yes | Facebook, Instagram, Tumblr, Twitter | No | Healthcare professionals |
| Schleider et al., 2022 (U.S.A.) | Social media | No | NA | No | No | Yes | Instagram | No | NA |
| Schwinn et al., 2015 (U.S.A.) | Social media | No | NA | No | No | Yes | Facebook | No | NA |
