## Supplementary material for "Online randomised trials with children: A scoping review": S7 Appendix: Amsalem.docx

| **Basic Study Characteristics** | |
| --- | --- |
| **Citation** | Amsalem D, Martin A. Reducing depression‐related stigma and increasing treatment seeking among adolescents: randomized controlled trial of a brief video intervention. Journal of Child Psychology & Psychiatry. 2022;63(2):210-7. doi: 10.1111/jcpp.13427. PubMed PMID: 155253966. |
| **Characteristic reported (indicate with X for decision or enter free text, where applicable)** | |
| Type of study | Randomised trial ☒ \| Cluster randomised ☐; Quasi-randomised trial ☐ \| Individually randomised ☒ |
| Type of publication | Peer-reviewed journal ☒; Pre-print ☐; Other, if so, describe ☐ |
| Aim of study as per author’s description | To (a) reduce stigma toward depression and (b) increase treatment-seeking intentions among adolescents. |
| **Participant Characteristics** | |
| Participant age demographics | Age range \| 14-18; Mean age \| 16.8 +/- 1.2 |
| Country(ies) of Participants’ Residence | U.S.A. |
| Number of participants randomised | Children 1,183; Caregivers NA |
| Gender demographics of participants | 47% female |
| Socioeconomic status of participants | Not reported |
| Race/Ethnicity of participants | 52% white  23% African American  8% Asian  2% Native American  9% other  21% Hispanic |
| Type of participant targeted | NA |
| Children’s advisory group formation | Yes ☐ No ☐ Unclear ☐ Not Applicable ☐ Not Reported ☒ Ages of advisory group participants \| Not Reported; Means of recruiting advisory group participants \| Not Reported; Phases of trial involvement \| Not Reported |
| Inclusion/ Exclusion Criteria | Included: English speakers, 14-18 years of age, living in the U.S.  Excluded non-English speakers and non-U.S. residents |
| **Methods of Recruitment, Retention & Consent** | |
| Online methods/platforms used for recruitment | Prime Panels (crowdsourcing platform) |
| Offline methods used for recruitment | NA |
| Methods/tools used for online consent/assent acquisition | Before initiating the study, respondents reviewed an informed consent document. |
| Caregiver consent acquired/ waived (If waived, by what authority?) | Yes ☐ No ☐ Unclear ☐ Not Applicable ☐ Not Reported ☒ |
| Children assent acquisition | Yes ☒ No ☐ Unclear ☐ Not Applicable ☐ Not Reported ☐ |
| What, if any, methods were used to validate that assent/consent was informed? | Not reported |
| Pushes/reminders sent to participants | Enrolment in trial Yes ☐ No ☒ Unclear ☐ Not Applicable ☐ Not Reported ☐; Retention/completion in trial Yes ☐ No ☒ Unclear ☐ Not Applicable ☐ Not Reported ☐; Follow-up in trial Yes ☐ No ☒ Unclear ☐ Not Applicable ☐ Not Reported ☐ |
| **Interventions** | |
| Number of arms | 4 |
| Intervention | Group 1 (intervention): Viewed 102-113s video viewed of a female (girl) actor describing  difficulties coping with depressive symptoms, thoughts that life is not worth living, false assumptions about treatment and how and when they decided to seek help, how life has changed since tx.  Group 2 (Intervention): Viewed a 102-113s video of a male (boy) actor describing  difficulties coping with depressive symptoms, thoughts that life is not worth living, false assumptions about treatment and how and when they decided to seek help, how life has changed since tx.  Post intervention, the participants’ stigma toward depression was assessed using the Depression Stigma Scale (DSS) and treatment-seeking intentions using the General Help-Seeking Questionnaire (GHSQ). |
| Comparison | Group 3 (control): Viewed 100s video of a male actor (the same male actor as in the treatment videos) describing hobbies (sports, social media) and what they like to do with friends.  Group 4 (control): Viewed 100s video of a female actor (the same female actor as in the treatment videos) describing hobbies (sports social media) and what they like to do with friends.  Post video, the participants’ stigma toward depression was assessed using the Depression Stigma Scale (DSS) and treatment-seeking intentions using the General Help-Seeking Questionnaire (GHSQ). |
| Operating Systems/Devices required for participants’ engagement with trial | Not reported |
| Tools/software used for data protection processes | Qualtrics.com, a secure, online data-collection platform |
| Tools/methods used for data collection | Qualtrics.com, a secure, online data-collection platform |
| Did caregivers participate/ contribute data to the trial? If so, describe. | Yes ☐ No ☒ Unclear ☐ Not Applicable ☐ Not Reported ☐ |
| Duration of intervention from first to final engagement | Single session intervention (SSI)  Time for completion of questionnaires is not reported |
| Compensation offered to participants | $3.50 for the study |
| **Outcomes** | |
| Outcomes relevant to this review | Post hoc analysis showed videos with white protagonists had a greater influence on white participants. |
| Baseline differences between completers & non-completers | No demographic characteristic differences between completers and non-completers. |
| Attrition rates of randomised participants | N=474, completed 386 Intervention arm 1 (group 1): 18.6%  N= 474, completed 395 Intervention arm 2 (group 2): 16.6%  N= 235, completed 208 Control arms (group 3 & 4 were reported combined): 11.5% |
| Limitations of study as described by authors | No evaluation of longer-term effects (post-intervention assessment was immediate). Prime Panels participants may not be fully representative of general population. Two videos limited the ability to test influence of different races and ethnicities of multiple video protagonists on stigma reduction and treatment seeking. Only attitude were assessed, not actual treatment seeking. |
| Funding source | Institutional, i.e. governmental, university \| Yes ☒ No ☐ Unclear ☐ Not Applicable ☐ Not Reported ☐; Private, i.e. pharmaceutical, software company \| Yes ☐ No ☒ Unclear ☐ Not Applicable ☐ Not Reported ☐ |
| Other aspects of study pertinent to online trial conception and execution | Used multiple methods to exclude possible participants that did not meet inclusion criteria and to ensure the quality of the collected data:   1. Open-ended question format requiring participant’s age as a validity question   (Allowed only a two-digit number as a valid answer)   2. Used a CAPTCHA question to prevent bots 3. Timer added to ‘next’ button to ensure participants read instructions (5-second minimum) and for watching the video (100 seconds) 4. Scanned participants who attempted to answer the survey multiple times (less time than min expected, coordinates outside U.S., suspicious IP addresses) |

| Risk of Bias assessment  ([**https://handbook-5-1.cochrane.org/chapter_8/8_assessing_risk_of_bias_in_included_studies.htm**](https://handbook-5-1.cochrane.org/chapter_8/8_assessing_risk_of_bias_in_included_studies.htm)**)** | | | | |
| --- | --- | --- | --- | --- |
| Bias domain | Source of bias | Risk of bias | Support for judgment (use direct quotes where possible with explanatory comments) | Location in text or source (pg. number, figure, table etc.) |
| Selection bias | Random sequence generation | Low risk ☐    High risk ☐  Unclear risk ☒ | Quote: “We randomly and proportionally divided subjects into four study groups along a 4:4:1:1 ratio.” | Pg. 212, under Results |
|  | Allocation concealment | Low risk ☐    High risk ☐  Unclear risk ☒ | Comment: No information provided. | Not reported |
| Performance bias | Blinding of participants and personnel | Low risk ☐    High risk ☐  Unclear risk ☒ | Comment: Insufficient information to assess risk. | Not reported |
| Detection bias | Blinding of outcome assessment | Low risk ☐    High risk ☒  Unclear risk ☐ | Comment: Not reported. It is likely the outcome assessors were aware of intervention assignment. | Not reported |
| Attrition bias | Incomplete outcome data | Low risk ☒    High risk ☐  Unclear risk ☐ | Comment:  1,313 adolescents in baseline assessment  10% (130) excluded for failing validity tests  1,183 randomised  996 completed post-intervention assessment  =16% attrition rate overall  Intervention arm 1 (group 1): N=474, completed 386:18.6%  Intervention arm 2 (group 2): N= 474, completed 395: 16.6%  Control arms (group 3 & 4 were reported combined): N= 235, completed 208: 11.5% | Figure 1, pg. 212  Pg. 212 under Results |
| Reporting bias | Selective reporting | Low risk ☒    High risk ☐  Unclear risk ☐ | Comment:  Study protocol not available. Clinical Trials register report indicates the pre-specified outcomes were reported and analysed using the pre-specified methods. | Clinical Trials Register for this trial [NCT04760223](https://www.clinicaltrials.gov/ct2/show/NCT04760223) |
| Other bias | Anything else, ideally pre-specified | Low risk ☒    High risk ☐  Unclear risk ☐ | Comment: None detected | Not applicable |
