## Supplementary material for "Online randomised trials with children: A scoping review": S7 Appendix: Arnaud.docx

| Basic Study Characteristics | |
| --- | --- |
| **Citation** | Arnaud N, Baldus C, Elgán TH, De Paepe N, Tønnesen H, Csémy L, et al. Effectiveness of a Web-Based Screening and Fully Automated Brief Motivational Intervention for Adolescent Substance Use: a Randomized Controlled Trial. Journal of medical Internet research. 2016;18(5):e103. doi: 10.2196/jmir.4643. PubMed PMID: CN-01260475. |
| **Characteristic reported (indicate with X for decision or enter free text, where applicable)** | |
| Type of study | Randomised trial ☒ \| Cluster randomised ☐; Quasi-randomised trial ☐ \| Individually randomised ☒ |
| Type of publication | Peer-reviewed journal ☒; Pre-print ☐; Other, if so, describe ☐ |
| Aim of study as per author’s description | To evaluate an intervention aimed at reducing drinking and promoting abstinence from illicit drugs among adolescents in four European countries. |
| **Participant Characteristics** | |
| Participant age demographics | Age range \| 16-18; Mean age \| 16.8 |
| Country(ies) of Participants’ Residence | Sweden, Germany, Belgium, and the Czech Republic |
| Number of participants randomised | Children 1,449; Caregivers NA |
| Gender demographics of participants | 48.2% female |
| Socioeconomic status of participants | Parental education level in all randomized sample N=1449:  intervention/control  Low: 10.1/10.9  Middle: 61.4/66.3  High: 28.5/22.9 |
| Race/Ethnicity of participants | Not reported |
| Type of participant targeted | Positive CRAFFT (Car, Relax, Alone, Forget, Friends, Trouble) score on screening, indicating risk of drug and alcohol use. |
| Children’s advisory group formation | Yes ☒ No ☐ Unclear ☐ Not Applicable ☐ Not Reported ☐ Ages of advisory group participants \| Not Reported; Means of recruiting advisory group participants \| Not Reported; Phases of trial involvement \| “The WISEteens intervention was pilot-tested in two steps. First, 10 adolescents chose their preferred design concept among three options. A preliminary version with the preferred “look & feel” was then pre-tested by 37 other adolescents to ensure ease of registration and navigation use, comprehensibility of intervention content, satisfaction with layout and design, appropriateness of dialogue style (eg, avoiding judgmental and confronting language), overall satisfaction with the program, and time to complete baseline assessment and intervention. Furthermore, open feedback, technical problems, translational ambiguities, and other problems were documented and the program was adapted accordingly.” Pg. 6 |
| Inclusion/ Exclusion Criteria | Included: Online access, informed consent and a positive CRAFFT (Car, Relax, Alone, Forget, Friends, Trouble) score of 1 or more positive items, putting the participant at risk for drug and alcohol use. |
| **Methods of Recruitment, Retention & Consent** | |
| Online methods/platform used for recruitment | High rank of websites’ domain (WISEteens portal)  in widely used search engines, advertisements via popular social media, and links on affiliated health promotion sites. |
| Offline methods used for recruitment | Info leaflets, flyers distributed in schools, youth-clubs, cafes, bars, stores, and adolescent events |
| Methods/tools used for online consent/assent acquisition | “Informed online consent”  From protocol & primary article: Participants matching the inclusion criteria received study information including confidentiality advice (create an email that did not contain their name), the indication of voluntariness of participation, and of human subject protections. They were then asked to electronically give informed consent. |
| Caregiver consent acquired/ waived (If waived, by what authority?) | Yes ☐ No ☐ Unclear ☐ Not Applicable ☐ Not Reported ☒ |
| Children assent acquisition | Yes ☒ No ☐ Unclear ☐ Not Applicable ☐ Not Reported ☐ |
| What, if any, methods were used to validate that assent/consent was informed? | (E.g. validation questions, CAPTCHA, email confirmation, video-conferencing verbal)  No |
| Pushes/reminders sent to participants | Enrolment in trial Yes ☐ No ☒ Unclear ☐ Not Applicable ☐ Not Reported ☐; Retention/completion in trial Yes ☐ No ☒ Unclear ☐ Not Applicable ☐ Not Reported ☐; Follow-up in trial Yes ☒ No ☐ Unclear ☐ Not Applicable ☐ Not Reported ☐ Three months after completing the baseline assessment, participants were automatically invited to participate in the follow-up assessment and guided by an integrated hyperlink in the email invitation with one reminder email after 1 week. |
| **Interventions** | |
| Number of arms | 2 |
| Intervention | “The WISEteens intervention was a single session brief motivational intervention that relied on an interactive system to generate individually tailored content. All system-generated information was presented in small units that combined text and graphics (eg, photos and illustrative drawings) and directly referred to the participant’s statements assessed in the first place (eg, substance use, sex, weight, perceptions of normative drinking). Navigation through the program was designed as a dialogue between the user and a virtual expert with “gates” (ie, choice options) at the end of each page to permit varying degrees of approval or disapproval with page content. The system used these responses to introduce subsequent content on the next page.” Pg. 5 |
| Comparison | Control participants were randomised to assessment-only control group.  However, participants in the control group are provided with contact information on suitable counselling service providers to get in touch with in case of severe substance use-related problems. All participants are invited to visit again and participate in followup assessment after the evaluation period of the intervention of 3 months. |
| Operating Systems/Devices required for participants’ engagement with trial | (E.g. computer, phone, iOS, Windows, Android, iPhone, Facebook or Instagram account etc.)  Not reported |
| Tools/software used for data protection processes | Anonymous registration, user name, email address, password that did not contain names. From protocol: Participants were encouraged to create a new e-mail address, one that does not contain their name, in order to register with the dedicated website and take part. |
| Tools/methods used for data collection | Dedicated website (WISEteens)  (E.g. data submitted via website, video-conferencing etc.) |
| Did caregivers participate/ contribute data to the trial? If so, describe. | Yes ☐ No ☒ Unclear ☐ Not Applicable ☐ Not Reported ☐ |
| Duration of intervention from first to final engagement | Three months |
| Compensation offered to participants | Prize draw for tablet computers among participants who provided follow-up assessment. |
| **Outcomes** | |
| Results relevant to this scoping review | “Most adolescents were satisfied or totally satisfied with the system, design, comprehensibility, and intervention dialogue. Table 1 provides a summary of the pilot-results.” Pg. 6 |
| Baseline differences between completers & non-completers | “Overall baseline group comparisons thus indicate that the randomization was successful and that the completer-only subsample appears largely similar to the randomized sample.” Pg. 8  “Country of residence was the only significant predictor for dropout.” Pg.8 Significantly, more completers were women. |
| Attrition rates of randomised participants | “Response rates were very similar for the intervention group (15%) and the control group (14%)” Pg. 8  Intervention: 108/715 randomised  Control: 103/734 randomised  loss to follow-up at 3 months is therefore: 84.9 in intervention  86 in control |
| Limitations of study as described by authors | High dropout rate for follow-up assessment (although authors state they detected “no serious attrition bias”). “The inconsistent results of the imputed and non-imputed analyses and substantial between-analyses deviations in obtained effect sizes serve as a quantitative indicator of uncertainty in these results due to the substantial amount of missing follow-up data”. Pg. 16  Self-reported data. Participants were not blinded to assigned interventions. Relied on a convenience sample from the general population. |
| Funding source | Institutional, i.e. governmental, university \| Yes ☒ No ☐ Unclear ☐ Not Applicable ☐ Not Reported ☐; Private, i.e. pharmaceutical, software company \| Yes ☐ No ☒ Unclear ☐ Not Applicable ☐ Not Reported ☐ |
| Other aspects of study pertinent to online trial conception and execution | “All analyses are based on a complete-case dataset and an intention-to-treat (ITT) sample with imputation of missing follow-up data based on expectation maximization (EM). Both results are relevant and commonly reported in Web-based interventions particularly when dropout is large [70]. EM is a single imputation method that was shown to outperform the multiple imputation module available in SPSS in eHealth studies with high dropout rates [66].” Pg. 7 |

| Risk of Bias assessment  ([**https://handbook-5-1.cochrane.org/chapter_8/8_assessing_risk_of_bias_in_included_studies.htm**](https://handbook-5-1.cochrane.org/chapter_8/8_assessing_risk_of_bias_in_included_studies.htm)**)** | | | | |
| --- | --- | --- | --- | --- |
| Bias domain | Source of bias | Risk of bias | Support for judgment (use direct quotes where possible with explanatory comments) | Location in text or source (pg. number, figure, table etc.) |
| Selection bias | Random sequence generation | Low risk ☒    High risk ☐  Unclear risk ☐ | “Randomization was generated automatically by an online computer program without stratification.” | Pg. 5, under Procedure and Randomization |
|  | Allocation concealment | Low risk ☒    High risk ☐  Unclear risk ☐ | “Randomization was generated automatically by an online computer program without stratification.” | Pg. 5, under Procedure and Randomization |
| Performance bias | Blinding of participants and personnel | Low risk ☐    High risk ☒  Unclear risk ☐ | “Participants were not blinded to random allocation.” | Pg. 5, under Procedure and Randomization |
| Detection bias | Blinding of outcome assessment | Low risk ☐    High risk ☒  Unclear risk ☐ | Comment: No information provided on blinding of assessors but all data were self-reported and instruments used for this are standardized. | Pg. 7, under Substance Use |
| Attrition bias | Incomplete outcome data | Low risk ☒  High risk ☐  Unclear risk ☐ | Comment: Insufficient information to determine attrition for the control group. Attrition for the intervention group directly after the brief intervention was. 36.6% and 14% at the three-month follow-up.  Quote: “A total of 569 (28.1%) dropped out during the baseline assessment leaving 1449 participants who completed baseline assessment and were randomized to either the intervention (N=715) or control group (N=734). In the intervention group, 453 (63.4%) completed the brief intervention as measured by a log file record whether the last page of the intervention has been visible to the user. A total of 211 adolescents participated in the follow-up assessment after 3 months, corresponding to a valid response rate of 14.5%. In this subsample, the completion rate for the brief intervention was higher than in the full randomized sample (82.4%). “ | Pg. 8, under Results |
| Reporting bias | Selective reporting | Low risk ☒    High risk ☐  Unclear risk ☐ | Comment: The study protocol is available and all the study’s pre-specified (primary and secondary) outcomes have been reported in the trial. | Protocol available: <https://doi.org/10.1186/1471-2458-12-826> |
| Other bias | Anything else, ideally pre-specified | Low risk ☒    High risk ☐  Unclear risk ☐ | Comment: The study appears to be free of other sources of bias. | Under Limitations |
