## Supplementary material for "Online randomised trials with children: A scoping review": S7 Appendix: Bragg.docx

| **Basic Study Characteristics** | |
| --- | --- |
| **Citation** | Bragg M, Lutfeali S, Greene T, Osterman J, Dalton M  How Food Marketing on Instagram Shapes Adolescents’ Food Preferences: Online Randomized Trial  J Med Internet Res 2021;23(10):e28689  DOI: 10.2196/28689 |
| **Characteristic reported (indicate with X for decision or enter free text, where applicable)** | |
| Type of study | Randomised trial ☒ \| Cluster randomised ☐; Quasi-randomised trial ☐ \| Individually randomised ☒ |
| Type of publication | Peer-reviewed journal ☒; Pre-print ☐; Other, if so, describe ☐ |
| Aim of study as per author’s description | To examine whether adolescents could distinguish between food companies’ Instagram ads and their more traditional ads, and the extent to which Instagram food ads generate more appeal compared with traditional ads. |
| **Participant Characteristics** | |
| Participant age demographics | Age range \| 13-17; Mean age \| 14.73 |
| Country(ies) of Participants’ Residence | U.S.A. |
| Number of participants randomised | Children 832; Caregivers NA |
| Gender demographics of participants | 51.2% male, 48.8% female |
| Socioeconomic status of participants | Free- or reduced school lunch price program (indication of lower SES): 36.7% (yes); 13.3% (unsure) |
| Race/Ethnicity of participants | 46.5% Black/African American  53.5% non-Latino White |
| Type of participant targeted | Only Black and non-Latino White adolescents  (E.g. LQBTQ, disease-specific, racial/ethnic minority etc.) |
| Children’s advisory group formation | (Was an advisory group formed in the planning/execution of the study?) Yes ☐ No ☐ Unclear ☐ Not Applicable ☐ Not Reported ☒ Ages of advisory group participants \| Not Reported; Means of recruiting advisory group participants \| Not Reported; Phases of trial involvement \| Not Reported |
| Inclusion/ Exclusion Criteria | Excluded: Adolescents who identified as a race/ethnicity other than Black/African American or non-Latino White |
| **Methods of Recruitment, Retention & Consent** | |
| Online methods/platforms used for recruitment | Dynata. Dynata recurits research participants using online panels, digital networks and websites |
| Offline methods used for recruitment | SMS text messaging, and telephone alerts through Dynata |
| Methods/tools used for online consent/assent acquisition | Interested adolescents clicked on study advertisements, answered a brief web-based questionnaire and if eligible, completed consent. No specific tool reported. |
| Caregiver consent acquired/ waived (If waived, by what authority?) | Yes ☒ No ☐ Unclear ☐ Not Applicable ☐ Not Reported ☐  Caregiver consent acquired |
| Children assent acquisition | Yes ☒ No ☐ Unclear ☐ Not Applicable ☐ Not Reported ☐ |
| What, if any, methods were used to validate that assent/consent was informed? | Data integrity question required. “We included an attention check question (ie, “Type Facebook in the box”) to ensure participants were carefully reading directions and questions. Those who did not type “Facebook” were excluded from analysis.” Pg. 5  (E.g. validation questions, CAPTCHA, email confirmation, video-conferencing verbal) |
| Pushes/reminders sent to participants | Enrolment in trial Yes ☐ No ☒ Unclear ☐ Not Applicable ☐ Not Reported ☐; Retention/completion in trial Yes ☐ No ☒ Unclear ☐ Not Applicable ☐ Not Reported ☐; Follow-up in trial Yes ☐ No ☒ Unclear ☐ Not Applicable ☐ Not Reported ☐ |
| **Interventions** | |
| Number of arms | Survey 1: all participants completed same intervention  Survey 2: 2 arms, intervention and comparison |
| Intervention | Survey 1: “adolescents viewed 8 pairs of unhealthy food and beverage ads presented in random order (eg, a Starbucks magazine ad alongside a Starbucks Instagram ad). To remove cues indicating which ads originated from Instagram, we used Photoshop to compare advertising images with and without an “Instagram frame” which includes the Instagram logo, “likes,” and comments. After viewing each pair, adolescents answered questions” Pg.4-5.  Survey 2: “Adolescents were then randomly assigned to 1 of 2 conditions (Part 2) where ads ostensibly originated from (1) Instagram (ie, “labeled ad condition” in which advertising images have Instagram frames). The participants then answered survey questions to rate the ad on how much they liked the image, trendiness, artistic appeal, how delicious they thought the featured product might be, and how likely they were to purchase the product (Table 2)” Pg. 5. |
| Comparison | Comparison participants took part in survey 1, which was the same as the intervention participants.  Survey 2: Comparison participants viewed ads from traditional sources (unlabelled condition) and asked to rate the ad on how much they like the image, trendiness, artistic appeal, how delicious they thought the featured product might be, and how likely they were to purchase the product Fig.1, Pg. 4 |
| Operating Systems/Devices required for participants’ engagement with trial | Not reported  (E.g. computer, phone, iOS, Windows, Android, iPhone, Facebook or Instagram account etc.) |
| Tools/software used for data protection processes | Qualtics hosted the online surveys  (E.g. dedicated website, REDCap, institutional server etc.) |
| Tools/methods used for data collection | Qualtrics  (E.g. data submitted via website, video-conferencing etc.) |
| Did caregivers participate/ contribute data to the trial? If so, describe. | Yes ☐ No ☒ Unclear ☐ Not Applicable ☐ Not Reported ☐ |
| Duration of intervention from first to final engagement | Single session; median completion time of 19 minutes |
| Compensation offered to participants | Not reported |
| **Outcomes** | |
| Results relevant to this scoping review | NA  (E.g. satisfaction with online methods of trial, demographics of participants’ in relation to their outcomes especially in regarding online components, i.e., recruitment, retention, completion etc) |
| Differences between completers & non-completers | Not reported |
| Attrition rates of randomised participants | 6.5 %: Of the 1044 adolescents who started the survey, 976 completed. Not delineated by arms. |
| Limitations of study as described by authors | “We did not ask participants about their rationale for choosing the traditional ad or the Instagram ad in Part 1, and additional research is needed to determine whether differences between heavy and light users’ ratings translate into increased susceptibility to advertising. We did not collect data on self-reported height and weight, and it is possible that BMI could moderate the observed effects.” P. 10 |
| Funding source | Institutional, i.e. governmental, university \| Yes ☒ No ☐ Unclear ☐ Not Applicable ☐ Not Reported ☐; Private, i.e. pharmaceutical, software company \| Yes ☐ No ☒ Unclear ☐ Not Applicable ☐ Not Reported ☐ |
| Other aspects of study pertinent to online trial conception and execution | Gathered info on which social media adolescents used:  Instagram: 70%  Facebook: 85.3%  Snapchat: 49.3%  Tumblr:8.5%  Twitter: 47.4% |

| Risk of Bias assessment  ([**https://handbook-5-1.cochrane.org/chapter_8/8_assessing_risk_of_bias_in_included_studies.htm**](https://handbook-5-1.cochrane.org/chapter_8/8_assessing_risk_of_bias_in_included_studies.htm)**)** | | | | |
| --- | --- | --- | --- | --- |
| Bias domain | Source of bias | Risk of bias | Support for judgment (use direct quotes where possible with explanatory comments) | Location in text or source (pg. number, figure, table etc.) |
| Selection bias | Random sequence generation | Low risk ☐    High risk ☐  Unclear risk ☒ | Comment: Randomisation of participants into intervention and comparison groups was not reported in sufficient detail to make a risk assessment.    “Adolescents were then randomly assigned to 1 of 2 conditions (Part 2) where ads ostensibly originated from (1) Instagram (ie, “labeled ad condition” in which advertising images have Instagram frames); or (2) traditional sources (ie, “unlabeled ad condition” in which advertising images do not have Instagram frames.” | Pg. 5, under Survey Procedures |
|  | Allocation concealment | Low risk ☐    High risk ☐  Unclear risk ☒ | Comment: Insufficient information to assess risk. | Not reported |
| Performance bias | Blinding of participants and personnel | Low risk ☒    High risk ☐  Unclear risk ☐ | “Unbeknownst to participants, half of the ads in their condition originated from Instagram and half originated from traditional sources. Ads were presented in random order.” | Pg. 5, under Survey Procedures |
| Detection bias | Blinding of outcome assessment | Low risk ☐    High risk ☒  Unclear risk ☐ | Comment: No information provided on blinding of assessors but all data were self-reported. | Pgs. 8 & 9, under Statistical Analyses and Results |
| Attrition bias | Incomplete outcome data | Low risk ☒    High risk ☐  Unclear risk ☐ | “Of the 1044 adolescents who started the survey, 976 completed it, and 884 correctly answered our data integrity question (ie, “Type ‘Facebook’ in the box below.”). A total of 52 adolescents identified as a race/ethnicity other than Black/African American or non-Latino White and were excluded from the analyses.” | Pg. 2, under Methods |
| Reporting bias | Selective reporting | Low risk ☐    High risk ☐  Unclear risk ☒ | Comment: The study protocol is not available. There is insufficient information to assess risk. | Not reported |
| Other bias | Anything else, ideally pre-specified | Low risk ☒    High risk ☐  Unclear risk ☐ | Comment: No other bias detected. | Not applicable |
