## Supplementary material for "Online randomised trials with children: A scoping review": S7 Appendix: Craig.docx

| **Basic Study Characteristics** | |
| --- | --- |
| **Citation** | Craig A, Brown E, Upright J, DeRosier M. Enhancing Children's Social Emotional Functioning Through Virtual Game-Based Delivery of Social Skills Training. Journal of child and family studies. 2016;25(3):959-68. doi: 10.1007/s10826-015-0274-8. PubMed PMID: CN-02110005. |
| **Characteristic reported (indicate with X for decision or enter free text, where applicable)** | |
| Type of study | Randomised trial ☒ \| Cluster randomised ☐; Quasi-randomised trial ☐ \| Individually randomised ☒ |
| Type of publication | Peer-reviewed journal ☒; Pre-print ☐; Other, if so, describe ☐ |
| Aim of study as per author’s description | “The purpose of this study was to provide an initial evaluation of Zoo U as a first step in understanding how this game-based approach to SST can effectively improve children’s prosocial behaviors, social self-perceptions, and social literacy.” Pg. 965  *SST is Social Skills Training* |
| **Participant Characteristics** | |
| Participant age demographics | Age range \| 7-11; Mean age \| 9.65 |
| Country(ies) of Participants’ Residence | U.S.A. |
| Number of participants randomised | Children 59; Caregivers NA |
| Gender demographics of participants | 59% male |
| Socioeconomic status of participants | Not reported |
| Race/Ethnicity of participants | 28% racial or ethnic minority group and no significant differences in demographic variables across conditions |
| Type of participant targeted | No  (E.g. LQBTQ, disease-specific, racial/ethnic minority etc.) |
| Children’s advisory group formation | (Was an advisory group formed in the planning/execution of the study?) Yes ☐ No ☐ Unclear ☐ Not Applicable ☐ Not Reported ☒ Ages of advisory group participants \| Not Reported; Means of recruiting advisory group participants \| Not Reported; Phases of trial involvement \| Not Reported |
| Inclusion/ Exclusion Criteria | Included: English-language proficiency, access to an internet-enabled computer |
| **Methods of Recruitment, Retention & Consent** | |
| Online methods/platforms used for recruitment | Postings on parenting and educational listservs and social media sites targeted interested parents |
| Offline methods used for recruitment | NA |
| Methods/tools used for online consent/assent acquisition | “Informed consent/assent was obtained from all individual participants included in this study.” |
| Caregiver consent acquired/ waived (If waived, by what authority?) | Yes ☒ No ☐ Unclear ☐ Not Applicable ☐ Not Reported ☐  Caregiver consent acquired |
| Children assent acquisition | Yes ☒ No ☐ Unclear ☐ Not Applicable ☐ Not Reported ☐ |
| What, if any, methods were used to validate that assent/consent was informed? | “Prior to consenting (parents) and assenting (children) procedures, eligible parents and children viewed an online orientation video introducing them to the study design and procedures, and training them in the use of the project website through which they accessed all study materials.” Pg. 962  (E.g. validation questions, CAPTCHA, email confirmation, video-conferencing verbal) |
| Pushes/reminders sent to participants | Enrolment in trial Yes ☐ No ☒ Unclear ☐ Not Applicable ☐ Not Reported ☐; Retention/completion in trial Yes ☒ No ☐ Unclear ☐ Not Applicable ☐ Not Reported ☐; Follow-up in trial Yes ☐ No ☐ Unclear ☐ Not Applicable ☒ Not Reported ☐ Parents of children in the TX group were notified via email and through project website when a unit was available. |
| **Interventions** | |
| Number of arms | 2 |
| Intervention | “During the course of the trial, children completed a total of 30 Zoo U skill-building scenes. As Table 1 displays, Zoo U’s skill-building scenes are arranged into six units with five scenes of increasing difficulty within each unit.” “In keeping with the mastery model design of Zoo U, the scenes within each unit were delivered in progression; in order to advance, children had to either meet mastery criterion or play each scene three times, whichever came first.” Pg. 962 |
| Comparison | Control did not have access to Zoo U until the study trial period was over. |
| Operating Systems/Devices required for participants’ engagement with trial | Not reported  (E.g. computer, phone, iOS, Windows, Android, iPhone, Facebook or Instagram account etc.) |
| Tools/software used for data protection processes | Not reported  (E.g. dedicated website, REDCap, institutional server etc.) |
| Tools/methods used for data collection | Dedicated Zoo U website  (E.g. data submitted via website, video-conferencing etc.) |
| Did caregivers participate/ contribute data to the trial? If so, describe. | Yes ☒ No ☐ Unclear ☐ Not Applicable ☐ Not Reported ☐  Parents submitted baseline and post-intervention data. |
| Duration of intervention from first to final engagement | Ten weeks + 2-week follow-up |
| Compensation offered to participants | Not reported |
| **Outcomes** | |
| Results relevant to this scoping review | Not reported  (E.g. satisfaction with online methods of trial, demographics of participants’ in relation to their outcomes especially in regarding online components, i.e., recruitment, retention, completion etc) |
| Differences between completers & non-completers | Based on Chi square analyses there were no significant differences between both control and treatment in attrition |
| Attrition rates of randomised participants | 59 eligible participants were stratified, based on their BASC-2 scores, and randomly assigned to treatment (TX) or waitlist control (CO)  “Twenty percent of participants failed to respond to requests to complete post-tests, resulting in a final sample of 47 participants with complete data (TX = 23, CO = 24).”  Pg.961 Not reported how many were originally randomised into which group but attrition is between 16-20% for each group. |
| Limitations of study as described by authors | Small sample size.  Reliance on child and parent-report outcome measures of children’s social skills and social functioning.  The current study examined Zoo U’s effectiveness as implemented in the home; however, Zoo U is intended to be used both in schools and at home |
| Funding source | Institutional, i.e. governmental, university \| Yes ☒ No ☐ Unclear ☐ Not Applicable ☐ Not Reported ☐; Private, i.e. pharmaceutical, software company \| Yes ☐ No ☒ Unclear ☐ Not Applicable ☐ Not Reported ☐ |
| Other aspects of study pertinent to online trial conception and execution | NA |

| Risk of Bias assessment  ([**https://handbook-5-1.cochrane.org/chapter_8/8_assessing_risk_of_bias_in_included_studies.htm**](https://handbook-5-1.cochrane.org/chapter_8/8_assessing_risk_of_bias_in_included_studies.htm)**)** | | | | |
| --- | --- | --- | --- | --- |
| Bias domain | Source of bias | Risk of bias | Support for judgment (use direct quotes where possible with explanatory comments) | Location in text or source (pg. number, figure, table etc.) |
| Selection bias | Random sequence generation | Low risk ☐    High risk ☐  Unclear risk ☒ | Comment: Randomisation of participants into intervention and comparison groups was not reported in sufficient detail to make a risk assessment.    “Fifty-nine eligible participants were stratified by BASC-2 subscale scores, age, sex, and race, and then randomly assigned to either the treatment condition (TX) or the waitlist control condition (CO).” | Pg. 961, under Participants |
|  | Allocation concealment | Low risk ☐    High risk ☐  Unclear risk ☒ | Comment: Insufficient information to assess risk. | Not reported |
| Performance bias | Blinding of participants and personnel | Low risk ☐    High risk ☒  Unclear risk ☐ | “Another limitation of this study was reliance on child- and parent-report outcome measures of children’s social skills and social functioning.”  Comment: Participants in the intervention group would most likely be aware of their allocation to the intervention given the pre- and post-test assessments and interaction with the intervention and this is likely to have influenced the outcome. | Pg.966, under Limitations |
| Detection bias | Blinding of outcome assessment | Low risk ☐    High risk ☒  Unclear risk ☐ | “Inclusion of observational methods and additional reporters who are blind to treatment condition, such as teachers, would strengthen our understanding of how Zoo U impacts real-world behavior change and reduce concerns that results are influenced by the reporter’s knowledge that the child is receiving an intervention.”  Comment: There is no other mention of blinding by assessors in the publication and so based on the above quote, there was no assessor blinding and the outcome measurement is likely to be influenced by lack of blinding. | Pg.966, under Limitations |
| Attrition bias | Incomplete outcome data | Low risk ☐    High risk ☒  Unclear risk ☐ | “Fifty-nine eligible participants were stratified by BASC-2 subscale scores, age, sex, and race, and then randomly assigned to either the treatment condition (TX) or the wait- list control condition (CO). Children in the TX condition completed the Zoo U game over a 10-week period while children in the CO condition did not have access to Zoo U until the study trial period was over. Twenty percent of participants failed to respond to requests to complete posttests, resulting in a final sample of 47 participants with complete data (TX = 23, CO = 24).”  Therefore, attrition = 20% | Pg. 961, under Participants |
| Reporting bias | Selective reporting | Low risk ☐    High risk ☐  Unclear risk ☒ | Comment: The study protocol is not available. There is insufficient information to assess risk. | Not reported |
| Other bias | Anything else, ideally pre-specified | Low risk ☒    High risk ☐  Unclear risk ☐ | Comment: No other bias detected | Not applicable |
