## Supplementary material for "Online randomised trials with children: A scoping review": S7 Appendix: Dobias.docx

| **Basic Study Characteristics** | |
| --- | --- |
| **Citation** | Dobias ML, Schleider JL, Jans L, Fox KR. An online, single-session intervention for adolescent self-injurious thoughts and behaviors: results from a randomized trial. Behaviour research and therapy. 2021;147. doi: 10.1016/j.brat.2021.103983. PubMed PMID: CN-02342148. |
| **Characteristic reported (indicate with X for decision or enter free text, where applicable)** | |
| Type of study | Randomised trial ☒ \| Cluster randomised ☐; Quasi-randomised trial ☐ \| Individually randomised ☒ |
| Type of publication | Peer-reviewed journal ☒; Pre-print ☐; Other, if so, describe ☐ |
| Aim of study as per author’s description | “To test whether a half-hour, self-guided, online intervention designed to reduce the frequency of self-injurious thoughts and behaviors (“Project SAVE”) could reduce non-suicidal self-injury (NSSI) and negative self-beliefs.” Pg. 3 |
| **Participant Characteristics** | |
| Participant age demographics | Age range \| 13-16; Mean age \| 14.95 |
| Country(ies) of Participants’ Residence | U.S.A. |
| Number of participants randomised | Children 565; Caregivers NA |
| Gender demographics of participants | 37.35% identified with a gender other than that assigned at birth;  66.37% girl/woman; 19.29% as nonbinary; 10.62% as not sure |
| Socioeconomic status of participants | Not reported |
| Race/Ethnicity of participants | 75.04% White;  21.06% Hispanic/latinx;  9.73% Black/African American |
| Type of participant targeted | Children that engage in NSSI. Many ads were designed to reach LGBTQ+ youth. |
| Children’s advisory group formation | (Was an advisory group formed in the planning/execution of the study?) Yes ☐ No ☒ Unclear ☐ Not Applicable ☐ Not Reported ☐ However, participants in both conditions rated their respective online programs as acceptable, per pre-registered criteria. Ages of advisory group participants \| No; Means of recruiting advisory group participants \| No; Phases of trial involvement \| No |
| Inclusion/ Exclusion Criteria | Included: English-language proficiency, no self-reported learning disability, visual impairment or similar making answering computer questions difficult, access to an internet-enabled computer, laptop, or smartphone, engaged in NSSI in last month, disliking or hating themselves; additional criteria based on other questions |
| **Methods of Recruitment, Retention & Consent** | |
| Online methods/platforms used for recruitment | Advertisements on online social networking sites: Instagram. |
| Offline methods used for recruitment | NA |
| Methods/tools used for online consent/assent acquisition | Online assent after eligibility survey |
| Caregiver consent acquired/ waived (If waived, by what authority?) | Yes ☐ No ☒ Unclear ☐ Not Applicable ☐ Not Reported ☐  Waived by the University of Denver IRB |
| Children assent acquisition | Yes ☒ No ☐ Unclear ☐ Not Applicable ☐ Not Reported ☐ |
| What, if any, methods were used to validate that assent/consent was informed? | “to be included, participants met additional data quality criteria as indexed by their qualitative responses (i.e., did not respond with: direct copying/pasting survey text, lack of English fluency, random text, or three words or fewer when asked for two sentences or more; see study pre-registration for more information, https://osf.io/x5cd9). Although not formally pre-registered as an exclusion criterion, the present analysis also excluded participants who self-disclosed that they had skipped through or not read entire portions of the survey or online program.” Pg. 8  (E.g. validation questions, CAPTCHA, email confirmation, video-conferencing verbal) |
| Pushes/reminders sent to participants | Enrolment in trial Yes ☐ No ☐ Unclear ☐ Not Applicable ☐ Not Reported ☒; Retention/completion in trial Yes ☐ No ☐ Unclear ☐ Not Applicable ☐ Not Reported ☒; Follow-up in trial Yes ☐ No ☐ Unclear ☐ Not Applicable ☐ Not Reported ☒ |
| **Interventions** | |
| Number of arms | 2 |
| Intervention | Project SAVE (“Stop Adolescent Violence Everywhere”), a single-session intervention (SSI) is a ~30 minute, self-administered, web-based program to decrease self-injurious behaviours in youth with 4 content sections. The program also consists of a baseline, post-intervention, and follow-up questionnaire that encompassed demographic information, self-injurious thoughts and behaviours, self-hatred, and program feedback. |
| Comparison | Active Control: SSI Supportive theory was a web-based program that encourages feelings sharing (30 min self-administered online) |
| Operating Systems/Devices required for participants’ engagement with trial | computer, laptop, or smartphone  (E.g. computer, phone, iOS, Windows, Android, iPhone, Facebook or Instagram account etc.)  ads were on Instagram so most, if not all, participants would likely have an account |
| Tools/software used for data protection processes | “the study is completely confidential and done online” Pg. 14  (E.g. dedicated website, REDCap, institutional server etc.) |
| Tools/methods used for data collection | Qualtrics  (E.g. data submitted via website, video-conferencing etc.) |
| Did caregivers participate/ contribute data to the trial? If so, describe. | Yes ☐ No ☒ Unclear ☐ Not Applicable ☐ Not Reported ☐ |
| Duration of intervention from first to final engagement | SSI +Three month follow-up |
| Compensation offered to participants | “Participants earned up to $30 USD for their participation, via a 1-in-10 chance of winning $25 after the baseline survey plus a guaranteed $5 awarded upon follow-up survey completion.” Pg. 15 |
| **Outcomes** | |
| Results relevant to this scoping review | “Participants in both conditions rated their respective online programs as acceptable, per pre-registered criteria” A score of 4.16 (out of 5) with a SD of 0.56 from the full sample on the Program Feedback Scale. Table 3, Pg. 48  (E.g. satisfaction with online methods of trial, demographics of participants’ in relation to their outcomes especially in regarding online components, i.e., recruitment, retention, completion etc) |
| Differences between completers & non-completers | No significant differences were detected in dropout rates between the two groups |
| Attrition rates of randomised participants | Total sample = 565, 286 into SAVE intervention, and 279 into control (CO)  Immediate post-test: Intervention: 54/286 = 18.9% attrition; Control: 59/279 = 21.1% attrition  Follow-up:Intervention:58.04% attrition (no N given); Control: 61.29% attrition (no N given) P.21 |
| Limitations of study as described by authors | Generalizability of study due to sample demographics |
| Funding source | Institutional, i.e. governmental, university \| Yes ☒ No ☐ Unclear ☐ Not Applicable ☐ Not Reported ☐; Private, i.e. pharmaceutical, software company \| Yes ☐ No ☒ Unclear ☐ Not Applicable ☐ Not Reported ☐ |
| Other aspects of study pertinent to online trial conception and execution | Surpassed participation goal of 500 without their knowledge; ie. Instagram recruitment worked very quickly for them |

| Risk of Bias assessment  ([**https://handbook-5-1.cochrane.org/chapter_8/8_assessing_risk_of_bias_in_included_studies.htm**](https://handbook-5-1.cochrane.org/chapter_8/8_assessing_risk_of_bias_in_included_studies.htm)**)** | | | | |
| --- | --- | --- | --- | --- |
| Bias domain | Source of bias | Risk of bias | Support for judgment (use direct quotes where possible with explanatory comments) | Location in text or source (pg. number, figure, table etc.) |
| Selection bias | Random sequence generation | Low risk ☐    High risk ☐  Unclear risk ☒ | “per automated, 1:1 randomization in Qualtrics—a secure data collection software”  Comment: Insufficient information to make an assessment | Pg. 14, under Procedure |
|  | Allocation concealment | Low risk ☒    High risk ☐  Unclear risk ☐ | “As a result, both participants and study investigators were masked to participant condition throughout the entire data collection process.” | Pg. 14, under Procedure |
| Performance bias | Blinding of participants and personnel | Low risk ☐    High risk ☐  Unclear risk ☒ | “both participants and study investigators were masked to participant condition throughout the entire data collection process.”  Comment: Although masked to allocation, it is possible that participants would have been able to ascertain their group, once the intervention had been started as the Active Control did not address self-injurious behaviour, which was the study’s focus. | Pg. 14, under Procedure |
| Detection bias | Blinding of outcome assessment | Low risk ☐    High risk ☒  Unclear risk ☐ | Comment: All outcomes were self-reported. Blinding of assessors is not mentioned but it is likely that the outcome measurement was influenced by a lack of blinding. | Pg. 16, under Analytic Plan |
| Attrition bias | Incomplete outcome data | Low risk ☐    High risk ☒  Unclear risk ☐ | Comment: Total sample = 565, 286 into SAVE intervention, and 279 into control (CO)  Immediate post-test: Intervention: 54/286 = 18.9% attrition; Control: 59/279 = 21.1% attrition  Follow-up:Intervention:58.04% attrition (no N given); Control: 61.29% attrition (no N given) P.21  Comment: An attrition rate of 18.9% and 21.1% at an immediate post-test suggests a high risk of attrition bias. | Pg. 21, under Dropout and Acceptability of Online Programs |
| Reporting bias | Selective reporting | Low risk ☒    High risk ☐  Unclear risk ☐ | Comment: The study protocol is not available but the trial is registered at <https://clinicaltrials.gov/ct2/show/NCT04498143>. The published report includes all expected outcomes, including those that were pre-specified.  “Second, we evaluated effects of the Project SAVE intervention on pre-registered outcomes of interest, relative to the control group. In accordance with our pre-registration, we checked for differential dropout by condition and imputed missing data prior to assessing intervention effects.” | Pg. 17, under Intervention Effects on Study Outcomes  Pg. 19, under Deviations from Pre-Registration |
| Other bias | Anything else, ideally pre-specified | Low risk ☒    High risk ☐  Unclear risk ☐ | Comment: No other bias detected. | Not applicable |
