## Supplementary material for "Online randomised trials with children: A scoping review": S7 Appendix: Egan.docx

| **Basic Study Characteristics** | |
| --- | --- |
| **Citation** | Egan JE, Corey SL, Henderson ER, Abebe KZ, Louth-Marquez W, Espelage D, et al. Feasibility of a Web-Accessible Game-Based Intervention Aimed at Improving Help Seeking and Coping Among Sexual and Gender Minority Youth: results From a Randomized Controlled Trial. Journal of adolescent health. 2021;69(4):604-14. doi: 10.1016/j.jadohealth.2021.03.027. PubMed PMID: CN-02287880. |
| **Characteristic reported (indicate with X for decision or enter free text, where applicable)** | |
| Type of study | Randomised trial ☒ \| Cluster randomised ☐; Quasi-randomised trial ☐ \| Individually randomised ☒ |
| Type of publication | Peer-reviewed journal ☒; Pre-print ☐; Other, if so, describe ☐ |
| Aim of study as per author’s description | To test the feasibility of a game-based intervention to increase help-seeking (knowledge, intentions, self-efficacy, and behaviours), productive coping skills use, and coping flexibility and reduce health risk factors and behaviours among Sexual and Gender Minority Youth (SGMY). |
| **Participant Characteristics** | |
| Participant age demographics | Age range \| 14-18; Mean age \| 15.7 |
| Country(ies) of Participants’ Residence | U.S.A. |
| Number of participants randomised | Children 240; Caregivers NA |
| Gender demographics of participants | Cisgender girl: 16.3%  Cisgender boy: 36.7%  Gender minority: 47.1% |
| Socioeconomic status of participants | Free- or reduced school lunch price program (indication of lower SES): 36.7% (yes); 13.3% (unsure) |
| Race/Ethnicity of participants | White: 62.1%  Latinx: 20.8%  Asian or Pacific Islander: 3.8%  Black: 3.3%  Multiracial: 10% |
| Type of participant targeted | SGMY: Sexual and Gender Minority Youth  (E.g. LQBTQ, disease-specific, racial/ethnic minority etc.) |
| Children’s advisory group formation | (Was an advisory group formed in the planning/execution of the study?) Yes ☒ No ☐ Unclear ☐ Not Applicable ☐ Not Reported ☐ Ages of advisory group participants \| Not Reported; Means of recruiting advisory group participants \| Not Reported; Phases of trial involvement \| Conception: Before game development, one-on-one, in-depth interviews with 20 SGMY were conducted re their gaming preferences. As per the published protocol Coulter et al., 2019 |
| Inclusion/ Exclusion Criteria | Included: English literate, residing in the U.S.A., 14-18 years of age, experienced bullying/cyberbullying victimization in the past year, had a sexual minority identity or a gender minority identity, had appropriate technology for game, and an email address. |
| **Methods of Recruitment, Retention & Consent** | |
| Online methods/platforms used for recruitment | Advertisements on social media: Facebook, Instagram, SGM-related web-based gaming groups (Geeks OUT, Gay Geeks (FB group), GaymerX (FB group),Transmission Gaming, and Reddit Gaymer forums), Pitt+Me (Univ of Pittsburgh community for trials), dedicated FB page; Depending on the prior week’s enrolment numbers, we tailored which ads were used for the upcoming week. |
| Offline methods used for recruitment | NA |
| Methods/tools used for online consent/assent acquisition | Adolescents interested in taking part in the study clicked on a study advertisement and were directed to a web-based screening questionnaire and consent form. |
| Caregiver consent acquired/ waived (If waived, by what authority?) | Yes ☐ No ☐ Unclear ☐ Not Applicable ☐ Not Reported ☒ |
| Children assent acquisition | Yes ☒ No ☐ Unclear ☐ Not Applicable ☐ Not Reported ☐ |
| What, if any, methods were used to validate that assent/consent was informed? | Not reported  (E.g. validation questions, CAPTCHA, email confirmation, video-conferencing verbal) |
| Pushes/reminders sent to participants | Enrolment in trial Yes ☐ No ☒ Unclear ☐ Not Applicable ☐ Not Reported ☐; Retention/completion in trial Yes ☒ No ☐ Unclear ☐ Not Applicable ☐ Not Reported ☐; Follow-up in trial Yes ☒ No ☐ Unclear ☐ Not Applicable ☐ Not Reported ☐ |
| **Interventions** | |
| Number of arms | 2 |
| Intervention | Intervention participants received a list of national SGM-inclusive resources via email the day after randomisation as well as the link to download the role-playing game, Singularities, which incorporated three primary components: encouraging help-seeking behaviours, encouraging the use of productive coping, and raising awareness of web-based resources. |
| Comparison | Control participants received a list of national SGM-inclusive resources via email the day after randomization, additionally, after the completion of the final T3 survey, control participants were offered a free download of the intervention game. |
| Operating Systems/Devices required for participants’ engagement with trial | All laptops, desktops were acceptable for game download and play. No OS or type specified. Phones (no type specified) to complete surveys were also accepted.  (E.g. computer, phone, iOS, Windows, Android, iPhone, Facebook or Instagram account etc.) |
| Tools/software used for data protection processes | REDCap and Text files containing milestones achieved, time played, and player choices were automatically sent via a secure File Transfer Protocol (FTP) system to a secure server. Participants’ game play data were tracked using a unique identification number that did not rely on identifiable information.  (E.g. dedicated website, REDCap, institutional server etc.) |
| Tools/methods used for data collection | Via REDCap  (E.g. data submitted via website, video-conferencing etc.) |
| Did caregivers participate/ contribute data to the trial? If so, describe. | Yes ☐ No ☒ Unclear ☐ Not Applicable ☐ Not Reported ☐ |
| Duration of intervention from first to final engagement | Intervention lasted for 16 weeks per participant but was open from April to October, 2018. |
| Compensation offered to participants | $10 for T1, $25 for T2, $50 for T3 |
| **Outcomes** | |
| Outcomes relevant to this review | (E.g. satisfaction with online methods of trial, demographics of participants’ in relation to their outcomes especially in regarding online components, i.e., recruitment, retention, completion etc)  From Table 4:  **Time to enrol 240 participants:** 4 months  **Recruitment venues:** 2146 clicks from Facebook and Instagram; 3 from Reddit, 4 from Pitt+me  **How many youth were excluded for not having an email address:** 32  **Among intervention group: Would participants like to play the game on other platforms, such as phone?:** 55.4% both phone and computer; 29.2% phone only; 15.4% computer only |
| Differences between completers & non-completers | “There were no significant demographic differences between the intervention and control groups, as hypothesized.” Pg 610 |
| Attrition rates of randomised participants | Attrition \| Intervention \| Control; T1 n (%) \| 120 (100%) \| 120 (100%); T2 n (%) \| 47 (39.2%) \| 30 (25%); T3 n (%) \| 47 (39.2%) \| 39 (32.5%) |
| Limitations of study as described by authors | “The biggest concern from this study is that only half of the intervention group reported downloading/ playing the game. In addition, retention rates at each time point were lower than hoped (75%-80%), but we were able to maintain about 60% retention over time, suggesting the need for additional reengagement measures to improve retention.”  Pg. 612 |
| Funding source | Institutional, i.e. governmental, university \| Yes ☒ No ☐ Unclear ☐ Not Applicable ☐ Not Reported ☐; Private, i.e. pharmaceutical, software company \| Yes ☐ No ☒ Unclear ☐ Not Applicable ☐ Not Reported ☐ |
| Other aspects of study pertinent to online trial conception and execution | 2153 clicked to screening questionnaire, 2146 from FB and Instagram, 3 from Reddit, 4 from Pitt+me |

| Risk of Bias assessment  ([**https://handbook-5-1.cochrane.org/chapter_8/8_assessing_risk_of_bias_in_included_studies.htm**](https://handbook-5-1.cochrane.org/chapter_8/8_assessing_risk_of_bias_in_included_studies.htm)**)** | | | | |
| --- | --- | --- | --- | --- |
| Bias domain | Source of bias | Risk of bias | Support for judgment (use direct quotes where possible with explanatory comments) | Location in text or source (pg. number, figure, table etc.) |
| Selection bias | Random sequence generation | Low risk ☒    High risk ☐  Unclear risk ☐ | Quote: “We used permuted block randomization with equal allocation (using block sizes of 2, 4, 6, 8, and 10) to randomize participants.” | Pg. 606, under Randomization |
|  | Allocation concealment | Low risk ☐    High risk ☐  Unclear risk ☒ | Quote: “The randomization schema was created using the ralloc package for Stata and implemented in REDCap using the Randomization Module” | Pg. 606, under Randomization |
| Performance bias | Blinding of participants and personnel | Low risk ☐    High risk ☒  Unclear risk ☐ | Quote: “Participants and researchers were unblinded to intervention assignments.” | Pg. 606, under Randomization |
| Detection bias | Blinding of outcome assessment | Low risk ☐    High risk ☒  Unclear risk ☐ | Primary, secondary, and tertiary Outcomes:  Comment: All data were self-reported. | Table 1, pg. 607  Table 3, pg. 610 |
| Attrition bias | Incomplete outcome data | Low risk ☐    High risk ☒  Unclear risk ☐ | Comment:  Attrition at T2 32.1% (77/240)  Attrition at T3 35.8% (86/240)  240 total participants, 120 randomised to both intervention and control; at | Table 1, pg. 607  Figure 1, pg. 608 |
| Reporting bias | Selective reporting | Low risk ☒    High risk ☐  Unclear risk ☐ | Comment:  Study protocol is available and pre-specified outcomes are reported on. | Pg. 3 under Study Aims from the study protocol, Coulter et al., 2019. |
| Other bias | Anything else, ideally pre-specified | Low risk ☐    High risk ☐  Unclear risk ☒ | Quote:  “Our study may have social desirability bias. Despite using primarily validated self-report measures, in most instances, the scales were not specifically validated among SGMY, thereby potentially leading to measurement bias.” | Pg. 612 from Limitations |
