## Supplementary material for "Online randomised trials with children: A scoping review": S7 Appendix: Ghaderi.docx

| **Basic Study Characteristics** | |
| --- | --- |
| **Citation** | Ghaderi A, Stice E, Andersson G, Enö Persson J, Allzén E. A randomized controlled trial of the effectiveness of virtually delivered Body Project (vBP) groups to prevent eating disorders. Journal of consulting and clinical psychology. 2020;88(7):643-56. doi: 10.1037/ccp0000506. PubMed PMID: CN-02140414. |
| **Characteristic reported (indicate with X for decision or enter free text, where applicable)** | |
| Type of study | Randomised trial ☒ \| Cluster randomised ☐; Quasi-randomised trial ☐ \| Individually randomised ☒ |
| Type of publication | Peer-reviewed journal ☒; Pre-print ☐; Other, if so, describe ☐ |
| Aim of study as per author’s description | “To investigate the effectiveness of Body Project groups delivered virtually (vBP) by peer educators for prevention of eating disorders.” Pg. 643 |
| **Participant Characteristics** | |
| Participant age demographics | Age range \| 15-20; Mean age \| 17.3 |
| Country(ies) of Participants’ Residence | Sweden |
| Number of participants randomised | Children 443; Caregivers NA |
| Gender demographics of participants | 100% female |
| Socioeconomic status of participants | Not reported |
| Race/Ethnicity of participants | Not reported |
| Type of participant targeted | Females with body image issues |
| Children’s advisory group formation | (Was an advisory group formed in the planning/execution of the study?) Yes ☐ No ☒ Unclear ☐ Not Applicable ☐ Not Reported ☐ Ages of advisory group participants \| No; Means of recruiting advisory group participants \| No; Phases of trial involvement \| No |
| Inclusion/ Exclusion Criteria | Included: Females with body image concerns, fluency in Swedish.  Excluded: “Current Diagnostic and Statistical Manual for Mental Disorders-Fifth Edition diagnosis of eating disorders with the exception of unspecified eating disorders, concurrent psychological treatment, severe depression, suicidality, or other serious conditions that required psychiatric care.” Pg.645 |
| **Methods of Recruitment, Retention & Consent** | |
| Online methods/platforms used for recruitment | Dedicated website, ads and banners on websites. Facebook & Instagram advertisements. |
| Offline methods used for recruitment | High school principals in Sweden were contacted and asked to put up ads. Psychology students were sent to schools as project ambassadors. |
| Methods/tools used for online consent/assent acquisition | Informed consent given over a secure platform, iTerapi. |
| Caregiver consent acquired/ waived (If waived, by what authority?) | Yes ☐ No ☒ Unclear ☐ Not Applicable ☐ Not Reported ☐  Waived (not needed for >/=15 year olds in Sweden) by the Regional Ethics Board in Stockholm |
| Children assent acquisition | Yes ☒ No ☐ Unclear ☐ Not Applicable ☐ Not Reported ☐ |
| What, if any, methods were used to validate that assent/consent was informed? | Not reported  (E.g. validation questions, CAPTCHA, email confirmation, video-conferencing verbal) |
| Pushes/reminders sent to participants | Enrolment in trial Yes ☐ No ☒ Unclear ☐ Not Applicable ☐ Not Reported ☐; Retention/completion in trial Yes ☐ No ☒ Unclear ☐ Not Applicable ☐ Not Reported ☐; Follow-up in trial Yes ☒ No ☐ Unclear ☐ Not Applicable ☐ Not Reported ☐ |
| **Interventions** | |
| Number of arms | 3 |
| Intervention | 1) The Body Project was delivered through virtual groups (through Google Hangouts app), and consisted of four weekly 1-hr sessions across four consecutive weeks. 2) An expressive writing alternative intervention (EW) consisted of brief written instructions sent to participants weekly over a four-week period. |
| Comparison | A waitlist control condition who were offered the vBP after 6 months. |
| Operating Systems/Devices required for participants’ engagement with trial | Not reported  (E.g. computer, phone, iOS, Windows, Android, iPhone, Facebook or Instagram account etc.) |
| Tools/software used for data protection processes | Consent was done via iTerapi, a high security platform. All data were encrypted. virtual meetings could be done anonymously.  (E.g. dedicated website, REDCap, institutional server etc.) |
| Tools/methods used for data collection | Dedicated Website  (E.g. data submitted via website, video-conferencing etc.) |
| Did caregivers participate/ contribute data to the trial? If so, describe. | Yes ☐ No ☒ Unclear ☐ Not Applicable ☐ Not Reported ☐ |
| Duration of intervention from first to final engagement | 4 weeks and 24-month follow-up |
| Compensation offered to participants | No |
| **Outcomes** | |
| Results relevant to this scoping review | NA  (E.g. satisfaction with online methods of trial, demographics of participants’ in relation to their outcomes especially in regarding online components, i.e., recruitment, retention, completion etc) |
| Differences between completers & non-completers | “Drop-out was significant, and a marked limitation of the study,  by decreasing the internal validity and power of the study. Detailed dropout analysis (for the entire sample, and on each condition separately) did not reveal any statistically or clinically meaningful differences between the drop-out group and the completers.” Pg.654 |
| Attrition rates of randomised participants | “In total, 68 (92%) of the 74 in the vBP who still were in the trial, and 75 (85%) of 88 in the EW completed the booster assignment.” Pg. 647 **Group** \| **Random-ised** \| **Immediate Post-treatment** \| **6- month FU** \| **12-month FU** \| **18-month FU** \| **24- month FU** \| **% attrition**; vBP \| 149 \| 100 \| 97 \| 77 \| 71 \| 74 \| 50%; EW \| 148 \| 97 \| 92 \| 88 \| 91 \| 78 \| 47%; Waitlist \| 146 \| 112 \| 100 \| na \| na \| na \| 31.5% |
| Limitations of study as described by authors | The EW intervention was not group-based to match the format of the vBP |
| Funding source | Institutional, i.e. governmental, university \| Yes ☒ No ☐ Unclear ☐ Not Applicable ☐ Not Reported ☐; Private, i.e. pharmaceutical, software company \| Yes ☐ No ☒ Unclear ☐ Not Applicable ☐ Not Reported ☐ |
| Other aspects of study pertinent to online trial conception and execution | “recruitment was slow” “We increased the age range from 15–18 to 15–20 years to have a broader recruitment base” Use of a media company after slow recruitment was noted was employed, which increased recruitment. Pg. 646 |

| Risk of Bias assessment  ([**https://handbook-5-1.cochrane.org/chapter_8/8_assessing_risk_of_bias_in_included_studies.htm**](https://handbook-5-1.cochrane.org/chapter_8/8_assessing_risk_of_bias_in_included_studies.htm)**)** | | | | |
| --- | --- | --- | --- | --- |
| Bias domain | Source of bias | Risk of bias | Support for judgment (use direct quotes where possible with explanatory comments) | Location in text or source (pg. number, figure, table etc.) |
| Selection bias | Random sequence generation | Low risk ☒    High risk ☐  Unclear risk ☐ | “At the final stage, 443 were eligible and randomized using a list obtained from the Research Randomizer (www.randomizer.org).” | Pg. 645-646, under Study Design and Participants |
|  | Allocation concealment | Low risk ☒    High risk ☐  Unclear risk ☐ | “As soon as a set of three participants were ready for enrolment, the research coordinator sent their codes to the PI, who declared the condition for each participant according to the randomisation list.” | Pg. 645-646, under Study Design and Participants |
| Performance bias | Blinding of participants and personnel | Low risk ☐    High risk ☒  Unclear risk ☐ | “The website, ads and banners that were used for recruitment described the trial as a comparison of body acceptance interventions. To reach the target population (i.e., young females with body image concerns), two recurrent questions in all recruitment material were: “Do you have body image concerns?” and “Are you dissatisfied with your body?”  “Those randomized to the expressive writing or waitlist condition were so informed” | Pg. 646, under Procedure |
| Detection bias | Blinding of outcome assessment | Low risk ☒    High risk ☐  Unclear risk ☐ | “Follow-up clinical interviews were done by trained research assistants who were blind to condition allocation. Some facilitators who implemented the vBP were involved in the baseline assessment only, before the participants were allocated to a condition.”  “Two undergraduate students from another university with no prior involvement in the study received training to do fidelity ratings according to the Body Project Session Adherence, and Group Leader Competence Assessment. Raters were provided with eight training sessions to rate and to compare with the rating of the first author until the raters achieved high agreement with each other (intraclass correlation ⫽ .94) and the first author.” | Pg. 646, under Study Design and Participants  Pg. 648, under Fidelity: Adherence and Competence |
| Attrition bias | Incomplete outcome data | Low risk ☐    High risk ☒  Unclear risk ☐ | Comment; See Fig. 1. 33% immediate post-test attrition for vBP, 34.5% immediate post-test attrition for EW are high attrition rates for immediate post-test responses. | Pg. 645, under Methods |
| Reporting bias | Selective reporting | Low risk ☒    High risk ☐  Unclear risk ☐ | Comment: The study protocol is not available but the clinical trials registry is and it shows that all pre-specified outcomes have been reported in the pre-specified way | Registered at ClinicalTrials.gov: [NCT02567890](https://clinicaltrials.gov/ct2/show/NCT02567890) |
| Other bias | Anything else, ideally pre-specified | Low risk ☒    High risk ☐  Unclear risk ☐ | Comment: No other bias detected. | Not applicable |
