## Supplementary material for "Online randomised trials with children: A scoping review": S7 Appendix: Greene.docx

| **Basic Study Characteristics** | |
| --- | --- |
| **Citation** | Greene K, Ray AE, Choi HJ, Glenn SD, Lyons RE, Hecht ML. Short term effects of the REAL media e-learning media literacy substance prevention curriculum: an RCT of adolescents disseminated through a community organization. Drug and alcohol dependence. 2020;214:108170. doi: 10.1016/j.drugalcdep.2020.108170. PubMed PMID: CN-02140538. |
| **Characteristic reported (indicate with X for decision or enter free text, where applicable)** | |
| Type of study | Randomised trial ☒ \| Cluster randomised ☐; Quasi-randomised trial ☐ \| Individually randomised ☒ |
| Type of publication | Peer-reviewed journal ☒; Pre-print ☐; Other, if so, describe ☐ |
| Aim of study as per author’s description | To evaluate the short-term effects of testing an e-learning program to reduce adolescent substance use and abuse. |
| **Participant Characteristics** | |
| Participant age demographics | Age range \| 12-17; Mean age \| 14.71 |
| Country(ies) of Participants’ Residence | U.S.A. |
| Number of participants randomised | Children 713 randomised; 639 completed the baseline survey and was the ITT value; Caregivers Not reported |
| Gender demographics of participants | 66% female; 34% male |
| Socioeconomic status of participants | Not reported |
| Race/Ethnicity of participants | White: 87%;  African American or Black: 3%,  Asian or Pacific Islander: 3%,  American Indian or Alaskan Native: 1%,  other: 5%,  Hispanic: 6% |
| Type of participant targeted | 4-H children (a program where children complete hands-on projects in areas like health, science, agriculture, and civic engagement) |
| Children’s advisory group formation | Ages of advisory group participants \| “high school-aged youths in grades 9 and 10” Ray et al. Pg. 3 (Developmental paper for Greene et al.); Means of recruiting advisory group participants \| 4-H club leaders were recruited via email and they shared the recruitment flyer with teen members; Phases of trial involvement \| Focus groups, pilot testing and usability testing Yes ☒ No ☐ Unclear ☐ Not Applicable ☐ Not Reported ☐ (Was an advisory group formed in the planning/execution of the study?) Information from Ray et al., 2019 (Development paper) |
| Inclusion/ Exclusion Criteria | Included: Only age criteria, members of 4-H |
| **Methods of Recruitment, Retention & Consent** | |
| Online methods/platforms used for recruitment | Dedicated website, Facebook page, presentations via video conference, online streaming technologies at club events |
| Offline methods used for recruitment | 4H leaders were recruited, county leaders distributed to club leaders or youth. Flyers, as well as presentations in person and via telephone |
| Methods/tools used for online consent/assent acquisition | Parental consent forms were returned via email, mail, fax, text and through the project website. |
| Caregiver consent acquired/ waived (If waived, by what authority?) | Yes ☒ No ☐ Unclear ☐ Not Applicable ☐ Not Reported ☐  As above |
| Children assent acquisition | Yes ☒ No ☐ Unclear ☐ Not Applicable ☐ Not Reported ☐ |
| What, if any, methods were used to validate that assent/consent was informed? | Children provided assent at each online survey  (E.g. validation questions, CAPTCHA, email confirmation, video-conferencing verbal) |
| Pushes/reminders sent to participants | Enrolment in trial Yes ☐ No ☐ Unclear ☐ Not Applicable ☐ Not Reported ☒; Retention/completion in trial Yes ☐ No ☐ Unclear ☐ Not Applicable ☐ Not Reported ☒; Follow-up in trial Yes ☐ No ☐ Unclear ☐ Not Applicable ☐ Not Reported ☒ |
| **Interventions** | |
| Number of arms | 2 |
| Intervention | The program required youth to proceed sequentially through 5 levels that cover media reach, media ethics, influence strategies, advertising claims and evidence, and production techniques, teaching youth to critique, plan, produce, and disseminate ads. In the fifth level, they planned their anti-substance use message, which they produced offline and submitted to a social media contest. |
| Comparison | Delayed use control |
| Operating Systems/Devices required for participants’ engagement with trial | Not reported  (E.g. computer, phone, iOS, Windows, Android, iPhone, Facebook or Instagram account etc.) |
| Tools/software used for data protection processes | The project employed a Data Safety and Monitoring Board (DSMB) consisting of three members who reviewed procedures and monitored compliance  (E.g. dedicated website, REDCap, institutional server etc.) |
| Tools/methods used for data collection | Individual survey links were sent to participating youth via email or text.  (E.g. data submitted via website, video-conferencing etc.) |
| Did caregivers participate/ contribute data to the trial? If so, describe. | Yes ☐ No ☒ Unclear ☐ Not Applicable ☐ Not Reported ☐ |
| Duration of intervention from first to final engagement | 3 weeks + 3-month follow-up survey |
| Compensation offered to participants | $10 for completing each of the three online study surveys |
| **Outcomes** | |
| Results relevant to this scoping review | NA  (E.g. satisfaction with online methods of trial, demographics of participants’ in relation to their outcomes especially in regarding online components, i.e., recruitment, retention, completion etc) |
| Baseline differences between completers & non-completers | No significant differences.  “However, females (χ2(1) = 13.91, p<.001) and youth in control (χ2(1) = 39.31, p<.001) were more likely than their counterparts to complete the 3-month follow-up, and both variables are included in the models. The retention rate from T1 to T3 was 81%.” Pg. 5 |
| Attrition rates of randomised participants | 3-month FU Intervention: 28% 3-month FU Control: 8.3% \| Randomised \| Baseline survey and assent \| Intervention completion \| Immediate FU \| 3-month FU; TX \| 392 \| 349 \| 187 \| 258 \| 251; CO \| 321 \| 290 \| NA \| NA \| 266 |
| Limitations of study as described by authors | “The first set of limitations are related to the sample that was primarily white and non-Hispanic 4-H club members. The sample included only youth affiliated with 4-H as an organization, and thus may not represent other organizations or unaligned youth. It would be desirable to include longer delayed posttest evaluations to track effects. Additionally, there may be mediators or moderators that are unexplored or unmeasured in the project, though we tested many to rule out alternative explanations.” Pg. 6 |
| Funding source | Institutional, i.e. governmental, university \| Yes ☒ No ☐ Unclear ☐ Not Applicable ☐ Not Reported ☐; Private, i.e. pharmaceutical, software company \| Yes ☐ No ☒ Unclear ☐ Not Applicable ☐ Not Reported ☐ |
| Other aspects of study pertinent to online trial conception and execution | NA |

| Risk of Bias assessment  ([**https://handbook-5-1.cochrane.org/chapter_8/8_assessing_risk_of_bias_in_included_studies.htm**](https://handbook-5-1.cochrane.org/chapter_8/8_assessing_risk_of_bias_in_included_studies.htm)**)** | | | | |
| --- | --- | --- | --- | --- |
| Bias domain | Source of bias | Risk of bias | Support for judgment (use direct quotes where possible with explanatory comments) | Location in text or source (pg. number, figure, table etc.) |
| Selection bias | Random sequence generation | Low risk ☐    High risk ☐  Unclear risk ☒ | Quote: “After assent, youth were randomly assigned to treatment (n = 349,  55%) or delayed use control (n = 290, 45%) conditions.”  Comment: Insufficient information to assess risk. | Pg. 3, under Condition assignment and comparability |
|  | Allocation concealment | Low risk ☐    High risk ☐  Unclear risk ☒ | Quote: “The study methodologist made efforts to balance condition assignment by participant state, sex, race, and urban/rural location. To ensure comparability between the groups, initial analyses conducted on pretest survey variables of interest, including demographic variables and outcomes variables of interests (e.g., efficacy), indicated few differences between groups at Time 1 (p>.05).”  Comment: Insufficient information to assess risk. | Pg. 3, under Condition assignment and comparability |
| Performance bias | Blinding of participants and personnel | Low risk ☐    High risk ☒  Unclear risk ☐ | Comment: Blinding may have been compromised | Pg. 3, under Intervention: REAL media |
| Detection bias | Blinding of outcome assessment | Low risk ☐    High risk ☐  Unclear risk ☒ | Comment: There is insufficient information to make an assessment. | Not reported |
| Attrition bias | Incomplete outcome data | Low risk ☐    High risk ☒  Unclear risk ☐ | Comment: 3 month follow-up attrition rate was 36% | Pg. 4, Figure 2 |
| Reporting bias | Selective reporting | Low risk ☒    High risk ☐  Unclear risk ☐ | Comment: Study protocol is not available but the pre-specified outcomes listed in the clinical trials register have been reported and additional measures were reported in the study. | Registered at ClinicalTrials.gov: [NCT03157700](https://clinicaltrials.gov/ct2/show/NCT03157700?term=REAL+media&type=Intr&draw=2&rank=2) |
| Other bias | Anything else, ideally pre-specified | Low risk ☒    High risk ☐  Unclear risk ☐ | Comment: No other bias detected. | Not applicable |
