## Supplementary material for "Online randomised trials with children: A scoping review": S7 Appendix: Hillhouse.docx

| **Basic Study Characteristics** | |
| --- | --- |
| **Citation** | Hillhouse J, Turrisi R, Scaglione N, Cleveland M, Baker K, Florence L, et al. A Web-Based Intervention to Reduce Indoor Tanning Motivations in Adolescents: a Randomized Controlled Trial. Prevention Science. 2017;18(2):131-40. doi: 10.1007/s11121-016-0698-4. PubMed PMID: 120784944. Language: English. Entry Date: 20180723. Revision Date: 20191107. Publication Type: journal article. |
| **Characteristic reported (indicate with X for decision or enter free text, where applicable)** | |
| Type of study | Randomised trial ☒ \| Cluster randomised ☐; Quasi-randomised trial ☐ \| Individually randomised ☒ |
| Type of publication | Peer-reviewed journal ☒; Pre-print ☐; Other, if so, describe ☐ |
| Aim of study as per author’s description | To examine the efficacy of a web-based intervention to reduce tanning motivations in female adolescents. |
| **Participant Characteristics** | |
| Participant age demographics | Age range \| 12-18; Mean age \| 15.2 |
| Country(ies) of Participants’ Residence | U.S.A. |
| Number of participants randomised | Children 443; Caregivers Not reported |
| Gender demographics of participants | 100% female |
| Socioeconomic status of participants | Not reported |
| Race/Ethnicity of participants | From ClinicalTrial.gov: 12.2% Hispanic or Latino  87.8% Not Hispanic or Latino |
| Type of participant targeted | Females |
| Children’s advisory group formation | Ages of advisory group participants \| Not reported; Means of recruiting advisory group participants \| Not reported; Phases of trial involvement \| Pilot test Yes ☒ No ☐ Unclear ☐ Not Applicable ☐ Not Reported ☐ (Was an advisory group formed in the planning/execution of the study?) |
| Inclusion/ Exclusion Criteria | Included: Parent-teen dyads with female daughters with previous indoor tanning use or strong intentions or willingness to indoor tan in the future |
| **Methods of Recruitment, Retention & Consent** | |
| Online methods/platforms used for recruitment | Recruited from Knowledge Networks KnowledgePanel, a commercial online panel for measurement of public opinion, attitudes and behaviours, uses probability-based sampling with an address-based sampling methodology for selecting panel members. |
| Offline methods used for recruitment | NA |
| Methods/tools used for online consent/assent acquisition | Parental and teen consent obtained |
| Caregiver consent acquired/ waived (If waived, by what authority?) | Yes ☒ No ☐ Unclear ☐ Not Applicable ☐ Not Reported ☐  As above |
| Children assent acquisition | Yes ☒ No ☐ Unclear ☐ Not Applicable ☐ Not Reported ☐ |
| What, if any, methods were used to validate that assent/consent was informed? | Not reported  (E.g. validation questions, CAPTCHA, email confirmation, video-conferencing verbal) |
| Pushes/reminders sent to participants | Enrolment in trial Yes ☐ No ☐ Unclear ☐ Not Applicable ☐ Not Reported ☒; Retention/completion in trial Yes ☐ No ☐ Unclear ☐ Not Applicable ☐ Not Reported ☒; Follow-up in trial Yes ☐ No ☐ Unclear ☐ Not Applicable ☐ Not Reported ☒ |
| **Interventions** | |
| Number of arms | 2 |
| Intervention | Participants viewed a website designed to reduce indoor tanning motivations and increase sunless tanning willingness. |
| Comparison | Control participants viewed a teen-oriented alcohol prevention website. |
| Operating Systems/Devices required for participants’ engagement with trial | Not reported  (E.g. computer, phone, iOS, Windows, Android, iPhone, Facebook or Instagram account etc.) |
| Tools/software used for data protection processes | Not reported  (E.g. dedicated website, REDCap, institutional server etc.) |
| Tools/methods used for data collection | Individual survey links were sent to participating youth via email or text.  (E.g. data submitted via website, video-conferencing etc.) |
| Did caregivers participate/ contribute data to the trial? If so, describe. | Yes ☐ No ☒ Unclear ☐ Not Applicable ☐ Not Reported ☐ |
| Duration of intervention from first to final engagement | Unclear, but appears to be an SSI + 6-month follow-up survey  “Participants from two cohorts completed baseline assessments in May of 2011 and 2012.” Pg. 8 |
| Compensation offered to participants | Participants were offered up to $120 for completing all study requirements. |
| **Outcomes** | |
| Results relevant to this scoping review | NA  (E.g. satisfaction with online methods of trial, demographics of participants’ in relation to their outcomes especially in regarding online components, i.e., recruitment, retention, completion etc) |
| Baseline differences between completers & non-completers | “Analyses of baseline differences between participants who completed follow-up versus those who did not revealed no significant differences. Furthermore, examination of baseline differences in intervention participants who did not complete follow-up versus control participants who did not complete follow-up revealed no significant differences.” Pg. 8 |
| Attrition rates of randomised participants | Intervention: 214 dyads randomised, 182 completed 6-month FU: 15% attrition  Control: 229 dyads randomised, 206 completed 6-month FU: 10% attrition |
| Limitations of study as described by authors | The technology used (website) became quickly outdated (vs social media sites). The intervention only assessed the effects of one possible alternative, sunless tanning. |
| Funding source | Institutional, i.e. governmental, university \| Yes ☒ No ☐ Unclear ☐ Not Applicable ☐ Not Reported ☐; Private, i.e. pharmaceutical, software company \| Yes ☐ No ☒ Unclear ☐ Not Applicable ☐ Not Reported ☐ |
| Other aspects of study pertinent to online trial conception and execution | “A professional digital marketing company, Marketing Strategies, Inc., converted modules developed by the research team into a website, which was pretested with adolescents and then further refined using an interactive process involving the investigative team, the web development team and adolescent tanner beta testers.” Pg. 5 |

| Risk of Bias assessment  ([**https://handbook-5-1.cochrane.org/chapter_8/8_assessing_risk_of_bias_in_included_studies.htm**](https://handbook-5-1.cochrane.org/chapter_8/8_assessing_risk_of_bias_in_included_studies.htm)**)** | | | | |
| --- | --- | --- | --- | --- |
| Bias domain | Source of bias | Risk of bias | Support for judgment (use direct quotes where possible with explanatory comments) | Location in text or source (pg. number, figure, table etc.) |
| Selection bias | Random sequence generation | Low risk ☐    High risk ☐  Unclear risk ☒ | “The invitation was accepted by 443 dyads (77.6% participation rate) who were randomized into intervention (n = 214) and control (n = 229) groups.”  Comment: Insufficient information to assess risk. | Pg. 4, under Participants |
|  | Allocation concealment | Low risk ☐    High risk ☐  Unclear risk ☒ | Comment: Insufficient information to assess risk. | Not reported |
| Performance bias | Blinding of participants and personnel | Low risk ☒    High risk ☒  Unclear risk ☐ | Comment: Participants were not blinded as the outcome variables were willingness and intentions to indoor tan, willingness to sunless tan and measures of indoor tanning attitudes and beliefs. | Pg.1, under Abstract |
| Detection bias | Blinding of outcome assessment | Low risk ☐    High risk ☐  Unclear risk ☒ | Comment: There is insufficient information to make an assessment. | Not reported |
| Attrition bias | Incomplete outcome data | Low risk ☒    High risk ☐  Unclear risk ☐ | Comment: Intervention: 214 dyads randomised, 182 completed 6-month FU: 15% attrition  Control: 229 dyads randomised, 206 completed 6-month FU: 10% attrition | Pg. 16, Fig. 2 |
| Reporting bias | Selective reporting | Low risk ☒    High risk ☐  Unclear risk ☐ | Comment: Study protocol is not available but the pre-specified outcomes listed in the clinical trials register have been reported and additional measures were reported in the study. | Registered at ClinicalTrials.gov: [NCT01508013](https://clinicaltrials.gov/ct2/show/record/NCT01508013) |
| Other bias | Anything else, ideally pre-specified | Low risk ☒    High risk ☐  Unclear risk ☐ | Comment: No other bias detected | Not applicable |
