## Supplementary material for "Online randomised trials with children: A scoping review": S7 Appendix: Kelleher.docx

| **hiBasic Study Characteristics** | |
| --- | --- |
| **Citation** | Kelleher E, Moreno M, Wilt MP. Recruitment of Participants and Delivery of Online Mental Health Resources for Depressed Individuals Using Tumblr: Pilot Randomized Control Trial. JMIR Res Protoc. 2018;7(4):e95. doi: 10.2196/resprot.9421. PubMed PMID: 29650507. |
| **Characteristic reported (indicate with X for decision or enter free text, where applicable)** | |
| Type of study | Randomised trial ☒ \| Cluster randomised ☐; Quasi-randomised trial ☐ \| Individually randomised ☒ Pilot* |
| Type of publication | Peer-reviewed journal ☒; Pre-print ☐; Other, if so, describe ☐ |
| Aim of study as per author’s description | “To determine whether a social media intervention offering resources to young people displaying references to depression appropriately targeted young people with depression and was accessed by, and deemed acceptable by, young people.” Pg. 2 |
| **Participant Characteristics** | |
| Participant age demographics | Age range \| 15-23; Mean age \| 17.5 |
| Country(ies) of Participants’ Residence | U.S.A. |
| Number of participants randomised | Children 45; Caregivers Not reported |
| Gender demographics of participants | 65% female |
| Socioeconomic status of participants | Not reported |
| Race/Ethnicity of participants | Not reported |
| Type of participant targeted | Young people at risk for depression |
| Children’s advisory group formation | Ages of advisory group participants \| Not reported; Means of recruiting advisory group participants \| Not reported; Phases of trial involvement \| All participants were asked to provide suggestions on key components of online mental health interventions that would support their acceptability. Yes ☐ No ☒ Unclear ☐ Not Applicable ☐ Not Reported ☐ (Was an advisory group formed in the planning/execution of the study?) |
| Inclusion/ Exclusion Criteria | Included: met the diagnostic criteria for a depressive episode using a codebook to code the Tumblr posts in the last 14 days, Tumblr profile in English, ability to receive private Tumblr messages, and timestamps on profile posts.  Excluded: Individuals with Tumblr profiles who displayed other mental health comorbidities (ie, #bipolar) |
| **Methods of Recruitment, Retention & Consent** | |
| Online methods/platforms used for recruitment | “Potential participants were identified via a post on Tumblr using the search term #depress to encompass both ‘depression’ and ‘depressed’ and ‘most recent posts’ was used as a filter. Profiles of users were reviewed and a private Tumblr message was sent explaining the study and the research team asking possible participants to message the group via Tumblr if they did not wish to be contacted again.” Pg. 2 |
| Offline methods used for recruitment | NA |
| Methods/tools used for online consent/assent acquisition | One month post intervention a private Tumblr message was sent to both groups with a secure, anonymous online survey link to consent. |
| Caregiver consent acquired/ waived (If waived, by what authority?) | Yes ☐ No ☒ Unclear ☐ Not Applicable ☐ Not Reported ☐  No caregiver consent reported. The Western Institutional Review Board approved the pilot study. |
| Children assent acquisition | Yes ☒ No ☐ Unclear ☐ Not Applicable ☐ Not Reported ☐ |
| What, if any, methods were used to validate that assent/consent was informed? | Not reported  (E.g. validation questions, CAPTCHA, email confirmation, video-conferencing verbal) |
| Pushes/reminders sent to participants | Enrolment in trial Yes ☐ No ☒ Unclear ☐ Not Applicable ☐ Not Reported ☐; Retention/completion in trial Yes ☐ No ☒ Unclear ☐ Not Applicable ☐ Not Reported ☒; Follow-up in trial Yes ☐ No ☒ Unclear ☐ Not Applicable ☐ Not Reported ☒ |
| **Interventions** | |
| Number of arms | 2 |
| Intervention | A resource sheet with a list of mental health resources that were nationally available, free, online, and publicly accessible were sent using a study team Tumblr profile via a private Tumblr message explaining the study and the research team. After one month, a private Tumblr message was sent to the intervention group to link to the secure, anonymous online survey and consent form. Pg. 3 |
| Comparison | “the control group were not initially contacted. After one month a private Tumblr message was sent to the control group to link to the secure, anonymous online survey and consent form.” Pg. 3 |
| Operating Systems/Devices required for participants’ engagement with trial | Tumblr only  (E.g. computer, phone, iOS, Windows, Android, iPhone, Facebook or Instagram account etc.) |
| Tools/software used for data protection processes | Tumblr allows creation and display of profiles anonymously and an email address and username is the only information needed for an account. “Tumblr profiles were identified, randomized, the username, profile URL, post with the depression reference, and hashtags of participants were stored in an excel spreadsheet.” Pg. 3  (E.g. dedicated website, REDCap, institutional server etc.) |
| Tools/methods used for data collection | Tumblr message sent to both groups with a link to a secure, anonymous online survey and consent form.  (E.g. data submitted via website, video-conferencing etc.) |
| Did caregivers participate/ contribute data to the trial? If so, describe. | Yes ☐ No ☒ Unclear ☐ Not Applicable ☐ Not Reported ☐ |
| Duration of intervention from first to final engagement | SSI +1-month follow-up |
| Compensation offered to participants | Participants who completed the survey were provided a $10 gift card |
| **Outcomes** | |
| Outcomes relevant to this review | NA  (E.g. satisfaction with online methods of trial, demographics of participants’ in relation to their outcomes especially in regarding online components, i.e., recruitment, retention, completion etc) |
| Baseline differences between completers & non-completers | Not reported |
| Attrition rates of randomised participants | 45 profiles randomised: 21 to intervention, 24 to control  11 completed survey post-intervention: 47.6%  14 completed survey post-control: 41.7% |
| Limitations of study as described by authors | “Only one social media site was used and the sample size was small, therefore findings cannot be generalized. Due to profile deletion and nonresponse to the survey, the intervention group had only female participants. Due to the anonymity of the surveys and the priority to protect participant identities, researchers had no capacity to follow up if participants answered that they had thoughts self-harm or suicide. The anonymity of the survey also prevented researchers from contacting participants to remind them to take the survey.” Pg. 5 |
| Funding source | Institutional, i.e. governmental, university \| Yes ☒ No ☐ Unclear ☐ Not Applicable ☐ Not Reported ☐; Private, i.e. pharmaceutical, software company \| Yes ☐ No ☒ Unclear ☐ Not Applicable ☐ Not Reported ☐ |
| Other aspects of study pertinent to online trial conception and execution | IP addresses were verified to prevent duplicate entries from the same user |

| Risk of Bias assessment  ([**https://handbook-5-1.cochrane.org/chapter_8/8_assessing_risk_of_bias_in_included_studies.htm**](https://handbook-5-1.cochrane.org/chapter_8/8_assessing_risk_of_bias_in_included_studies.htm)**)** | | | | |
| --- | --- | --- | --- | --- |
| Bias domain | Source of bias | Risk of bias | Support for judgment (use direct quotes where possible with explanatory comments) | Location in text or source (pg. number, figure, table etc.) |
| Selection bias | Random sequence generation | Low risk ☒    High risk ☐  Unclear risk ☐ | “After eligible individuals’ Tumblr profiles were identified and randomized, the username, profile URL, post with the depression symptom reference, and hashtags of participants were stored in an excel spreadsheet. Participants were randomized using a randomization website [27]. The numbers 1 through 45 were randomized into intervention and control numbers. Participants were assigned numbers in the order their post appeared on Tumblr.”  Comment: Randomisation website is Random.org. | Pg. 3, under Procedures |
|  | Allocation concealment | Low risk ☒    High risk ☐  Unclear risk ☐ | Comment: As above. Neither participants, nor researchers were aware group assignment prior to allocation. | Pg. 3, under Procedures |
| Performance bias | Blinding of participants and personnel | Low risk ☐    High risk ☒  Unclear risk ☐ | “There was no blinding for this study.”  Comment: It is likely that lack of blinding would affect the outcomes of this study. | Pg. 4, under Analysis |
| Detection bias | Blinding of outcome assessment | Low risk ☐    High risk ☐  Unclear risk ☒ | Comment: There is no mention of assessor blinding and there is insufficient information to make an assessment. | Not reported |
| Attrition bias | Incomplete outcome data | Low risk ☐    High risk ☒  Unclear risk ☐ | Comment: 45 profiles randomised: 21 to intervention, 24 to control  11 completed survey post-intervention: 47.6%  14 completed survey post-control: 41.7% | Pg. 5, under Limitations |
| Reporting bias | Selective reporting | Low risk ☐    High risk ☐  Unclear risk ☒ | Comment: Study protocol is not available and there is insufficient information to permit assessment. | Not reported |
| Other bias | Anything else, ideally pre-specified | Low risk ☐    High risk ☒  Unclear risk ☐ | “Due to profile deletion and nonresponse to the survey, the intervention group had only female participants. “  Comment: The study has a potential source of bias related to its study design. The number of participants is very low and this led to unbalanced gender distribution in the intervention vs control arms. | Pg. 5, under Limitations |
