## Supplementary material for "Online randomised trials with children: A scoping review": S7 Appendix: Lester.docx

| **Basic Study Characteristics** | |
| --- | --- |
| **Citation** | Lester E, DiStefano S, Mace R, Macklin E, Plotkin S, Vranceanu AM. Virtual mind-body treatment for geographically diverse youth with neurofibromatosis: A pilot randomized controlled trial. Gen Hosp Psychiatry. 202 Jan-Feb; 62:72-78. Doi: 10.1016/j.genhosppsych.2019.12.001. |
| **Characteristic reported (indicate with X for decision or enter free text, where applicable)** | |
| Type of study | Randomised trial ☒ \| Cluster randomised ☐; Quasi-randomised trial ☐ \| Individually randomised ☒ Pilot* |
| Type of publication | Peer-reviewed journal ☒; Pre-print ☐; Other, if so, describe ☐ |
| Aim of study as per author’s description | “To examine the feasibility, acceptability, preliminary effect, and durability of a mind-body video-conferencing program for youth with neurofibromatosis against an experimental educational control.” Pg. 72 |
| **Participant Characteristics** | |
| Participant age demographics | Age range \| 12-17; Mean age \| 14.37 |
| Country(ies) of Participants’ Residence | 42 from U.S.A., 8 from Canada, 1 from Columbia |
| Number of participants randomised | Children 51; Caregivers Not reported |
| Gender demographics of participants | 41% female; 59% male |
| Socioeconomic status of participants | Not reported |
| Race/Ethnicity of participants | 76.47% White  13.73% other  11.76% Hispanic of Latino |
| Type of participant targeted | Youth with neurofibromatosis |
| Children’s advisory group formation | Ages of advisory group participants \| Not reported; Means of recruiting advisory group participants \| Not reported; Phases of trial involvement \| The intervention was adapted for adolescents with NF from another intervention 3RP program (for adults) through an interactive process that included qualitative interviews, an open pilot study, and exit interviews. Yes ☒ No ☐ Unclear ☐ Not Applicable ☐ Not Reported ☐ (Was an advisory group formed in the planning/execution of the study?) |
| Inclusion/ Exclusion Criteria | Included: NF1 or NF2 diagnosis, able to provide informed assent/consent, English comprehension at a third-grade level, and parent and/or adolescent endorsed stress and difficulties coping with NF symptoms |
| **Methods of Recruitment, Retention & Consent** | |
| Online methods/platforms used for recruitment | Study flyer was emailed through the NF registry of the Children’s Tumor Foundation and distributed by clinicians in the NF clinic. Interested parents emailed the team and an intake visit via Skype was conducted. |
| Offline methods used for recruitment | NA |
| Methods/tools used for online consent/assent acquisition | Skype intake session |
| Caregiver consent acquired/ waived (If waived, by what authority?) | Yes ☒ No ☐ Unclear ☐ Not Applicable ☐ Not Reported ☐ |
| Children assent acquisition | Yes ☒ No ☐ Unclear ☐ Not Applicable ☐ Not Reported ☐ |
| What, if any, methods were used to validate that assent/consent was informed? | Over Skype  (E.g. validation questions, CAPTCHA, email confirmation, video-conferencing verbal) |
| Pushes/reminders sent to participants | Enrolment in trial Yes ☐ No ☒ Unclear ☐ Not Applicable ☐ Not Reported ☐; Retention/completion in trial Yes ☒ No ☐ Unclear ☐ Not Applicable ☐ Not Reported ☐; Follow-up in trial Yes ☒ No ☐ Unclear ☐ Not Applicable ☐ Not Reported ☐ Each participant received three reminder e-mails to complete study questionnaires before deemed lost to follow-up. |
| **Interventions** | |
| Number of arms | 2 |
| Intervention | All participants received an emailed patient manual. Intervention participants received mindfulness recording exercises (5-10 mins long) tailored to NF and adolescent specific concerns. Eight group sessions (45 mins each) via Skype, led by an experienced clinical psychologist. Each group capped at 5 participants. |
| Comparison | All participants received an emailed patient manual. Provided accessible educational information on stress, NF, and lifestyle behaviours. No resiliency skills taught. Eight group sessions (45 mins each) via Skype, led by an experienced clinical psychologist. Each group capped at 5 participants. |
| Operating Systems/Devices required for participants’ engagement with trial | Skype only for sessions; audio files were downloaded on phones or computers by participants  (E.g. computer, phone, iOS, Windows, Android, iPhone, Facebook or Instagram account etc.) |
| Tools/software used for data protection processes | REDCap  (E.g. dedicated website, REDCap, institutional server etc.) |
| Tools/methods used for data collection | REDCap  (E.g. data submitted via website, video-conferencing etc.) |
| Did caregivers participate/ contribute data to the trial? If so, describe. | Yes ☐ No ☒ Unclear ☐ Not Applicable ☐ Not Reported ☐ |
| Duration of intervention from first to final engagement | 8 weeks +6-month follow-up |
| Compensation offered to participants | Not reported |
| **Outcomes** | |
| Results relevant to this scoping review | A single item on a 5-point Likert scale measured satisfaction.  (E.g. satisfaction with online methods of trial, demographics of participants’ in relation to their outcomes especially in regarding online components, i.e., recruitment, retention, completion etc)  Intervention: 4.08/5  Control: 3.49/5 |
| Baseline differences between completers & non-completers | No differences in demographics or study outcomes between completers and non-completers were detected. |
| Attrition rates of randomised participants | Completion was determined if at least 6 sessions were attended. 51 randomised \| Randomised \| Immediate post-test complete \| % attrition \| 6-month FU complete \| % attrition; Intervention \| 27 \| 24 \| 11.11 \| 21 \| 22.2; Control \| 24 \| 21 \| 21.5 \| 18 \| 25 |
| Limitations of study as described by authors | Small sample size, short FU period (6 months). Other study limitations included the use of one treating clinician, not tracking the number of sessions attended or missed by each participant, the use of Skype versus a HIPAA compliant program (e.g., Vidyo software), and using a mean substitution versus a multiple imputation method for missing data. |
| Funding source | Institutional, i.e. governmental, university \| Yes ☒ No ☐ Unclear ☐ Not Applicable ☐ Not Reported ☐; Private, i.e. pharmaceutical, software company \| Yes ☐ No ☒ Unclear ☐ Not Applicable ☐ Not Reported ☐ |
| Other aspects of study pertinent to online trial conception and execution | Protocols in place in case participants presented with safety risks, participants were briefed in first session on group rules. The strengths of the adolescent program which may have led to the high retention rates were the focus on engaging youth by teaching skills through games and exercises, meeting participants at their own developmental and cognitive level, and addressing topics that are important to them in a fun and engaging manner |

| Risk of Bias assessment  ([**https://handbook-5-1.cochrane.org/chapter_8/8_assessing_risk_of_bias_in_included_studies.htm**](https://handbook-5-1.cochrane.org/chapter_8/8_assessing_risk_of_bias_in_included_studies.htm)**)** | | | | |
| --- | --- | --- | --- | --- |
| Bias domain | Source of bias | Risk of bias | Support for judgment (use direct quotes where possible with explanatory comments) | Location in text or source (pg. number, figure, table etc.) |
| Selection bias | Random sequence generation | Low risk ☒    High risk ☐  Unclear risk ☐ | “A free internet-based randomization program (randomization.com) was used to assign participants to conditions using a 1:1 ratio within blocks of ten: 5 to RY-NF and 5 to HE-NF.” | Pg. 73, under Randomization |
|  | Allocation concealment | Low risk ☐    High risk ☐  Unclear risk ☒ | Comment: Based on the randomisation process described, the researchers were most likely unaware of allocation to arm but it is unclear. | Not reported |
| Performance bias | Blinding of participants and personnel | Low risk ☐    High risk ☒  Unclear risk ☐ | “despite participants being masked (i.e., “participation in one of two stress management interventions”) and being given a credible experimental educational control (satisfaction was high and comparable) [11], it is still possible that some participants could have guessed their treatment condition.” | Pg. 76, under Discussion |
| Detection bias | Blinding of outcome assessment | Low risk ☐    High risk ☒  Unclear risk ☐ | Comment: Study notes it was ‘single-blind’ Does not state clearly what this means but does state efforts to blind participants. Therefore, assessors were most likely not blinded to participant allocation. | Not reported |
| Attrition bias | Incomplete outcome data | Low risk ☒    High risk ☐  Unclear risk ☐ | Comment: 45 profiles randomised: 21 to intervention, 24 to control  11 completed survey post-intervention: 47.6%  14 completed survey post-control: 41.7% | Pg. 74, Figure 1 |
| Reporting bias | Selective reporting | Low risk ☐    High risk ☐  Unclear risk ☒ | Comment: Study protocol is not available and there is insufficient information to permit assessment. | Not reported |
| Other bias | Anything else, ideally pre-specified | Low risk ☒    High risk ☐  Unclear risk ☐ | Comment: No other bias detected | Not applicable |
