## Supplementary material for "Online randomised trials with children: A scoping review": S7 Appendix: Manicavasagar.docx

| **Basic Study Characteristics** | |
| --- | --- |
| **Citation** | Manicavasagar V, Horswood D, Burckhardt R, Lum A, Hadzi-Pavlovic D, Parker G. Feasibility and effectiveness of a web-based positive psychology program for youth mental health: randomized controlled trial. Journal of medical Internet research. 2014;16(6):e140. doi: 10.2196/jmir.3176. PubMed PMID: CN-01116094. |
| **Characteristic reported (indicate with X for decision or enter free text, where applicable)** | |
| Type of study | Randomised trial ☒ \| Cluster randomised ☐; Quasi-randomised trial ☐ \| Individually randomised ☒ |
| Type of publication | Peer-reviewed journal ☒; Pre-print ☐; Other, if so, describe ☐ |
| Aim of study as per author’s description | Explore the feasibility of the online delivery of a youth positive psychology program to improve the well-being and mental health outcomes of Australian youth. |
| **Participant Characteristics** | |
| Participant age demographics | Age range \| 12-18; Mean age \| 15.4 |
| Country(ies) of Participants’ Residence | Australia |
| Number of participants randomised | Children 235; Caregivers Not reported |
| Gender demographics of participants | 67.5% female |
| Socioeconomic status of participants | Not reported |
| Race/Ethnicity of participants | Not reported |
| Type of participant targeted | NA |
| Children’s advisory group formation | Ages of advisory group participants \| Not reported; Means of recruiting advisory group participants \| Not reported; Phases of trial involvement \| Not reported Yes ☐ No ☐ Unclear ☐ Not Applicable ☐ Not Reported ☒ (Was an advisory group formed in the planning/execution of the study?) |
| Inclusion/ Exclusion Criteria | Included: resident in Australia, valid email address, access to a computer with an internet connection, providing parental consent if under 16 years old.  Excluded: duplicate email addresses resulted in the first application being retained and duplicate discarded. |
| **Methods of Recruitment, Retention & Consent** | |
| Online methods/platforms used for recruitment | Recruitment was through schools and youth organizations across Australia. Promotional info packs advertising the How Do You View the World Study were disseminated via mail and email and included a letter to the organization, principal, and/or school counsellor. Organizations were asked to distribute flyers promoting the study to young people in any manner they deemed appropriate: newsletters, websites, announcements. |
| Offline methods used for recruitment | As above, mixed. |
| Methods/tools used for online consent/assent acquisition | Interested adolescents provided their email address, name, date of birth, sex, postcode, and parental contact email if younger than 16. If eligible, they were emailed a link to the baseline questionnaires. |
| Caregiver consent acquired/ waived (If waived, by what authority?) | Yes ☒ No ☐ Unclear ☐ Not Applicable ☐ Not Reported ☐  If under 16, caregiver consent was required. |
| Children assent acquisition | Yes ☒ No ☐ Unclear ☐ Not Applicable ☐ Not Reported ☐ |
| What, if any, methods were used to validate that assent/consent was informed? | Not reported  (E.g. validation questions, CAPTCHA, email confirmation, video-conferencing verbal) |
| Pushes/reminders sent to participants | Enrolment in trial Yes ☐ No ☒ Unclear ☐ Not Applicable ☐ Not Reported ☐; Retention/completion in trial Yes ☒ No ☐ Unclear ☐ Not Applicable ☐ Not Reported ☐; Follow-up in trial Yes ☒ No ☐ Unclear ☐ Not Applicable ☐ Not Reported ☐ Both Bite Back and control participants received reminder emails once a week to encourage use and engagement. Six weeks from start date, participants were emailed postintervention questionnaire. |
| **Interventions** | |
| Number of arms | 2 |
| Intervention | The Bite Back website uses positive psychology via a combination of interactive exercises and information across 9 positive psychology domains and also provides information about the benefits of increasing well-being, methods of skill development, links to other resources, and allows for comments and online discussions. |
| Comparison | Control websites were: ABC3 digital channel website, and Nine MSN’s entertainment website, The Fix, which would engage young people and were similar to Bite Back in that they are multicomponent, self-guided, youth-oriented, and Australian-based, with options to contribute personal work, opinions, and stories. |
| Operating Systems/Devices required for participants’ engagement with trial | Any device with internet access  (E.g. computer, phone, iOS, Windows, Android, iPhone, Facebook or Instagram account etc.) |
| Tools/software used for data protection processes | Bite Back (dedicated study website)  (E.g. dedicated website, REDCap, institutional server etc.) |
| Tools/methods used for data collection | Bite Back (dedicated study website)  (E.g. data submitted via website, video-conferencing etc.) |
| Did caregivers participate/ contribute data to the trial? If so, describe. | Yes ☐ No ☒ Unclear ☐ Not Applicable ☐ Not Reported ☐ |
| Duration of intervention from first to final engagement | 6 weeks |
| Compensation offered to participants | Voucher for a media outlet worth AU$20 for completion of study |
| **Outcomes** | |
| Results relevant to this scoping review | Acceptability of the Bite Back website was determined using 7 point Likert scale. 49/62 participants who used the program for 6 weeks reported it was fun and 52/62 rated activities as interesting, 56/62 said the site was easy to use. 57% said they would revisit the website.  (E.g. satisfaction with online methods of trial, demographics of participants’ in relation to their outcomes especially in regarding online components, i.e., recruitment, retention, completion etc)  “Although a larger percentage of those younger than 16 year expressed interest in participating, fewer of the participants younger than 16 years progressed through to actually participate in the study because of the need for them to obtain parental permission.” Pg.14 |
| Baseline differences between completers & non-completers | “Completers and drop-outs were equivalent in demographics, the SWEMWBS, and the depression and anxiety subscales of the DASS-21, but drop-outs reported significantly higher levels of stress than completers.” Pg. 1 |
| Attrition rates of randomised participants | Intervention: 45/120: 37.5% attrition  Control: 23/115: 20% |
| Limitations of study as described by authors | Measurement tools, small sample size, self-reported data, limited information gathered about participants’ use of Bite Back, required parental consent for <16s limited their involvement. |
| Funding source | Institutional, i.e. governmental, university \| Yes ☒ No ☐ Unclear ☐ Not Applicable ☐ Not Reported ☐; Private, i.e. pharmaceutical, software company \| Yes ☐ No ☒ Unclear ☐ Not Applicable ☐ Not Reported ☐ |
| Other aspects of study pertinent to online trial conception and execution | Feedback from some participants on why they spent less than an hour/week on the site (as was recommended): “The website was very similar each time I visited it and thus lost the initial flair it once had”  The website was pre-moderated with each comment and upload being monitored and approved before being published. |

| Risk of Bias assessment  ([**https://handbook-5-1.cochrane.org/chapter_8/8_assessing_risk_of_bias_in_included_studies.htm**](https://handbook-5-1.cochrane.org/chapter_8/8_assessing_risk_of_bias_in_included_studies.htm)**)** | | | | |
| --- | --- | --- | --- | --- |
| Bias domain | Source of bias | Risk of bias | Support for judgment (use direct quotes where possible with explanatory comments) | Location in text or source (pg. number, figure, table etc.) |
| Selection bias | Random sequence generation | Low risk ☒    High risk ☐  Unclear risk ☐ | Quote: “Participants who completed the baseline questionnaires were randomly allocated to one of two conditions through a block randomization method. An independent researcher not associated with this study used a random number generator in Excel to allocate blocks of 10 participants to one of two conditions.” | Pg. 8, under Study Procedures |
|  | Allocation concealment | Low risk ☒    High risk ☐  Unclear risk ☐ | Comment: As described above.  Quote: ‘Following baseline assessment, an email was sent to participants that included a link to their allocated website and instructions on how to use it however and whenever they wanted over the next 6 weeks, but “for at least an hour a week.” Participants could access their allocated website from any Internet-enabled device and from any location.’ | Pg. 8, under Study Procedures |
| Performance bias | Blinding of participants and personnel | Low risk ☐    High risk ☒  Unclear risk ☐ | “The study was advertised as the “How Do You View the World Study: an investigation into how websites impact on the way young people think, react, and interact with the world.” It was important to conceal the clinical focus of this study and participants’ allocated condition for 2 reasons: (1) to ensure that control participants did not use the Bite Back website, and (2) to minimize any expectancy effects.”  Comment: It is possible that participants were aware of their allocation based on content of website. | Pg. 7, under Study Procedures |
| Detection bias | Blinding of outcome assessment | Low risk ☐    High risk ☒  Unclear risk ☐ | “The researchers who conducted the analyses were not blinded to the allocated condition of participants.” | Pg. 9, under Results, Attrition and Sample Characteriscs |
| Attrition bias | Incomplete outcome data | Low risk ☐    High risk ☒  Unclear risk ☐ | Intervention: 45/120: 37.5% attrition  Control: 23/115: 20% | Pg. 9, Fig. 4 |
| Reporting bias | Selective reporting | Low risk ☐    High risk ☒  Unclear risk ☐ | Comment: Study protocol is not available but the trial is registered at Australian New Zealand Clinical Trials Registry. Both the SWEMBS and DASS-21 score are reported in the study as outlined in the register; however, the registry also lists the Student Life Satisfaction Scale (SLSS) the Scale of Positive and Negative Experience (SPANE), modified Life Orientation Test-Revised (LOT-R), the General Self-Efficacy Scale (GSE), and modified Rosenberg’s Self-Esteem scale are all listed and not reported.  Comment: It is possible these outcomes will be assessed in the full-scale trial. | Australian New Zealand Clinical Trials Registry: ACTRN1261200057831; |
| Other bias | Anything else, ideally pre-specified | Low risk ☐    High risk ☒  Unclear risk ☐ | “Our study may have been affected by measurement sensitivity (ie, the psychopathology measure selected was designed for a clinical population and so floor effects could have impacted on our results). This problem may have been exacerbated by the small sample sizes in each condition making it less likely to obtain significant differences.”  “In addition, adolescent samples may be likely to underreport mental health symptoms, although the anonymous nature of this study may have lessened this likelihood.” | Pg. 14, under Limitations of the Study |
