## Supplementary material for "Online randomised trials with children: A scoping review": S7 Appendix: Mogil.docx

| **Basic Study Characteristics** | |
| --- | --- |
| **Citation** | Mogil C, Hajal N, Aralis H, Paley B, Milburn NG, Barrera W, et al. A Trauma-Informed, Family-Centered, Virtual Home Visiting Program for Young Children: one-Year Outcomes. Child psychiatry and human development [Internet]. 2021; Available from: <https://www.cochranelibrary.com/central/doi/10.1002/central/CN-02273506/full> |
| **Characteristic reported (indicate with X for decision or enter free text, where applicable)** | |
| Type of study | Randomised trial ☒ \| Cluster randomised ☐; Quasi-randomised trial ☐ \| Individually randomised ☒ |
| Type of publication | Peer-reviewed journal ☒; Pre-print ☐; Other, if so, describe ☐ |
| Aim of study as per author’s description | Evaluate the efficacy of Families OverComing Under Stress-Early Childhood (FOCUS-EC) delivered through an in-home, virtual telehealth platform. FOCUS-EC is a trauma-informed, family-centered preventive intervention designed to promote family resilience and well-being. Pg. 1 |
| **Participant Characteristics** | |
| Participant age demographics | Age range \| Caregivers and their 3- to 6-year old children; Mean age \| “53.7 months” (4.5 years) |
| Country(ies) of Participants’ Residence | U.S.A. |
| Number of participants randomised | Children 200; Caregivers 200 families, only one caregiver was needed to participate |
| Gender demographics of participants | 51.2% female children |
| Socioeconomic status of participants | Income:   $39,999 or less: 20.3%    $40,000 to $59,999: 25.4%  $60,000 or more: 54.3% |
| Race/Ethnicity of participants | Maternal Race  American Indian or Alaska Native 1  Asian 16  Black or African American 15  Native Hawaiian or Other Pacifc Islander 3  White 124  Other 16  More than one race 11  Paternal Race Asian 6  Black or African American 13  Native Hawaiian or Other Pacifc Islander 2  White 97  Other 15  More than one race 17  Maternal Ethnicity  Hispanic, Latino or Spanish origin 74  Not of Hispanic, Latino or Spanish origin 117  Paternal Ethnicity  Hispanic, Latino or Spanish origin 58  Not of Hispanic, Latino or Spanish origin 92 |
| Type of participant targeted | Military connected families (MCF) with 3- to 6-year old children |
| Children’s advisory group formation | Ages of advisory group participants \| Not reported; Means of recruiting advisory group participants \| Not reported; Phases of trial involvement \| Not reported Yes ☐ No ☐ Unclear ☐ Not Applicable ☐ Not Reported ☒ (Was an advisory group formed in the planning/execution of the study?) |
| Inclusion/ Exclusion Criteria | Included: at least one parent of the 3- to 6- year old serving post 9/11 in U.S. Army, Navy, Marine Corps or Air Force |
| **Methods of Recruitment, Retention & Consent** | |
| Online methods/platforms used for recruitment | Targeted social media advertising |
| Methods/tools used for online consent/assent acquisition | Video-conferencing platform: not named. |
| Caregiver consent acquired/ waived (If waived, by what authority?) | Yes ☒ No ☐ Unclear ☐ Not Applicable ☐ Not Reported ☐  “Informed consent was obtained from all individual participants included in the study.” |
| Children assent acquisition | Yes ☐ No ☐ Unclear ☒ Not Applicable ☐ Not Reported ☐ |
| What, if any, methods were used to validate that assent/consent was informed? | Video-conferencing platform: not named.  (E.g. validation questions, CAPTCHA, email confirmation, video-conferencing verbal) |
| Pushes/reminders sent to participants | Enrolment in trial Yes ☐ No ☐ Unclear ☐ Not Applicable ☐ Not Reported ☒; Retention/completion in trial Yes ☐ No ☐ Unclear ☐ Not Applicable ☐ Not Reported ☒; Follow-up in trial Yes ☐ No ☐ Unclear ☐ Not Applicable ☐ Not Reported ☒ |
| **Interventions** | |
| Number of arms | 2 |
| Intervention | “FOCUS-EC consists of core elements delivered in 6 modules that are typically delivered over 4–10 meetings that last 60–90 min. Consistent with the FOCUS model described previously [34, 35], the core elements include (1) webbased Family Resilience Check-In (FRCI); (2) personalized trauma-informed psychoeducation, parenting education and developmental guidance; (3) development of a parental narrative timeline to support refection, empathy, meaning making and communication; and (4) development of family resilience and parenting/co-parenting skills.” Pg. 3 |
| Comparison | The active control condition included access to online parent education (OPE). |
| Operating Systems/Devices required for participants’ engagement with trial | Any device with internet access  (E.g. computer, phone, iOS, Windows, Android, iPhone, Facebook or Instagram account etc.) |
| Tools/software used for data protection processes | Video-conferencing  (E.g. dedicated website, REDCap, institutional server etc.) |
| Tools/methods used for data collection | Video-conferencing  (E.g. data submitted via website, video-conferencing etc.) |
| Did caregivers participate/ contribute data to the trial? If so, describe. | Yes ☒ No ☐ Unclear ☐ Not Applicable ☐ Not Reported ☐ |
| Duration of intervention from first to final engagement | 4-10 weeks + 3-, 6-, and 12-months follow-ups |
| Compensation offered to participants | Not reported |
| **Outcomes** | |
| Results relevant to this scoping review | “Among the 91% (n=477) of visits for which family member participation was recorded, only 194 sessions (41%) were attended by two parents; of the 283 sessions attended by only one parent, the vast majority (88%, n=249) were attended by a mother only.” Pg. 12  (E.g. satisfaction with online methods of trial, demographics of participants’ in relation to their outcomes especially in regarding online components, i.e., recruitment, retention, completion etc) |
| Baseline differences between completers & non-completers | Not reported |
| Attrition rates of randomised participants | \| Randomised \| Baseline completed \| 3-month FU \| 6-month FU \| 12-month FU \| % attrition at 12 month FU; Intervention \| 100 \| 100 \| 86 \| 88 \| 86 \| 14; Control \| 100 \| 100 \| 92 \| 91 \| 94 \| 6 N are numbers of families |
| Limitations of study as described by authors | Small sample size, heterogeneous sample may have limited characterization of sample and intervention effects and generalizability. |
| Funding source | Institutional, i.e. governmental, university \| Yes ☒ No ☐ Unclear ☐ Not Applicable ☐ Not Reported ☐; Private, i.e. pharmaceutical, software company \| Yes ☐ No ☒ Unclear ☐ Not Applicable ☐ Not Reported ☐ |
| Other aspects of study pertinent to online trial conception and execution | NA |

| Risk of Bias assessment  ([**https://handbook-5-1.cochrane.org/chapter_8/8_assessing_risk_of_bias_in_included_studies.htm**](https://handbook-5-1.cochrane.org/chapter_8/8_assessing_risk_of_bias_in_included_studies.htm)**)** | | | | |
| --- | --- | --- | --- | --- |
| Bias domain | Source of bias | Risk of bias | Support for judgment (use direct quotes where possible with explanatory comments) | Location in text or source (pg. number, figure, table etc.) |
| Selection bias | Random sequence generation | Low risk ☐    High risk ☐  Unclear risk ☒ | “Among the remaining 230 families, 30 declined to participate and 200 enrolled and were randomly assigned with equal allocation across the intervention group (FOCUS-EC) and control condition (OPE).”  Comment: There is not sufficient information to assess this risk. | Pg. 4, under Study Sample |
|  | Allocation concealment | Low risk ☐    High risk ☐  Unclear risk ☒ | Comment: Insufficient evidence to assess. According to Clinical Trial registry: “Allocation: Randomized” | Not reported |
| Performance bias | Blinding of participants and personnel | Low risk ☐    High risk ☒  Unclear risk ☐ | Comment: There is insufficient evidence to assess this risk. | Not reported |
| Detection bias | Blinding of outcome assessment | Low risk ☒    High risk ☐  Unclear risk ☐ | Outcome (Parent-Child Relationships):  Quote: “Measurement of parent–child relationships consisted of both parent-reported and observational measures.”  Quote: “The last 5 min of each videotaped task were coded by undergraduate students who were blind to information regarding randomization status and assessment time point, under the guidance of a postdoctoral fellow and the project director.”  Comment: According to Clinical Trial registry “Masking: Double (Investigator, Outcomes Assessor)” | Pg. 4, under Parent-Child Relationships  Pg. 8, under Parent-Child Relationships  ClinicalTrials.gov: NCT04598100 |
| Attrition bias | Incomplete outcome data | Low risk ☒    High risk ☐  Unclear risk ☐ | Comment:  Intervention attrition 14%  Control attrition 6% | Pg. 5, Figure 2 |
| Reporting bias | Selective reporting | Low risk ☒    High risk ☐  Unclear risk ☐ | Comment: Study protocol is not available but the trial is registered at ClinicalTrials.gov and all pre-specified outcomes seem to be reported. | ClinicalTrials.gov: NCT04598100 |
| Other bias | Anything else, ideally pre-specified | Low risk ☒    High risk ☐  Unclear risk ☐ | Comment: No other bias detected. | Not applicable |
