## Supplementary material for "Online randomised trials with children: A scoping review": S7 Appendix: Moreno.docx

| **Basic Study Characteristics** | |
| --- | --- |
| **Citation** | Moreno MA, Binger KS, Zhao Q, Eickhoff JC. Effect of a Family Media Use Plan on Media Rule Engagement Among Adolescents. JAMA Pediatrics. 2021;175(4):351. |
| **Characteristic reported (indicate with X for decision or enter free text, where applicable)** | |
| Type of study | Randomised trial ☒ \| Cluster randomised ☐; Quasi-randomised trial ☐ \| Individually randomised ☒ |
| Type of publication | Peer-reviewed journal ☒; Pre-print ☐; Other, if so, describe ☐ |
| Aim of study as per author’s description | “To test the effect of a family media use plan on media rule engagement in adolescents.” Pg. 352 |
| **Participant Characteristics** | |
| Participant age demographics | Age range \| 12-17; Mean age \| 14.5 |
| Country(ies) of Participants’ Residence | U.S.A. |
| Number of participants randomised | Children/ participants: 1520 parent-adolescent dyads; Caregivers at least 1520 |
| Gender demographics of participants | 51.9% female |
| Socioeconomic status of participants | 53.9% of parents reported their family income as being below the national median income |
| Race/Ethnicity of participants | 14.67% Black  9.27% Latino  67.56% White  8.55% Other |
| Type of participant targeted | No group targeted |
| Children’s advisory group formation | Ages of advisory group participants \| Not reported; Means of recruiting advisory group participants \| Not reported; Phases of trial involvement \| Not reported Yes ☐ No ☐ Unclear ☐ Not Applicable ☐ Not Reported ☒ (Was an advisory group formed in the planning/execution of the study?) |
| Inclusion/ Exclusion Criteria | Included: parents with children aged 12-17 years, English speaking and reading |
| **Methods of Recruitment, Retention & Consent** | |
| Online methods/platforms used for recruitment | Qualtrics (“because previous studies reported that Qualtrics panels can achieve demographic attributes within a 10% range of their corresponding values in the US population.”) Pg. 352 |
| Offline methods used for recruitment | NA |
| Methods/tools used for online consent/assent acquisition | Qualtrics |
| Caregiver consent acquired/ waived (If waived, by what authority?) | Yes ☒ No ☐ Unclear ☐ Not Applicable ☐ Not Reported ☐  “Parents were then asked to allow their children to provide assent and complete the baseline survey independently and privately.” Pg. 352 |
| Children assent acquisition | Yes ☒ No ☐ Unclear ☐ Not Applicable ☐ Not Reported ☐ |
| What, if any, methods were used to validate that assent/consent was informed? | Not reported  (E.g. validation questions, CAPTCHA, email confirmation, video-conferencing verbal) |
| Pushes/reminders sent to participants | Enrolment in trial Yes ☐ No ☒ Unclear ☐ Not Applicable ☐ Not Reported ☐; Retention/completion in trial Yes ☐ No ☒ Unclear ☐ Not Applicable ☐ Not Reported ☐; Follow-up in trial Yes ☒ No ☐ Unclear ☐ Not Applicable ☐ Not Reported ☐ |
| **Interventions** | |
| Number of arms | 2 |
| Intervention | “The family media use plan contains a series of topic areas with potential rules for media use at home. Parent-adolescent dyads in the intervention group were asked to select the rules or guidelines that they endorsed or wanted to implement at home. Parent-adolescent dyads could also add their ideas through a write-in option. After completing the plan, participants were instructed to either print or take a screenshot of the plan and place it in a prominent place.” Pg. 353 |
| Comparison | “After completing the baseline survey, parent-adolescent dyads in the control group were not provided any information about the family media use plan. All control participants were contacted again 2 months later by an email from Qualtrics that invited them to complete the follow-up survey.” Pg. 353 |
| Operating Systems/Devices required for participants’ engagement with trial | Not reported  (E.g. computer, phone, iOS, Windows, Android, iPhone, Facebook or Instagram account etc.) |
| Tools/software used for data protection processes | Qualtrics and a study website  (E.g. dedicated website, REDCap, institutional server etc.) |
| Tools/methods used for data collection | Qualtrics and a study website.  (E.g. data submitted via website, video-conferencing etc.) |
| Did caregivers participate/ contribute data to the trial? If so, describe. | Yes ☒ No ☐ Unclear ☐ Not Applicable ☐ Not Reported ☐  The media-use plan was a family-wide intervention. |
| Duration of intervention from first to final engagement | 2-months |
| Compensation offered to participants | Not reported |
| **Outcomes** | |
| Results relevant to this scoping review | NA  (E.g. satisfaction with online methods of trial, demographics of participants’ in relation to their outcomes especially in regarding online components, i.e., recruitment, retention, completion etc) |
| Baseline differences between completers & non-completers | “No statistically significant difference between groups was found. In a sensitivity analysis that included all 1520 participants after multiple imputation of missing data, the corresponding within group changes were similar: –0.1 (95% CI, –0.8 to 0.6) for the intervention group and 0.2 (95% CI, –0.5 to 0.8) for the control group.” Pg. 354 |
| Attrition rates of randomised participants | \| Randomised \| FU completed \| % attrition; Intervention \| 760 \| 430 \| 43.4; Control \| 760 \| 360 \| 52.6 |
| Limitations of study as described by authors | Generalizability, loss to follow-up, |
| Funding source | Institutional, i.e. governmental, university \| Yes ☐ No ☒ Unclear ☐ Not Applicable ☐ Not Reported ☐; Private, i.e. pharmaceutical, software company \| Yes ☒ No ☐ Unclear ☐ Not Applicable ☐ Not Reported ☐ This study was supported by a research agreement with Facebook. |
| Other aspects of study pertinent to online trial conception and execution | NA |

| Risk of Bias assessment  ([**https://handbook-5-1.cochrane.org/chapter_8/8_assessing_risk_of_bias_in_included_studies.htm**](https://handbook-5-1.cochrane.org/chapter_8/8_assessing_risk_of_bias_in_included_studies.htm)**)** | | | | |
| --- | --- | --- | --- | --- |
| Bias domain | Source of bias | Risk of bias | Support for judgment (use direct quotes where possible with explanatory comments) | Location in text or source (pg. number, figure, table etc.) |
| Selection bias | Random sequence generation | Low risk ☒    High risk ☐  Unclear risk ☐ | “After individuals were deemed eligible, a computer algorithm within the Qualtrics platform randomized participants to the intervention group or control group using a 1:1 ratio.” | Pg. 352, under Study Procedures |
|  | Allocation concealment | Low risk ☒    High risk ☐  Unclear risk ☐ | “Investigators were blind to the randomization process.” | Pg. 352, under Study Procedures |
| Performance bias | Blinding of participants and personnel | Low risk ☐    High risk ☐  Unclear risk ☒ | Comment: Not reported, however blinding would have been difficult to achieve and may have influenced outcomes/self-reporting. | Not reported |
| Detection bias | Blinding of outcome assessment | Low risk ☐    High risk ☒  Unclear risk ☐ | Comment: All outcomes were self-reported, assessor blinding is not reported. | Not reported |
| Attrition bias | Incomplete outcome data | Low risk ☐    High risk ☒  Unclear risk ☐ | Comment:  Intervention: 760 randomised, 430 completed Follow-up: 43.34% attrition rate  Control: 760 randomised, 360 completed follow-up: 52.6% attrition rate | Pg. 353, Participant flow diagram |
| Reporting bias | Selective reporting | Low risk ☒    High risk ☐  Unclear risk ☐ | Comment: The study protocol is available and the outcomes assessed were those that were pre-specified.  [poi200090supp1_prod_1616606186.91397.pdf (jamanetwork.com)](https://cdn.jamanetwork.com/ama/content_public/journal/peds/938672/poi200090supp1_prod_1616606186.91397.pdf?Expires=1661064348&Signature=BOI6QaOlVnDJOUwlRaj6-Tk~RdqM8X8JQ2AiEg9~HLcQR1ycpBwVDjOvHajjEIZoq9k~tLuTCLv1JjktbOcjfIheGlk93M9PL5x7U6o4VBhN5LSnKzHOx1UVOfJZruhMIDzmqb0NEoJ2PS0Stk9aCy7CSFP2kZlm6p2Nybv5NUwz0Rswd5nJNfvB0CdYPVYTS5Py8RyR0S~tytI9q~rMCEyff5c3lgZI2O6MeJQ1MlX8Oa9qXarEfPUmH-cKzC0u7DVLSQnxOvisbV2PW00ARh1hfjhr3vJ1cgefjeRWQKbC8EpYTNlsFWcula4dwf1p0RqTT0IgagWXUGYYi3yK0Q__&Key-Pair-Id=APKAIE5G5CRDK6RD3PGA) | Protocol from supplementary data |
| Other bias | Anything else, ideally pre-specified | Low risk ☐    High risk ☒  Unclear risk ☐ | “Fourth, although we can assess completed family media use plan in the intervention group, we cannot determine whether a plan was created by a parent-adolescent dyad as intended or whether a parent or adolescent created it separately.” | Pg. 356, under Limitations |
