## Supplementary material for "Online randomised trials with children: A scoping review": S7 Appendix: Nelson.docx

| **Basic Study Characteristics** | |
| --- | --- |
| **Citation** | Nelson KM, Perry NS, Stout CD, Dunsiger SI, Carey MP. The Young Men and Media Study: A Pilot Randomized Controlled Trial of a Community-Informed, Online HIV Prevention Intervention for 14–17-Year-Old Sexual Minority Males. AIDS & Behavior. 2022;26(2):569–83. |
| **Characteristic reported (indicate with X for decision or enter free text, where applicable)** | |
| Type of study | Randomised trial ☒ \| Cluster randomised ☐; Quasi-randomised trial ☐ \| Individually randomised ☒ Pilot* |
| Type of publication | Peer-reviewed journal ☒; Pre-print ☐; Other, if so, describe ☐ |
| Aim of study as per author’s description | To increase sexual health knowledge, promote critical examination of pornography, and decrease sexual risk among adolescent sexual minority males (ASMM). |
| **Participant Characteristics** | |
| Participant age demographics | Age range \| 14-17; Mean age \| 16 |
| Country(ies) of Participants’ Residence | U.S.A. |
| Number of participants randomised | Children 154; Caregivers NA |
| Gender demographics of participants | 100% male |
| Socioeconomic status of participants | Not reported |
| Race/Ethnicity of participants | 52% White  26% Latino  11% Black/African American  11% Mixed race/other |
| Type of participant targeted | adolescent sexual minority males (ASMM), focused on racial and ethnic minorities as well to increase their recruitment in the study |
| Children’s advisory group formation | Ages of advisory group participants \| Cross-sectional survey (207 14- to 17- year olds) AND youth advisory board (4-5, 16- to 18- year olds); Means of recruiting advisory group participants \| Not reported; Phases of trial involvement \| informed design, content, and form of the intervention website. Yes ☒ No ☐ Unclear ☐ Not Applicable ☐ Not Reported ☐ (Was an advisory group formed in the planning/execution of the study?) |
| Inclusion/ Exclusion Criteria | Included: cisgender male, self-identify as gay/bisexual, report being sexually attracted to males, and/or report having voluntary sexual contact with a male partner (past year), have intentionally viewed pornography, reside in the U.S., have a personal email address, new to the study |
| **Methods of Recruitment, Retention & Consent** | |
| Online methods/platforms used for recruitment | Social media ads/posts (FB/IG) with GIFs featuring young men and emoji and written text describing how participating in the study could help researchers develop and test a more inclusive online sexual health program |
| Offline methods used for recruitment | NA |
| Methods/tools used for online consent/assent acquisition | Consent on dedicated website, hosted with REDCap |
| Caregiver consent acquired/ waived (If waived, by what authority?) | Yes ☐ No ☒ Unclear ☐ Not Applicable ☐ Not Reported ☐  Waiver of guardian consent was approved by The Boston Medical Center and Boston University Medical Campus Institutional Review Board |
| Children assent acquisition | Yes ☒ No ☐ Unclear ☐ Not Applicable ☐ Not Reported ☐ |
| What, if any, methods were used to validate that assent/consent was informed? | Participants were given 3 tries to verify/validate consent. Capacity to consent was confirmed via four questions that evaluated respondents’ ability to:  (E.g. validation questions, CAPTCHA, email confirmation, video-conferencing verbal)   1) name things they would be expected to do during the study, 2) understand randomization procedures, 3) to explain what they would do if they experienced distress during the study, 4) identify potential risks of participation: : (1) “If you agree to be in this study, what are we asking you to do?” (2) “How will it be decided which group of the study you are assigned to?” (3) “What can you do if you experience distress while taking part in this study?” and (4) “What are the potential risks of being in this study?” |
| Pushes/reminders sent to participants | Enrolment in trial Yes ☐ No ☒ Unclear ☐ Not Applicable ☐ Not Reported ☐; Retention/completion in trial Yes ☒ No ☐ Unclear ☐ Not Applicable ☐ Not Reported ☐; Follow-up in trial Yes ☐ No ☐ Unclear ☐ Not Applicable ☒ Not Reported ☐ “If a participant had not logged in, they were sent up to three reminder emails in a 10-day period.” Pg. 3 |
| **Interventions** | |
| Number of arms | 2 |
| Intervention | After randomisation, intervention participants were sent an email with a link, log-in, and temp password to the website and asked to complete all intervention modules (9) within 3 weeks of receiving the email. Modules were interactive (games, videos, animations) with an interface like Netflix and a responsive design. |
| Comparison | Automatically sent an email with links to the centers for disease control and prevention (CDC), HIV prevention, and the national HIV and STD testing resource websites. They were encouraged to visit at least one of those sites within 3 weeks of receiving the email. |
| Operating Systems/Devices required for participants’ engagement with trial | “Responsive design”, meaning it worked on a mobile device, tablet, or computer  (E.g. computer, phone, iOS, Windows, Android, iPhone, Facebook or Instagram account etc.) |
| Tools/software used for data protection processes | REDCap via dedicated website  (E.g. dedicated website, REDCap, institutional server etc.) |
| Tools/methods used for data collection | Dedicated Study Website  (E.g. data submitted via website, video-conferencing etc.) |
| Did caregivers participate/ contribute data to the trial? If so, describe. | Yes ☐ No ☒ Unclear ☐ Not Applicable ☐ Not Reported ☐ |
| Duration of intervention from first to final engagement | 3 weeks + 3-month follow-up |
| Compensation offered to participants | “Participants were compensated via electronic gift cards at the completion of the baseline assessment ($15) and after each of the follow-up assessments ($25 for post-intervention, $35 for 3-month follow-up). Participants were given a $20 bonus if they completed all three assessments.” Pg.4 |
| **Outcomes** | |
| Results relevant to this scoping review | “Across all modules, on average, intervention participants rated the intervention content 4.3 stars out of 5 (SD =0.8).” Pg.9  (E.g. satisfaction with online methods of trial, demographics of participants’ in relation to their outcomes especially in regarding online components, i.e., recruitment, retention, completion etc) |
| Baseline differences between completers & non-completers | Not reported |
| Attrition rates of randomised participants | \| Randomised \| Post-intervention completed \| 3-month FU completed \| % attrition at 3-month FU; Intervention \| 77 \| 71 \| 67 \| 13%; Control \| 77 \| 66 \| 65 \| 15.6% |
| Limitations of study as described by authors | Possibly not generalizable to all ASMM. Covid-19 prevented following of sexual behaviours as many people were socially isolated. |
| Funding source | Institutional, i.e. governmental, university \| Yes ☒ No ☐ Unclear ☐ Not Applicable ☐ Not Reported ☐; Private, i.e. pharmaceutical, software company \| Yes ☐ No ☒ Unclear ☐ Not Applicable ☐ Not Reported ☐ |
| Other aspects of study pertinent to online trial conception and execution | “To protect against fraudulent or duplicative enrolments, screening and baseline survey responses were cross-referenced using age (age vs. date of birth), location (zip code vs. state of residence), sexual activity (multiple questions across the screener and survey assessing sexual behavior), and email address.” Pg. 3 |

| Risk of Bias assessment  ([**https://handbook-5-1.cochrane.org/chapter_8/8_assessing_risk_of_bias_in_included_studies.htm**](https://handbook-5-1.cochrane.org/chapter_8/8_assessing_risk_of_bias_in_included_studies.htm)**)** | | | | |
| --- | --- | --- | --- | --- |
| Bias domain | Source of bias | Risk of bias | Support for judgment (use direct quotes where possible with explanatory comments) | Location in text or source (pg. number, figure, table etc.) |
| Selection bias | Random sequence generation | Low risk ☒    High risk ☐  Unclear risk ☐ | “After confirming eligibility, participants were randomized to intervention or control based on a permuted block randomization procedure, with small, random-sized blocks. Randomization assignment was given out via REDCap.” | Pg. 3, under Procedures |
|  | Allocation concealment | Low risk ☐    High risk ☐  Unclear risk ☒ | Comment: Insufficient information to assess risk. | Not reported |
| Performance bias | Blinding of participants and personnel | Low risk ☒    High risk ☐  Unclear risk ☐ | Comment: No mention of blinding, but the review authors judge that the outcome is not likely influenced by lack of blinding. | Not reported |
| Detection bias | Blinding of outcome assessment | Low risk ☐    High risk ☐  Unclear risk ☒ | Comment: Insufficient information to assess risk. | Not reported |
| Attrition bias | Incomplete outcome data | Low risk ☒    High risk ☐  Unclear risk ☐ | Comment:  Intervention: post-test completed: 71/77: 8% attrition  3-mo follow-up: 67/77: 13% attrition  Control: post-test completed: 66/77: 14% attrition  3-mo follow-up: 65/77: 15% attrition | Pg. 8, Figure 1 |
| Reporting bias | Selective reporting | Low risk ☒    High risk ☐  Unclear risk ☐ | Comment: Study protocol is not available but the trial is registered at ClinicalTrials.gov and all pre-specified outcomes are not reported. The authors address this in the paper, citing the COVID-19 pandemic as severely limiting social contact and therefore limiting any behavioural data analysis.  “Although sexual behavior data were collected and planned to be analyzed with the other efficacy outcomes, study concurrence with the initial months of the COVID-19 pandemic in the United States reduced participants sexual contacts because most participants were no longer seeking out or interacting with sexual partners in-person [39]. Given this limitation we have chosen not to include sexual behavior data in our analyses.” | ClinicalTrials.gov: NCT04109443  Pg. 2, Footnote 1, under Methods |
| Other bias | Anything else, ideally pre-specified | Low risk ☒    High risk ☐  Unclear risk ☐ | Comment: No other bias detected. | Not applicable |
