## Supplementary material for "Online randomised trials with children: A scoping review": S7 Appendix: O'Connor.docx

| **Basic Study Characteristics** | |
| --- | --- |
| **Citation** | O’Connor K, Bagnell A, McGrath P, Wozney L, Radomski A, Rosychuk RJ, Curtis S, Jabbour M, Fitapatrick E, Johnson DW, Ohinmaa A, Joyce A, Newton A. An Internet-Based Cognitive Behavioral Program for Adolescents with Anxiety: Pilot Randomized Controlled Trial. JMIR Ment Health. 2020: 7(7) |
| **Characteristic reported (indicate with X for decision or enter free text, where applicable)** | |
| Type of study | Randomised trial ☒ \| Cluster randomised ☐; Quasi-randomised trial ☐ \| Individually randomised ☒ Pilot* |
| Type of publication | Peer-reviewed journal ☒; Pre-print ☐; Other, if so, describe ☐ |
| Aim of study as per author’s description | “to pilot procedures and obtain data on methodological processes and intervention satisfaction to determine the feasibility of a definitive randomized controlled trial (RCT) to test the effectiveness of a self-managed ICBT (internet-based cognitive behavioural therapy) program, Breathe (Being Real, Easing Anxiety: Tools Helping Electronically), for adolescents with anxiety concerns.” Pg. 1 |
| **Participant Characteristics** | |
| Participant age demographics | Age range \| 13-17; Mean age \| 15.3 |
| Country(ies) of Participants’ Residence | Canada |
| Number of participants randomised | Children 94; Caregivers N |
| Gender demographics of participants | 90% female |
| Socioeconomic status of participants | Not reported |
| Race/Ethnicity of participants | Not reported |
| Type of participant targeted | Children at risk for anxiety |
| Children’s advisory group formation | Yes ☒ No ☐ Unclear ☐ Not Applicable ☐ Not Reported ☐ (Was an advisory group formed in the planning/execution of the study?) Ages of advisory group participants \| Not reported; Means of recruiting advisory group participants \| Not reported; Phases of trial involvement \| “The program underwent an evaluation for usability with adolescents and clinicians before the start of the trial to improve the intervention’s technical interface, therapeutic messaging, and user experience (eg, esthetics, presentation of rating scales) [28].” |
| Inclusion/ Exclusion Criteria | Included: read/write English, regular access to a phone and a computer system with a high-speed internet service, able to use a computer to interact with web-based material, and reported the presence of anxiety symptoms  Excluded: self-report of suicidal thoughts in the past week; initially there was a second exclusion criterion, receipt of face-to-face CBT; however, this was removed halfway through cycle 1 due to emails from prospective participants who found it confusing |
| **Methods of Recruitment, Retention & Consent** | |
| Online methods/platforms used for recruitment | Three recruitment cycles:   1) Offline 2) Mixture: Distribution of study updates through email (MailChimp) on a monthly basis and fostering relationships between Breathe research staff and recruitment partners through teleconferences and site visits, as requested 3) Social media recruitment via Facebook, Twitter, and Instagram with posts that appeared when adolescents searched or posted about anxiety or stress. These adolescents were directed to the study website |
| Offline methods used for recruitment | 1) health care professionals provided study pamphlets to prospective participants seeking mental health care from EDs, mobile or school-based crisis teams, and primary care clinics 2) Distribution of study updates through email (MailChimp) on a monthly basis and fostering relationships between Breathe research staff and recruitment partners through teleconferences and site visits, as requested |
| Methods/tools used for online consent/assent acquisition | Consent/assent was indicated electronically via the secure myStudies website.  2 stages:   1) Adolescents screened via a secure web-based process, myStudies, A Connec Service URL. Telephone-based and email support during this stage were available from a research team member 2) Those that met the first set of criteria were screened via the secure, internet-based platform, Intelligent Research Intervention Software (IRIS) |
| Caregiver consent acquired/ waived (If waived, by what authority?) | Yes ☒ No ☐ Unclear ☐ Not Applicable ☐ Not Reported ☐  “Adolescents aged 15 to 17 years were asked to consent to the study on their own behalf; adolescents aged 13 and 14 years were asked to assent to study participation. We also required parental consent for all adolescents aged 13 and 14 years, even if they were assessed as being able to consent. The intent was to have parents involved so that they could support their child with the enrolment process.” Pg. 4 |
| Children assent acquisition | Yes ☒ No ☐ Unclear ☐ Not Applicable ☐ Not Reported ☐ |
| What, if any, methods were used to validate that assent/consent was informed? | “The first webpage confirmed that the individual understood that he/she could ask questions about the study at any time during the study or in the future. Each webpage included a ‘Contact Us’ button that provided a pop-up email box with a toll-free phone and email contact info for a member of the research team. The Contact Us button triggered a message to the participant that a research team member would contact them to answer any questions they may have before proceeding with consent/assent.” Pg. 4  (E.g. validation questions, CAPTCHA, email confirmation, video-conferencing verbal)  They could also save or print the consent/assent form. They were also guided through sections describing participation, right to withdraw, confirmation questions, risks, benefits and True/False questions were added before the final consent/assent.  Incorrect answers triggered a pop-up box with the correct answer and explanation |
| Pushes/reminders sent to participants | Enrolment in trial Yes ☐ No ☒ Unclear ☐ Not Applicable ☐ Not Reported ☐; Retention/completion in trial Yes ☒ No ☐ Unclear ☐ Not Applicable ☐ Not Reported ☐; Follow-up in trial Yes ☐ No ☒ Unclear ☐ Not Applicable ☐ Not Reported ☐ IRIS platform sent emails to participants who did not log in for 1 week encouraging them to complete their weekly module. |
| **Interventions** | |
| Number of arms | 2 |
| Intervention | Breathe is an 8-module CBT program where participants were encouraged to complete 1 module per week. Adolescents were given a choice if they wanted parents to receive an email that included educational materials about the nature of adolescent anxiety and highlights of key topics that they worked on for that module. |
| Comparison | Control group received minimal intervention “—access to a secure, password-protected static study webpage housed in IRIS. The website offered suggested anxiety-related trade publications, print-based workbooks for adolescents, and the names of national and local organizations and websites where the adolescent might find support. There was no interactivity or personalization included in the webpage. Adolescents assigned to the control group were provided with the option to access the Breathe program for clinical use at the end of their 8-week control group participation.” Pg. 7 |
| Operating Systems/Devices required for participants’ engagement with trial | Not reported  (E.g. computer, phone, iOS, Windows, Android, iPhone, Facebook or Instagram account etc.) |
| Tools/software used for data protection processes | Breathe was delivered via IRIS, the same platform used for eligibility screening.  (E.g. dedicated website, REDCap, institutional server etc.) |
| Tools/methods used for data collection | IRIS hosted both the Breathe website and the static webpage  (E.g. data submitted via website, video-conferencing etc.) |
| Did caregivers participate/ contribute data to the trial? If so, describe. | Yes ☐ No ☒ Unclear ☐ Not Applicable ☐ Not Reported ☐ |
| Duration of intervention from first to final engagement | 8 weeks + 3-month follow-up |
| Compensation offered to participants | No |
| **Outcomes** | |
| Results relevant to this scoping review | “All participants liked that the program was completed on the web, with 79% (11/14) indicating no concerns with privacy. Responses were divided as to whether the program should include a social media component (5/14 in agreement), be more personalized to the participant (7/14 in agreement), and include a module for parents (8/14 in agreement). The most common barriers to program completion were difficulty completing exposure activities and remembering/finding time to complete modules, among other life commitments.” Pg. 12  (E.g. satisfaction with online methods of trial, demographics of participants’ in relation to their outcomes especially in regarding online components, i.e., recruitment, retention, completion etc) |
| Baseline differences between completers & non-completers | “Program completers and noncompleters did not differ significantly in their responses to any of the 4 ASQ screening questions (P=.32, .93, .49, and .49), the manner in which they learned about the study (social media/on the web, health care provider/guidance counselor, friend, or not specified; P=.17), age (P=.85), or baseline MASC2 T scores (P=.44). Completers and noncompleters could not be compared on self-identified gender due to the limited number of males enrolled in the study.” Pg. 11 |
| Attrition rates of randomised participants | \| Randomised \| 8-week FU completed \| % attrition at 8-weeks \| 3-month FU completed \| % attrition; Intervention \| 49 \| 13 \| 73.5 \| 11 \| 77.6; Control \| 45 \| 11 \| 75.6 \| NA \| NA |
| Limitations of study as described by authors | “The most significant limitation of this pilot RCT was the lack of data at post-treatment and 3-month follow-up. Another important limitation was our reliance on adolescents’ own recall when providing information about their utilization of other health care services. A final limitation was the exclusion of 150 adolescents early on in the trial due to their report of CBT participation. Although we do not know how many of these adolescents would have consented/assented to participate in Breathe, their exclusion introduces the potential for further selection bias in the study.” Pg. 14 |
| Funding source | Institutional, i.e. governmental, university \| Yes ☒ No ☐ Unclear ☐ Not Applicable ☐ Not Reported ☐; Private, i.e. pharmaceutical, software company \| Yes ☐ No ☒ Unclear ☐ Not Applicable ☐ Not Reported ☐ |
| Other aspects of study pertinent to online trial conception and execution | “In testing different recruitment strategies, however, we learned that recruitment was most successful via social media (39/94, 42% enrolled). We were able to recruit approximately 4 times the number of adolescents in cycle 3 (n=75) once we launched our social media strategy, compared with cycles 1 and 2 during which we relied on health care providers.” Pg. 14  Screening and eligibility will be stream-lined in full RCT as this was burdensome for participants. |

| Risk of Bias assessment  ([**https://handbook-5-1.cochrane.org/chapter_8/8_assessing_risk_of_bias_in_included_studies.htm**](https://handbook-5-1.cochrane.org/chapter_8/8_assessing_risk_of_bias_in_included_studies.htm)**)** | | | | |
| --- | --- | --- | --- | --- |
| Bias domain | Source of bias | Risk of bias | Support for judgment (use direct quotes where possible with explanatory comments) | Location in text or source (pg. number, figure, table etc.) |
| Selection bias | Random sequence generation | Low risk ☐    High risk ☒  Unclear risk ☐ | “Adolescents were randomly assigned using a computer-generated allocation sequence with a 1:1 ratio to 1 of 2 groups.”  “A final limitation was the exclusion of 150 adolescents early on in the trial due to their report of CBT participation. Although we do not know how many of these adolescents would have consented/assented to participate in Breathe, their exclusion introduces the potential for further selection bias in the study.”  Comment: The study design change after the exclusion of 150 possible participants could have introduced additional bias. | Pg. 4, under Randomization and Blinding  Pg. 14, under Limitations |
|  | Allocation concealment | Low risk ☐    High risk ☒  Unclear risk ☐ | “A graduate student trainee affiliated with the project generated this sequence and an email was sent to each participant with information on their assigned intervention and log-in/website information to begin participation. A permuted block randomization procedure [27] with random block sizes of 4 to 6 was used. Given the methodological objectives of the pilot study, no blinding took place.”  Comment: Allocation was not concealed. | Pg. 4, under Randomization and Blinding |
| Performance bias | Blinding of participants and personnel | Low risk ☐    High risk ☒  Unclear risk ☐ | “Given the methodological objectives of the pilot study, no blinding took place.”  Comment: The lack of blinding likely influenced the outcome. | Pg. 4, under Randomization and Blinding |
| Detection bias | Blinding of outcome assessment | Low risk ☐    High risk ☒  Unclear risk ☐ | Comment: No blinding, and the review authors judge that the outcome measurement is likely to be influenced by lack of blinding as all data were-self-reported. | Pg. 4, under Randomization and Blinding |
| Attrition bias | Incomplete outcome data | Low risk ☐    High risk ☒  Unclear risk ☐ | Comment:  Intervention 13/49 completed 8-week follow-up: 73.5% attrition rate  11/49 completed 3-mo follow-up: 77.6% attrition rate  Control: 11/45 completed 8-week follow-up: 75.6% attrition rate  NA 3-mo follow-up | Pg. 10, Fig. 3 |
| Reporting bias | Selective reporting | Low risk ☒    High risk ☐  Unclear risk ☐ | Comment: No study protocol available but the study is registered at ClinicalTrials.gov and all outcomes were assessed with the exception of MCID, which the authors address below.  “However, challenges with retention did not permit us to calculate an MCID as planned.” | ClinicalTrials.gov: NCT02059226  Pg. 14, under Principal Findings |
| Other bias | Anything else, ideally pre-specified | Low risk ☒    High risk ☐  Unclear risk ☐ | Comment: No other bias detected | Not applicable |
