## Supplementary material for "Online randomised trials with children: A scoping review": S7 Appendix: O'Dea.docx

| **Basic Study Characteristics** | |
| --- | --- |
| **Citation** | O’Dea B, Han J, Batterham PJ, Achilles MR, Calear AL, Werner-Seidler A, Parker B, Shand F, Christensen H. A randomised controlled trial of a relationship-focussed mobile phone application for improving adolescents’ mental health. Journal of Child Psychology and Psychiatry 61:8 (2020), pp 899-913 |
| **Characteristic reported (indicate with X for decision or enter free text, where applicable)** | |
| Type of study | Randomised trial ☒ \| Cluster randomised ☐; Quasi-randomised trial ☐ \| Individually randomised ☒ |
| Type of publication | Peer-reviewed journal ☒; Pre-print ☐; Other, if so, describe ☐ |
| Aim of study as per author’s description | To evaluate the acceptability and effectiveness of a relationship-focussed mobile phone application (WeClick) on improving wellbeing, help-seeking for mental health in adolescents. |
| **Participant Characteristics** | |
| Participant age demographics | Age range \| 12-16; Mean age \| 14.82 |
| Country(ies) of Participants’ Residence | Australia |
| Number of participants randomised | Children/ participants: 193; Caregivers NA |
| Gender demographics of participants | 86.5% female |
| Socioeconomic status of participants | Not reported |
| Race/Ethnicity of participants | 3.62% Aboriginal and/or Torres Strait Islander |
| Type of participant targeted | No specific group targeted. |
| Children’s advisory group formation | (Was an advisory group formed in the planning/execution of the study?) Yes ☒ No ☐ Unclear ☒ Not Applicable ☐ Not Reported ☐ Ages of advisory group participants \| Not reported; Means of recruiting advisory group participants \| Not reported; Phases of trial involvement \| “The app content was reviewed by young people who deemed it to be helpful and relatable.” |
| Inclusion/ Exclusion Criteria | Included: Australia residents, fluent in English, could provide parental consent, internet access, active email address, access to a mobile phone |
| **Methods of Recruitment, Retention & Consent** | |
| Online methods/platforms used for recruitment | Recruitment was through a Facebook ad campaign. After viewing the ad, prospective participants were directed to the study website, which provided information and a brief online screener to assess the inclusion criteria. |
| Offline methods used for recruitment | NA |
| Methods/tools used for online consent/assent acquisition | . If eligible, prospective participants were invited to create a study account, download and complete the consent (including parental signature), and return the forms via email. |
| Caregiver consent acquired/ waived (If waived, by what authority?) | Yes ☒ No ☐ Unclear ☐ Not Applicable ☐ Not Reported ☐  Parental signature required |
| Children assent acquisition | Yes ☒ No ☐ Unclear ☐ Not Applicable ☐ Not Reported ☐ |
| What, if any, methods were used to validate that assent/consent was informed? | No  (E.g. validation questions, CAPTCHA, email confirmation, video-conferencing verbal) |
| Pushes/reminders sent to participants | Enrolment in trial Yes ☒ No ☐ Unclear ☐ Not Applicable ☐ Not Reported ☐; Retention/completion in trial Yes ☒ No ☐ Unclear ☐ Not Applicable ☐ Not Reported ☐; Follow-up in trial Yes ☒ No ☐ Unclear ☐ Not Applicable ☐ Not Reported ☐ “The research team reviewed the consent forms upon receipt. Using the online research platform, the research team approved participants, which triggered an automatic email/ SMS invitation to complete the baseline survey. Participants also received an SMS and email invitation for the post-test and follow-up surveys. All surveys remained active for 5 days, with two reminders.” Pg. 902 |
| **Interventions** | |
| Number of arms | 2 |
| Intervention | “Taking approximately one hour to complete, the WeClick app was designed as a brief ‘single session’ intervention.” Pg. 900  It is an interactive story-telling app with four characters, each facing different relationship difficulties (e.g., family, peer, intimate, other adolescent issues). The users selects a character and work through activities that aim to develop the skills to overcome negative thinking and problem solving. |
| Comparison | The control group was a wait list and were provided access to the app after completing the 4-week post-test survey |
| Operating Systems/Devices required for participants’ engagement with trial | iOS or Android phone, email account, App was downloaded through Apple and Google Play stores and participants had four weeks of access using their study code  (E.g. computer, phone, iOS, Windows, Android, iPhone, Facebook or Instagram account etc.) |
| Tools/software used for data protection processes | Data were collected and stored securely via the Black Dog Institute online research platform. Users created a password-protected account and were allocated a unique id number. All other identifiers were removed before data were downloaded into Excel and exported to SPSS Version 22.0 for analysis  (E.g. dedicated website, REDCap, institutional server etc.) |
| Tools/methods used for data collection | Dedicated website via the Black Dog Institute online research platform  (E.g. data submitted via website, video-conferencing etc.) |
| Did caregivers participate/ contribute data to the trial? If so, describe. | Yes ☐ No ☒ Unclear ☐ Not Applicable ☐ Not Reported ☐ |
| Duration of intervention from first to final engagement | SSI +4 week post-test + 12-week follow-up |
| Compensation offered to participants | “15AUD voucher for each survey completed, with a maximum study reimbursement of 45AUD.” Pg. 902 |
| **Outcomes** | |
| Results relevant to this scoping review | “More than 90% reported that the app was enjoyable, easy to understand, and that they would recommend it to a friend.” Pg. 909  (E.g. satisfaction with online methods of trial, demographics of participants’ in relation to their outcomes especially in regarding online components, i.e., recruitment, retention, completion etc) |
| Baseline differences between completers & non-completers | “There were no differences in the rates of missingness in the data between conditions at post-test (p = .44) or follow-up (p = .38). There were also no associations between age, gender or experience of previous mental illness and completion of the post-test or follow-up assessments (all p > .05).” Pg. 905 |
| Attrition rates of randomised participants | \| Randomised \| 4-week FU completed \| % attrition \| 12-week FU completed \| % attrition (from randomisation); Intervention \| 98 \| 80 \| 18.4 \| 55 \| 43.9; Control \| 95 \| 82 \| 13.7 \| 60 \| 36.8 |
| Limitations of study as described by authors | Small sample size, over-representation of females, short follow-up time |
| Funding source | Institutional, i.e. governmental, university \| Yes ☒ No ☐ Unclear ☐ Not Applicable ☐ Not Reported ☐; Private, i.e. pharmaceutical, software company \| Yes ☐ No ☒ Unclear ☐ Not Applicable ☐ Not Reported ☐ |
| Other aspects of study pertinent to online trial conception and execution | “The use of Facebook recruitment enabled a sample of Australian youth to be recruited in less than 10 weeks.” Pg. 911 |

| Risk of Bias assessment  ([**https://handbook-5-1.cochrane.org/chapter_8/8_assessing_risk_of_bias_in_included_studies.htm**](https://handbook-5-1.cochrane.org/chapter_8/8_assessing_risk_of_bias_in_included_studies.htm)**)** | | | | |
| --- | --- | --- | --- | --- |
| Bias domain | Source of bias | Risk of bias | Support for judgment (use direct quotes where possible with explanatory comments) | Location in text or source (pg. number, figure, table etc.) |
| Selection bias | Random sequence generation | Low risk ☒    High risk ☐  Unclear risk ☐ | “Randomisation was carried out according to the International Council for Harmonisation (ICH) guidelines (Lewis, 1999) and performed immediately after participants completed baseline using a computerised adaptive randomisation procedure hosted by the Black Dog Institute’s online research platform. A stratification approach with a block size of 4 (1: 1 ratio) was used to ensure balance across the two conditions for age (12– 14 years vs. 15–16 years) and gender (male vs. female).” | Pg. 901, under Randomisation and masking |
|  | Allocation concealment | Low risk ☒    High risk ☐  Unclear risk ☐ | “Although participants and researchers were not blinded to the allocation assignment, the allocation was fully automatic with no interference from researchers.” | Pg. 901, under Randomisation and masking |
| Performance bias | Blinding of participants and personnel | Low risk ☐    High risk ☒  Unclear risk ☐ | Comment: As described above. The lack of blinding likely influenced the outcome. | Pg. 901, under Randomisation and masking |
| Detection bias | Blinding of outcome assessment | Low risk ☐    High risk ☒  Unclear risk ☐ | “To support the validity of the analysis, analyst triangulation was used whereby higher-order codes and final themes were determined by consensus among the researchers (Patton, 1999; Tracy, 2010). Furthermore, the researchers coding the data regularly reflected on their personal reactions and were considerate not to contaminate the data.”  Comment: All data were self-reported and allocation was not blinded and therefore outcomes were likely influenced by these factors. | Pg. 905, under Data Storage and Analysis |
| Attrition bias | Incomplete outcome data | Low risk ☒    High risk ☐  Unclear risk ☐ | Comment:  Intervention: 80/98 completed at 4-wk follow-up: 18.4% attrition  55/98 completed at 12-wk follow-up: 43.9% attrition  Control: 82/95 completed 4-wk follow-up: 13.7% attrition  60/95 completed 3-mo follow-up: 36.8% attrition | Pg. 904, Fig. 2 |
| Reporting bias | Selective reporting | Low risk ☒    High risk ☐  Unclear risk ☐ | Comment: No study protocol available but the study is registered at the Australian and New Zealand Clinical Trials Register and all outcomes were assessed as pre-specified. | Anzctr.org.au: ACTRN12618001982202 |
| Other bias | Anything else, ideally pre-specified | Low risk ☒    High risk ☐  Unclear risk ☐ | Comment: No other bias detected. | Not applicable |
