## Supplementary material for "Online randomised trials with children: A scoping review": S7 Appendix: Parker.docx

| **Basic Study Characteristics** | |
| --- | --- |
| **Citation** | Parker AE, Scull TM, Morrison AM. DigiKnowIt News: Educating youth about pediatric clinical trials using an interactive, multimedia educational website. Journal of Child Health Care. Journal of Child Health Care: 2022; 26(1): 139-53 |
| **Characteristic reported (indicate with X for decision or enter free text, where applicable)** | |
| Type of study | Randomised trial ☒ \| Cluster randomised ☐; Quasi-randomised trial ☐ \| Individually randomised ☒ |
| Type of publication | Peer-reviewed journal ☒; Pre-print ☐; Other, if so, describe ☐ |
| Aim of study as per author’s description | “The aim of the current study was to evaluate the effectiveness of DigiKnowIt News for improving factors related to pediatric clinical trial participation.” Pg. 4 |
| **Participant Characteristics** | |
| Participant age demographics | Age range \| 8-14; Mean age \| 10.92 |
| Country(ies) of Participants’ Residence | U.S.A. |
| Number of participants randomised | Children/ participants: 132; Caregivers NA |
| Gender demographics of participants | 91 completed the post test questionnaire, of those: 49.45% female |
| Socioeconomic status of participants | Not reported |
| Race/Ethnicity of participants | 7.69% Black  15.38% Multiracial  2.2% Native American or Alaska Native  74.73% White  9% Hispanic, Latino, or Spanish descent  90.11% Not of Hispanic, Latino, or Spanish descent |
| Type of participant targeted | Attempted to recruit half healthy, and half chronically-ill children |
| Children’s advisory group formation | Ages of advisory group participants \| No; Means of recruiting advisory group participants \| No; Phases of trial involvement \| “Children were vital to include in the design to ensure that the visuals, content, and activities were appropriate and relevant.” Pg. 3 Yes ☒ No ☐ Unclear ☐ Not Applicable ☐ Not Reported ☐ (Was an advisory group formed in the planning/execution of the study?) |
| Inclusion/ Exclusion Criteria | Included: access to a computer or tablet with internet connection; ability to speak and read English; and no current or previous participation in a clinical trial after kindergarten. |
| **Methods of Recruitment, Retention & Consent** | |
| Online methods/platforms used for recruitment | Recruitment flyers and a recruitment website were advertised on social media and e-flyers shared with parents in schools |
| Offline methods used for recruitment | NA |
| Methods/tools used for online consent/assent acquisition | ” Eligible families reviewed and completed an online parent permission form and a child assent form located on the study recruitment website” |
| Caregiver consent acquired/ waived (If waived, by what authority?) | Yes ☒ No ☐ Unclear ☐ Not Applicable ☐ Not Reported ☐  “Parents received an email informing them that their child had been enrolled in the study and providing a link to access the child’s pretest questionnaire.” Pg. 5 |
| Children assent acquisition | Yes ☒ No ☐ Unclear ☐ Not Applicable ☐ Not Reported ☐ |
| What, if any, methods were used to validate that assent/consent was informed? | Not reported  (E.g. validation questions, CAPTCHA, email confirmation, video-conferencing verbal) |
| Pushes/reminders sent to participants | Enrolment in trial Yes ☐ No ☐ Unclear ☐ Not Applicable ☐ Not Reported ☒; Retention/completion in trial Yes ☐ No ☐ Unclear ☐ Not Applicable ☐ Not Reported ☒; Follow-up in trial Yes ☐ No ☐ Unclear ☐ Not Applicable ☐ Not Reported ☒ |
| **Interventions** | |
| Number of arms | 2 |
| Intervention | “The intervention included four core topic areas related to participating in clinical trials (i.e., importance of clinical trials, participant rights and safety, costs and benefits, and communication with researchers and medical professionals) in addition to one of two randomly selected procedure-specific areas (A: needles; B: scans).” Pg. 5 |
| Comparison | “Wait-list control participants did not receive access to the intervention after pretest. However, after completing the posttest, they were given the option to review DigiKnowIt News.” Pg. 5 |
| Operating Systems/Devices required for participants’ engagement with trial | Access to a computer or tablet with internet connection  (E.g. computer, phone, iOS, Windows, Android, iPhone, Facebook or Instagram account etc.) |
| Tools/software used for data protection processes | Not reported  (E.g. dedicated website, REDCap, institutional server etc.) |
| Tools/methods used for data collection | Links were sent to parents of participants via email to access the online questionnaires.  (E.g. data submitted via website, video-conferencing etc.) |
| Did caregivers participate/ contribute data to the trial? If so, describe. | Yes ☒ No ☐ Unclear ☐ Not Applicable ☐ Not Reported ☐  “Parents received an email informing them that their child had been enrolled in the study and providing a link to access the child’s pretest questionnaire. Parents of participants in the intervention groups were then provided via email with a link to access one of two customized versions (A/B) of DigiKnowIt News for their child to review.” “Parents of participating children were asked to respond to demographic questions on the eligibility screening questionnaire about their child including age, gender, grade, race, and ethnicity.” Pg. 5 |
| Duration of intervention from first to final engagement | 3 weeks |
| Compensation offered to participants | “Participants received US$20 for completing the pretest and US$30 for completing the posttest.” Pg. 5 |
| **Outcomes** | |
| Results relevant to this scoping review | “The vast majority of intervention participants responded affirmatively (agree/strongly agree) that they learned new information from using the website (88%); they could use the information they learned from the website to help them make decisions in the future (88%); they enjoyed using the website (78%); and they would tell a friend who wanted to know about clinical trials about the website (69%).” Pg. 10  (E.g. satisfaction with online methods of trial, demographics of participants’ in relation to their outcomes especially in regarding online components, i.e., recruitment, retention, completion etc) |
| Baseline differences between completers & non-completers | Not reported |
| Attrition rates of randomised participants | \| Randomised \| Pre-test completed \| % attrition \| Post-test completed \| % attrition; Intervention \| 64 \| 54 \| 15.6 \| 44 \| 31.3; Control \| 68 \| 51 \| 25 \| 47 \| 30.9 |
| Limitations of study as described by authors | Small sample size, only a quarter of the children in the sample had a chronic illness, limiting the geralizability of the findings to children with chronic illnesses. “The study could not determine how changes in knowledge, attitudes, beliefs, and self-efficacy might influence children’s decision-making process.” Pg. 11 |
| Funding source | Institutional, i.e. governmental, university \| Yes ☒ No ☐ Unclear ☐ Not Applicable ☐ Not Reported ☐; Private, i.e. pharmaceutical, software company \| Yes ☐ No ☒ Unclear ☐ Not Applicable ☐ Not Reported ☐ The authors are employees of innovation Research & Training (iRT). iRT has a financial interest in the sale of a commercially-available customizable version of the website. |
| Other aspects of study pertinent to online trial conception and execution | NA |

| Risk of Bias assessment  ([**https://handbook-5-1.cochrane.org/chapter_8/8_assessing_risk_of_bias_in_included_studies.htm**](https://handbook-5-1.cochrane.org/chapter_8/8_assessing_risk_of_bias_in_included_studies.htm)**)** | | | | |
| --- | --- | --- | --- | --- |
| Bias domain | Source of bias | Risk of bias | Support for judgment (use direct quotes where possible with explanatory comments) | Location in text or source (pg. number, figure, table etc.) |
| Selection bias | Random sequence generation | Low risk ☒    High risk ☐  Unclear risk ☐ | “After informed consent, participants were randomized, stratifying based upon gender (boy; girl), race/ethnicity (white and not Hispanic/Latino; nonwhite and/or Hispanic/Latino), and health status (healthy; chronic illness), to either the intervention group or wait-list control group.”  “An investigator generated the random allocation sequences using Excel, and the project coordinator assigned participants to condition according to those sequences.” | Pg. 5, under Procedures |
|  | Allocation concealment | Low risk ☐    High risk ☐  Unclear risk ☒ | ”…and the project coordinator assigned participants to condition according to those sequences.” | Pg. 5, under Procedures |
| Performance bias | Blinding of participants and personnel | Low risk ☒    High risk ☐  Unclear risk ☐ | “Participants were blind to assignment, but the research team was not. However, the questionnaires were web-based and not administered in person by the research team.” | Pg. 5, under Procedures |
| Detection bias | Blinding of outcome assessment | Low risk ☐    High risk ☐  Unclear risk ☒ | Comment: Insufficient evidence to make an assessment. | Not reported |
| Attrition bias | Incomplete outcome data | Low risk ☐    High risk ☒  Unclear risk ☐ | Comment:  Intervention: 44/64 completed post-test: 31.3% attrition  Control: 47/68 completed post-test: 30.9% attrition | Pg. 7, Fig. 1 |
| Reporting bias | Selective reporting | Low risk ☐    High risk ☒  Unclear risk ☐ | Comment: There is no study protocol, but it is registered at ClinicalTrials.gov and three of the pre-specified secondary outcomes i.e., Changes in Hope and Changes in Parent-Child Communication are not reported. | ClinicalTrials.gov: NCT03531866 |
| Other bias | Anything else, ideally pre-specified | Low risk ☒    High risk ☐  Unclear risk ☐ | Comment:  No other bias detected | Not applicable |
