## Supplementary material for "Online randomised trials with children: A scoping review": S7 Appendix: Radomski.docx

| **Basic Study Characteristics** | |
| --- | --- |
| **Citation** | Radomski AD, Bagnell A, Curtis S, Hartling L, Newton AS. Examining the Usage, User Experience, and Perceived Impact of an Internet-Based Cognitive Behavioral Therapy Program for Adolescents with Anxiety: Randomized Controlled Trial. JMIR Ment Health 2020; 7(2) |
| **Characteristic reported (indicate with X for decision or enter free text, where applicable)** | |
| Type of study | Randomised trial ☒ \| Cluster randomised ☐; Quasi-randomised trial ☐ \| Individually randomised ☒ |
| Type of publication | Peer-reviewed journal ☒; Pre-print ☐; Other, if so, describe ☐ |
| Aim of study as per author’s description | Using the iCBT program, Being Real, Easing Anxiety: Tools Helping Electronically (Breathe), the authors aimed to describe intervention use, describe and compare user experiences between groups and calculate an MCID for anxiety and explore relationships between iCBT use, experiences, and treatment response among Breathe respondents. Pg.1 |
| **Participant Characteristics** | |
| Participant age demographics | Age range \| 13-19; Mean age \| 16.6 |
| Country(ies) of Participants’ Residence | Canada |
| Number of participants randomised | Children/ participants: 536; Caregivers NA |
| Gender demographics of participants | 71.3% female |
| Socioeconomic status of participants | Not reported |
| Race/Ethnicity of participants | Not reported |
| Type of participant targeted | Adolescents at risk for anxiety |
| Children’s advisory group formation | (Was an advisory group formed in the planning/execution of the study?) Yes ☐ No ☐ Unclear ☐ Not Applicable ☐ Not Reported ☒ Ages of advisory group participants \| Not reported; Means of recruiting advisory group participants \| Not reported; Phases of trial involvement \| Not reported |
| Inclusion/ Exclusion Criteria | Included: minimum score of 25 on the Screen for Child Anxiety Related Disorders (screening was via REDCap), ability to read and write English, regular access to a telephone and a computer system with high-speed internet, the ability to use the computer to interact with web material  Excluded: screened as high-rsk for self-harm via four items from the Ask Suicide-Screening Questionnaire, indicated the possible presence of a psychosis-related disorder via the 5-item Schizophrenia Test and Early Psychosis Indicator, screened positive for harmful or hazardous alcohol consumption via the 3-item Alcohol Use Disorders Identification Test Consumption subscale, resided outside of CA. |
| **Methods of Recruitment, Retention & Consent** | |
| Online methods/platforms used for recruitment | Recruitment was through Breathe’s social media platforms (Facebook, Twitter, Tumblr, and Instagram) with posts and paid ads across CA. |
| Offline methods used for recruitment | Health care professionals provided study pamphlets to prospective participants seeking mental health care in specialty care clinics, primary care clinics, and schools. |
| Methods/tools used for online consent/assent acquisition | Via REDCap |
| Caregiver consent acquired/ waived (If waived, by what authority?) | Yes ☒ No ☐ Unclear ☐ Not Applicable ☐ Not Reported ☐  “adolescents aged 13 and 14 years required online parental consent in addition to their assent to participate. Parental consent followed the same Web-based process described for adolescents.” Pg.3 |
| Children assent acquisition | Yes ☒ No ☐ Unclear ☐ Not Applicable ☐ Not Reported ☐ |
| What, if any, methods were used to validate that assent/consent was informed? | “Adolescents were provided an information sheet on the trial and asked several yes/no questions to ensure consent/assent was informed.” Pg. 3  (E.g. validation questions, CAPTCHA, email confirmation, video-conferencing verbal) |
| Pushes/reminders sent to participants | Enrolment in trial Yes ☐ No ☒ Unclear ☐ Not Applicable ☐ Not Reported ☐; Retention/completion in trial Yes ☒ No ☐ Unclear ☐ Not Applicable ☐ Not Reported ☐; Follow-up in trial Yes ☐ No ☒ Unclear ☐ Not Applicable ☐ Not Reported ☐ Weekly emails were provided to help users continue with the program and provide notifications of the release of new sessions. |
| **Interventions** | |
| Number of arms | 2 |
| Intervention | “The program consisted of six iCBT sessions, with each session requiring approximately 30 min to complete; it was suggested that participants complete one session per week in a location convenient for them.” Pg. 3 |
| Comparison | The control group received resource-based webpages with suggestions of anxiety-based books and educational websites, contact information for local and national crisis lines, and information on the emergency department and other crisis mental health resources. |
| Operating Systems/Devices required for participants’ engagement with trial | Any device with internet access  (E.g. computer, phone, iOS, Windows, Android, iPhone, Facebook or Instagram account etc.) |
| Tools/software used for data protection processes | Intelligent Research and Intervention Software (IRIS), a secure, password-protected website. “Data collection was embedded in IRIS to allow for electronically captured, securely stored, encrypted, and password-protected data.” Pg. 6  (E.g. dedicated website, REDCap, institutional server etc.) |
| Tools/methods used for data collection | Intelligent Research and Intervention Software (IRIS) hosted both the Breathe website and the static webpage  (E.g. data submitted via website, video-conferencing etc.) |
| Did caregivers participate/ contribute data to the trial? If so, describe. | Yes ☐ No ☒ Unclear ☐ Not Applicable ☐ Not Reported ☐  “Users were also provided with the option for a summary of each session to be emailed to an identified parent or guardian after each completed session.” Pg.5 |
| Duration of intervention from first to final engagement | 6 weeks |
| Compensation offered to participants | “Adolescents who completed outcome measures at the postintervention time point were given a token of appreciation (Can $25 electronic gift card).” Pg.6 |
| **Outcomes** | |
| Results relevant to this scoping review | Intervention respondents rated the program at 62/84 (Good)  (E.g. satisfaction with online methods of trial, demographics of participants’ in relation to their outcomes especially in regarding online components, i.e., recruitment, retention, completion etc)  Control respondents rated the control webpage at 51.2/84 (Moderate) |
| Baseline differences between completers & non-completers | Not reported |
| Attrition rates of randomised participants | Intervention: 258  Control: 278  TX: 50/258 completed intervention: 80.6% loss but the authors considered completing 4 sessions to be an active participant, therefore 72 completed at least 4: 72.1% loss  CO:196/278 visited at least one site:29.5% loss |
| Limitations of study as described by authors | Mainly female respondents, very high attrition rates. |
| Funding source | Institutional, i.e. governmental, university \| Yes ☒ No ☐ Unclear ☐ Not Applicable ☐ Not Reported ☐; Private, i.e. pharmaceutical, software company \| Yes ☐ No ☒ Unclear ☐ Not Applicable ☐ Not Reported ☐ |
| Other aspects of study pertinent to online trial conception and execution | “As part of the Breathe program, adolescents received one telephone-based coaching call after completing their first session to prepare adolescents for the skills-based program activities to follow, including exposure activities, that would begin in session 2.” But it was not required to continue with the program and some avoided it as they found it stressful. Pg.16 |

| Risk of Bias assessment  ([**https://handbook-5-1.cochrane.org/chapter_8/8_assessing_risk_of_bias_in_included_studies.htm**](https://handbook-5-1.cochrane.org/chapter_8/8_assessing_risk_of_bias_in_included_studies.htm)**)** | | | | |
| --- | --- | --- | --- | --- |
| Bias domain | Source of bias | Risk of bias | Support for judgment (use direct quotes where possible with explanatory comments) | Location in text or source (pg. number, figure, table etc.) |
| Selection bias | Random sequence generation | Low risk ☒    High risk ☐  Unclear risk ☐ | “Once consent and assent were obtained, adolescents were enrolled in the trial and randomly assigned using a computer-generated sequence with a 1:1 allocation ratio to either the Breathe program or the resource-based webpages.” | Pg. 3, under Procedures for Informed Consent and Assent |
|  | Allocation concealment | Low risk ☒    High risk ☐  Unclear risk ☐ | Comment: No mention of blinding of assessors in the text, but according to the ClinicalTrials.gov register the investigators were blind. It is unclear whether this refers to outcome assessments or allocation of participants, and therefore there is not enough information to make an assessment. | Not reported |
| Performance bias | Blinding of participants and personnel | Low risk ☐    High risk ☒  Unclear risk ☐ | “This was an open-label trial, and adolescents were notified of their assigned intervention via an email that included instructions for logging into the study website.”  Comment: The lack of blinding likely influenced the outcome. | Pg. 3, under Procedures for Informed Consent and Assent |
| Detection bias | Blinding of outcome assessment | Low risk ☐    High risk ☐  Unclear risk ☒ | Comment: No mention of blinding of assessors in the text, but according to the ClinicalTrials.gov register the investigators were blind. It is unclear whether this refers to outcome assessments or allocation of participants, and therefore there is not enough information to make an assessment. | Not reported |
| Attrition bias | Incomplete outcome data | Low risk ☐    High risk ☒  Unclear risk ☐ | Comment:  Intervention:  72/258 completed 6-wk follow-up: 72.1% attrition rate  Control: 196/278 completed 6-wk follow-up: 29.5% attrition rate | Pgs. 9, 10, Tables 4 and 5 |
| Reporting bias | Selective reporting | Low risk ☒    High risk ☐  Unclear risk ☐ | Comment: No study protocol available but the study is registered at ClinicalTrials.gov and all outcomes were assessed as pre-specified. | ClinicalTrials.gov: NCT02970734 |
| Other bias | Anything else, ideally pre-specified | Low risk ☒    High risk ☐  Unclear risk ☐ | Comment: No other bias detected. | Not applicable |
