## Supplementary material for "Online randomised trials with children: A scoping review": S7 Appendix: Schleider.docx

| **Basic Study Characteristics** | |
| --- | --- |
| **Citation** | Schleider JL, Mullarkey MC, Fox KR, Dobias ML, Shroff A, Hart EA, and Roulston CA. A randomized trial of online single-session interventions for adolescent depression during COVID-19.2022. Nature Human Behaviour, 6, pp.258-268. |
| **Characteristic reported (indicate with X for decision or enter free text, where applicable)** | |
| Type of study | Randomised trial ☒ \| Cluster randomised ☐; Quasi-randomised trial ☐ \| Individually randomised ☒ |
| Type of publication | Peer-reviewed journal ☒; Pre-print ☐; Other, if so, describe ☐ |
| Aim of study as per author’s description | There were three aims:   1) To test if an online, self-guided SSI (single-session intervention) would improve hopelessness, perceived agency, and three-month depressive symptoms compared to an active control condition and 2) To test whether the GM-SSI or the BA-SSI is more impactful in this context and 3) To test whether either SSI improved COVID-19-related trauma, hopelessness, agency and generalized anxiety compared to the control |
| **Participant Characteristics** | |
| Participant age demographics | Age range \| 13-16; Mean age \| 14.82 |
| Country(ies) of Participants’ Residence | U.S.A. |
| Number of participants randomised | Children/ participants: 2,452; Caregivers NA |
| Gender demographics of participants | 88.09% female  80% of adolescents identified with a sexual minority identity |
| Socioeconomic status of participants | Not reported |
| Race/Ethnicity of participants | 3.75% American Indian  12.64% Asian  10.48% Black  1.59% Native Hawaiian  19.21% Hispanic  66.56% White (Hispanic and White were not mutually exclusive) |
| Type of participant targeted | Adolescents with elevated depression symptoms. |
| Children’s advisory group formation | Ages of advisory group participants \| No; Means of recruiting advisory group participants \| No; Phases of trial involvement \| No Yes ☐ No ☒ Unclear ☐ Not Applicable ☐ Not Reported ☐ (Was an advisory group formed in the planning/execution of the study?) |
| Inclusion/ Exclusion Criteria | Included: comfortable reading and writing in English, internet and computer, laptop or smartphone access, endorsement of elevated depressive symptoms (>/= 2 on PHQ-2) |
| **Methods of Recruitment, Retention & Consent** | |
| Online methods/platforms used for recruitment | Recruitment was through Instagram ads following established ethics guidelines for passive, social-media-based recruitment. Ads linked to a Qualtrics survey. |
| Offline methods used for recruitment | NA |
| Methods/tools used for online consent/assent acquisition | Qualtrics |
| Caregiver consent acquired/ waived (If waived, by what authority?) | Yes ☐ No ☒ Unclear ☐ Not Applicable ☐ Not Reported ☐  “To maintain adolescents’ confidentiality and minimize access barriers (for example, discomfort disclosing psychological distress, as parents are often unaware of their adolescents’ depressive symptoms, including suicidal ideation, in up to 80% of cases)39, parent permission was not required to participate in this study (waived by the university institutional review board).” Pg. 265 |
| Children assent acquisition | Yes ☒ No ☐ Unclear ☐ Not Applicable ☐ Not Reported ☐ |
| What, if any, methods were used to validate that assent/consent was informed? | No  (E.g. validation questions, CAPTCHA, email confirmation, video-conferencing verbal) |
| Pushes/reminders sent to participants | Enrolment in trial Yes ☐ No ☒ Unclear ☐ Not Applicable ☐ Not Reported ☐; Retention/completion in trial Yes ☐ No ☒ Unclear ☐ Not Applicable ☐ Not Reported ☐; Follow-up in trial Yes ☒ No ☐ Unclear ☐ Not Applicable ☐ Not Reported ☐ “Youths then received an email invitation three months later to complete a 10-minute follow-up questionnaire” Pg. 265 |
| **Interventions** | |
| Number of arms | 3 |
| Intervention | 1) The BA-SSI (Behaviour Activation: ABC Project) had 5 elements: intro to programme’s rationale, psychoeducation about depression, life values assessment, creation of an activity action plan, writing exercise 2) The GM-SSI (Growth mind-set: Project Personality) had 5 elements: an intro to brain and neuroplasticity, testimonials from older youths, further stories about growth mind-sets, study summaries, writing exercise   Additionally, the participants received a resource list of hotlines, text-lines and online psychoeducational resources to facilitate engagement with additional mental health supports, if desired. Adolescents were also invited to contact the research team at any time during the study with questions or for further support in accessing mental health support beyond the study’s scope |
| Comparison | Supportive Therapy SSI (placebo) was matched in length and peer narratives to interventions but was designed to control for non-specific aspects of completing a generally supportive online activity. After all data collection was complete, the researchers learned each youth’s condition assignment, and both active interventions were offered to all participants.  “Additionally, the participants received a resource list of hotlines, text-lines and online psychoeducational resources to facilitate engagement with additional mental health supports, if desired. Adolescents were also invited to contact the research team at any time during the study with questions or for further support in accessing mental health support beyond the study’s scope.” Pg. 265 |
| Operating Systems/Devices required for participants’ engagement with trial | Any internet-equipped device  (E.g. computer, phone, iOS, Windows, Android, iPhone, Facebook or Instagram account etc.) |
| Tools/software used for data protection processes | Qualtrics  (E.g. dedicated website, REDCap, institutional server etc.) |
| Tools/methods used for data collection | Qualtrics  (E.g. data submitted via website, video-conferencing etc.) |
| Did caregivers participate/ contribute data to the trial? If so, describe. | Yes ☐ No ☒ Unclear ☐ Not Applicable ☐ Not Reported ☐ |
| Duration of intervention from first to final engagement | SSI + 3-month follow-up |
| Compensation offered to participants | “The posts included invitations to determine eligibility for a confidential, online psychology study, for which participants could earn up to $20USD in gift cards.” Pg. 265 |
| **Outcomes** | |
| Results relevant to this scoping review | NA  (E.g. satisfaction with online methods of trial, demographics of participants’ in relation to their outcomes especially in regarding online components, i.e., recruitment, retention, completion etc) |
| Baseline differences between completers & non-completers | “Thus, the overall results patterns were similar—showing only minor differences with respect to secondary outcomes—regardless of our approach to handling missing data.” Pg. 263 |
| Attrition rates of randomised participants | \| Randomised \| Intervention received \| % attrition \| 3-month FU completed \| % attrition; Control \| 818 \| 669 \| 18.2 \| 455 \| 26.2; BA-SSI \| 821 \| 732 \| 10.8 \| 480 \| 30.7; GM-SSI \| 813 \| 653 \| 19.7 \| 433 \| 27.1 |
| Limitations of study as described by authors | Limited to English, some groups of youth were over-represented in our sample (for example, sexual minority youth), whereas others were underrepresented (for example, boys). |
| Funding source | Institutional, i.e. governmental, university \| Yes ☒ No ☐ Unclear ☐ Not Applicable ☐ Not Reported ☐; Private, i.e. pharmaceutical, software company \| Yes ☐ No ☒ Unclear ☐ Not Applicable ☐ Not Reported ☐ |
| Other aspects of study pertinent to online trial conception and execution | “Notably, our final sample (N=2,452) was 52 participants greater than our pre-registered sample size, as a group of youths completed our eligibility screener within hours of a study advertisement gaining traction on Instagram. We stopped recruitment as soon as we learned that our pre-registered threshold had been met.” Pg. 266 |

| Risk of Bias assessment  ([**https://handbook-5-1.cochrane.org/chapter_8/8_assessing_risk_of_bias_in_included_studies.htm**](https://handbook-5-1.cochrane.org/chapter_8/8_assessing_risk_of_bias_in_included_studies.htm)**)** | | | | |
| --- | --- | --- | --- | --- |
| Bias domain | Source of bias | Risk of bias | Support for judgment (use direct quotes where possible with explanatory comments) | Location in text or source (pg. number, figure, table etc.) |
| Selection bias | Random sequence generation | Low risk ☒    High risk ☐  Unclear risk ☐ | “youths were randomly assigned to one of three SSIs in a 1:1:1 ratio per a Qualtrics-embedded randomizer” | Pg. 265, under Procedures |
|  | Allocation concealment | Low risk ☒    High risk ☐  Unclear risk ☐ | “youth and investigators were masked to condition assignment until after all data collection was complete” | Pg.265, under Procedures |
| Performance bias | Blinding of participants and personnel | Low risk ☒    High risk ☐  Unclear risk ☐ | “After all data collection was complete, the researchers learned each youth’s condition assignment”  “The data collection procedure occurred entirely online (phone, tablet, computer, or any internet-connected device), and the participants/researchers were masked to condition assignment as the randomization occurred automatically within the Qualtrics survey platform.” | Pg.265, under Procedures  Pg. 2 of reporting summary, under Behavioural & social sciences study design: Data collection |
| Detection bias | Blinding of outcome assessment | Low risk ☐    High risk ☐  Unclear risk ☒ | Comment: Insufficient information to make an assessment | Not reported |
| Attrition bias | Incomplete outcome data | Low risk ☒    High risk ☐  Unclear risk ☐ | Comment:  Intervention: 669/818 received intervention: 18.2% attrition  455/818 3-mo follow-up: 26.2% attrition  BA-SSI: 732/821 received intervention: 10.8% attrition  480/821 completed 3-mo follow-up: 30.7% attrition  GM-SSI: 653/813 received intervention: 19.7% attrition  433/813 completed 3-mo follow-up: 27.1% attrition | Pg. 263, Fig. 2 |
| Reporting bias | Selective reporting | Low risk ☒    High risk ☐  Unclear risk ☐ | Comment: All primary and secondary trial outcomes were pre-registered in the ClinicalTrials.gov registration. | Pg. 266, Outcomes  ClinicalTrials.gov: NCT04634903 |
| Other bias | Anything else, ideally pre-specified | Low risk ☒    High risk ☐  Unclear risk ☐ | No other bias detected | Not applicable |
