## Supplementary material for "Online randomised trials with children: A scoping review": S7 Appendix: Schwinn.docx

| **Basic Study Characteristics** | |
| --- | --- |
| **Citation** | Schwinn TM, Thom B, Schinke SP, Hopkins J. Preventing drug use among sexual-minority youths: findings from a tailored, web-based intervention. Journal of adolescent health. 2015;56(5):571-3. doi: 10.1016/j.jadohealth.2014.12.015. PubMed PMID: CN-01078656. |
| **Characteristic reported (indicate with X for decision or enter free text, where applicable)** | |
| Type of study | Randomised trial ☒ \| Cluster randomised ☐; Quasi-randomised trial ☐ \| Individually randomised ☒ |
| Type of publication | Peer-reviewed journal ☒; Pre-print ☐; Other, if so, describe ☐ |
| Aim of study as per author’s description | To test   1) The efficacy of a tailored intervention for drug use and associated risk factors among sexual-minority youths 2) The feasibility of internet recruitment procedures 3) The feasibility of data collection over multiple time points and retaining participants with minimal participant contact information |
| **Participant Characteristics** | |
| Participant age demographics | Age range \| 15-16; Mean age \| 16.08 |
| Country(ies) of Participants’ Residence | U.S.A. |
| Number of participants randomised | Children/ participants: 236; Caregivers NA |
| Gender demographics of participants | 32.1% male  49.6% female  Queer/fluid/other 18.3% |
| Socioeconomic status of participants | Not reported |
| Race/Ethnicity of participants | 6.4% Asian  7.3% Black  7.4% Other  12.8% Hispanic  66.1% White |
| Type of participant targeted | Sexual-minority youth |
| Children’s advisory group formation | (Was an advisory group formed in the planning/execution of the study?) Yes ☐ No ☒ Unclear ☐ Not Applicable ☐ Not Reported ☐ Ages of advisory group participants \| No; Means of recruiting advisory group participants \| No; Phases of trial involvement \| No |
| Inclusion/ Exclusion Criteria | Included: U.S. resident, access to a personal computer, identify as gay, lesbian, bisexual, transgender, or questioning |
| **Methods of Recruitment, Retention & Consent** | |
| Online methods/platforms used for recruitment | Six ads ran on Facebook for 9 days |
| Offline methods used for recruitment | NA |
| Methods/tools used for online consent/assent acquisition | Secure website |
| Caregiver consent acquired/ waived (If waived, by what authority?) | Yes ☐ No ☒ Unclear ☐ Not Applicable ☐ Not Reported ☐  “Columbia University’s institutional review board approved the study procedures and granted a waiver of parental permission.” Pg. 572 |
| Children assent acquisition | Yes ☒ No ☐ Unclear ☐ Not Applicable ☐ Not Reported ☐ |
| What, if any, methods were used to validate that assent/consent was informed? | Correctly answer a five-question quiz on study procedures  (E.g. validation questions, CAPTCHA, email confirmation, video-conferencing verbal) |
| Pushes/reminders sent to participants | Enrolment in trial Yes ☐ No ☒ Unclear ☐ Not Applicable ☐ Not Reported ☐; Retention/completion in trial Yes ☐ No ☒ Unclear ☐ Not Applicable ☐ Not Reported ☐; Follow-up in trial Yes ☐ No ☐ Unclear ☐ Not Applicable ☐ Not Reported ☒ Not described, but there must have been a reminder to complete follow-up. |
| **Interventions** | |
| Number of arms | 2 |
| Intervention | “The three-session intervention was guided by a social competency skill-building strategy and minority stress theory. An animated young adult narrator led youths through the tailored content and practice scenarios that included interactive games, role-playing, and writing activities.” Pg. 572 |
| Comparison | “Control” |
| Operating Systems/Devices required for participants’ engagement with trial | E-mail address, personal computer access  (E.g. computer, phone, iOS, Windows, Android, iPhone, Facebook or Instagram account etc.) |
| Tools/software used for data protection processes | “After enrollment, youths entered their names, birthdates, e-mail addresses, and cell phone numbers (optional) onto a secure webpage.” Pg. 572  (E.g. dedicated website, REDCap, institutional server etc.) |
| Tools/methods used for data collection | “After enrollment, youths entered their names, birthdates, e-mail addresses, and cell phone numbers (optional) onto a secure webpage.” Pg. 572  (E.g. data submitted via website, video-conferencing etc.) |
| Did caregivers participate/ contribute data to the trial? If so, describe. | Yes ☐ No ☒ Unclear ☐ Not Applicable ☐ Not Reported ☐ |
| Duration of intervention from first to final engagement | 4 weeks + 3-month follow-up |
| Compensation offered to participants | “Youths received online gift cards of $25, $30, and $45, for pretest, posttest, and 3-month follow-up, respectively.” Pg. 572 |
| **Outcomes** | |
| Results relevant to this scoping review | (E.g. satisfaction with online methods of trial, demographics of participants’ in relation to their outcomes especially in regarding online components, i.e., recruitment, retention, completion etc)  NA |
| Baseline differences between completers & non-completers | “Chi-square and t tests indicated that intervention and control youths were comparable on measures of pretest variables (Table 1).” Pg. 572 |
| Attrition rates of randomised participants | \| Randomised \| Post-test completed \| 3-month FU completed \| % attrition; Intervention \| 119 \| “82%” \| 97 \| 18.5; Control \| 117 \| NR \| 103 \| 12 |
| Limitations of study as described by authors | “Study limitations include small sample size, small program effects, short follow-up, brief intervention, and self-report drug use measures. Although adequately powered to detect changes between study arms, the small sample size precluded analysis by gender, disclosure status, and other covariates.” Pg. 573 |
| Funding source | Institutional, i.e. governmental, university \| Yes ☒ No ☐ Unclear ☐ Not Applicable ☐ Not Reported ☐; Private, i.e. pharmaceutical, software company \| Yes ☐ No ☒ Unclear ☐ Not Applicable ☐ Not Reported ☐ |
| Other aspects of study pertinent to online trial conception and execution | NA |

| Risk of Bias assessment  ([**https://handbook-5-1.cochrane.org/chapter_8/8_assessing_risk_of_bias_in_included_studies.htm**](https://handbook-5-1.cochrane.org/chapter_8/8_assessing_risk_of_bias_in_included_studies.htm)**)** | | | | |
| --- | --- | --- | --- | --- |
| Bias domain | Source of bias | Risk of bias | Support for judgment (use direct quotes where possible with explanatory comments) | Location in text or source (pg. number, figure, table etc.) |
| Selection bias | Random sequence generation | Low risk ☐    High risk ☐  Unclear risk ☒ | “Randomly, youths were assigned to the intervention arm or to the control arm.”  Comment: insufficient information to make an assessment | Pg. 572, under Methods |
|  | Allocation concealment | Low risk ☐    High risk ☐  Unclear risk ☒ | Comment: Insufficient evidence to make an assessment. | Not reported |
| Performance bias | Blinding of participants and personnel | Low risk ☐    High risk ☐  Unclear risk ☒ | Comment: Insufficient evidence to make an assessment. | Not reported |
| Detection bias | Blinding of outcome assessment | Low risk ☐    High risk ☐  Unclear risk ☒ | Comment: Insufficient evidence to make an assessment. | Not reported |
| Attrition bias | Incomplete outcome data | Low risk ☒    High risk ☐  Unclear risk ☐ | Comment: Intervention: 97/119 completed 3-mo follow-up: 18.5% attrition  Control: 103/117 completed 3-mo follow-up: 12% attrition | Pg. 572, 572, Tables 1 & 2 |
| Reporting bias | Selective reporting | Low risk ☐    High risk ☐  Unclear risk ☒ | Comment: There is no study protocol or trial register available and therefore insufficient information to make an assessment. | Not reported |
| Other bias | Anything else, ideally pre-specified | Low risk ☒    High risk ☐  Unclear risk ☐ | Comment: No other bias detected | Not applicable |
